# Vaccine Induction of Heterologous HIV-1 Neutralizing Antibody B Cell Lineages in Humans

**DOI:** 10.1101/2023.03.09.23286943

**Authors:** Wilton B. Williams, S. Munir Alam, Gilad Ofek, Nathaniel Erdmann, David Montefiori, Michael S. Seaman, Kshitij Wagh, Bette Korber, Robert J. Edwards, Katayoun Mansouri, Amanda Eaton, Derek W. Cain, Mitchell Martin, Robert Parks, Maggie Barr, Andrew Foulger, Kara Anasti, Parth Patel, Salam Sammour, Ruth J. Parsons, Xiao Huang, Jared Lindenberger, Susan Fetics, Katarzyna Janowska, Aurelie Niyongabo, Benjamin M. Janus, Anagh Astavans, Christopher B. Fox, Ipsita Mohanty, Tyler Evangelous, Yue Chen, Madison Berry, Helene Kirshner, Elizabeth Van Itallie, Kevin Saunders, Kevin Wiehe, Kristen W. Cohen, M. Juliana McElrath, Lawrence Corey, Priyamvada Acharya, Stephen R. Walsh, Lindsey R. Baden, Barton F. Haynes

**Affiliations:** Duke Human Vaccine Institute, Duke School of Medicine, Durham, NC 27710; Department of Surgery, Duke School of Medicine, Durham, NC 27710; Department of Cell Biology and Molecular Genetics, University of Maryland, College Park, MD 20742; Department of Medicine, Duke School of Medicine, Durham, NC 27710; Department of Integrative Immunobiology, Duke School of Medicine, Durham, NC 27710; Department of Molecular Genetics and Microbiology, Duke School of Medicine, Durham, NC 27710; Fred Hutchinson Cancer Center, Seattle WA 98109; Brigham and Women’s Hospital, Harvard Medical School, Boston, MA 02115; University of Alabama Medical Center, Birmingham, AL 35223; Department of Biochemistry, Duke School of Medicine, Durham, NC 27710; Duke Global Health Institute, Duke School of Medicine, Durham, NC 27710; Beth Israel Deaconess Medical Center, Harvard Medical School, Boston, MA 02215; Access to Advanced Health Institute, Seattle, WA, 98102; Los Alamos National Laboratory, Los Alamos, NM, 87545; New Mexico Consortium, Los Alamos, NM, 87544; Institute for Bioscience and Biotechnology Research, University of Maryland, Rockville, MD 20850

## Abstract

A critical roadblock to HIV vaccine development is the inability to induce B cell lineages of broadly neutralizing antibodies (bnAbs) in humans. In people living with HIV-1, bnAbs take years to develop. The HVTN 133 clinical trial studied a peptide/liposome immunogen targeting B cell lineages of HIV-1 envelope membrane proximal external region (MPER) bnAbs (NCT03934541). Here, we report MPER peptide-liposome induction of polyclonal HIV-1 B cell lineages of mature bnAbs and their precursors, the most potent of which neutralized 15% of global tier 2 HIV-1 strains and 35% of clade B strains, with lineage initiation after the second immunization. Neutralization was enhanced by vaccine-selection of improbable mutations that increased antibody binding to gp41 and lipids. This study demonstrates proof-of-concept for rapid vaccine induction of human B cell lineages with heterologous neutralizing activity and selection of antibody improbable mutations, and outlines a path for successful HIV-1 vaccine development.

## Introduction

Development of a preventive HIV-1 vaccine is a global priority. However, in ten HIV-1 vaccine efficacy trials, none showed protection from HIV-1 except the RV144 trial in Thailand that showed 31.2% estimated efficacy that was insufficient to deploy the vaccine ^1–4^. Moreover, a vaccine similar to that used in RV144 was tested against clade C HIV-1 in South Africa and showed no efficacy ^5^. HIV-1 envelope (Env)-containing vaccines tested to date have only induced non-neutralizing antibodies (nnAbs) ^1, 6^, and no previous vaccine trials induced broadly neutralizing antibodies (bnAbs) ^3, 7–9^. With the failure of nnAb vaccines, attention has turned to developing HIV-1 vaccine regimens that can induce heterologous HIV-1 neutralizing antibodies as a major priority ^7, 8^.

Most bnAbs identified to date from people living with HIV-1 (PLWH) have unusual traits, such as long third heavy chain complementary determinant regions (HCDR3s), high levels of rare, improbable antibody maturation mutations that are required for neutralization breadth ^10–12^, and/or polyreactivity with host or environmental antigens ^13–15^. The unusual traits required for bnAb activity can be disfavored by tolerance mechanisms of the immune system, such as deletion or anergy ^16–19^. Moreover, the HIV-1 Env is heavily glycosylated with host, high mannose and complex glycans that are poorly immunogenic and shield a major portion of the Env surface ^20^.

BnAbs protect monkeys from simian human immunodeficiency virus (SHIV) challenges ^21–23^, and protect against susceptible HIV-1 strains in humans ^24^. One Env target for bnAb induction is the gp41 membrane proximal external region (MPER) ^25^. MPER targeted antibodies are among bnAbs with the best neutralization breadth ^26–30^. The MPER is physically located near the virion lipid membrane, and the lipid membrane is a component of the epitopes for both proximal (with the prototype bnAb 2F5, with a core epitope of ELDKWA, located at Env positions 662-667 in HXB2) and distal MPER bnAbs (the prototype bnAb was 4E10 with a core epitope of NWFDIT, located at positions 671-676 in HXB2) ^13, 31–34^. Binding to lipids has been proposed to tether the bnAb on the virion membrane to enable binding to gp41 neutralizing MPER bnAb epitopes upon their exposure by Env receptor-mediated activation ^32^. A neutralization assay that detects MPER bnAb precursors has been developed that promotes tethering of MPER-reactive antibodies to cell surface via FcγRI/CD64 expression in the TZM-bl infection indicator cell line ^35–37^. While lipid binding is an essential component for the broad neutralizing activity of MPER bnAbs, induction of antibodies to self-lipids has been a substantial roadblock in MPER bnAb elicitation ^13, 38^.

Potent bnAbs develop in PLWH, but only rarely, and after many months to years after transmission ^11, 12, 39, 40^. A strategy for inducing bnAbs, termed B cell lineage immunogen design, aims to more rapidly induce bnAbs by selecting Envs that can bind with high affinity to bnAb lineage naïve or unmutated common ancestor (UCA) B cell lineage precursors, and then guide their bnAb development from early intermediate to mature bnAbs via sequential immunizations ^3, 15^. Immunogens that bind to CD4-binding site UCA B cell receptors can expand bnAb precursors in animal models ^22, 41–43^ and now in humans ^44^. A second strategy is epitope-focused immunogen design whereby a linear peptide such as the HIV-1 fusion domain is targeted by repeated immunizations ^36, 45–47^. Wang and colleagues have shown that such peptide immunogens can expand HIV-1 fusion domain lineage precursors in humans ^48^. However, to date, there has been no success in inducing tier 2 neutralizing heterologous antibodies in humans. We have designed a MPER peptide-liposome with proximal and distal MPER bnAb epitopes ^47, 49, 50^ and demonstrated induction of antibodies in rhesus macaques that neutralize HIV-1 in the MPER bnAb precursor-sensitive TZM-bl/FcγRI neutralization assay, but did not neutralize HIV-1 in the more rigorous TZM-bl assay ^36^. The TZM-bl/FcγRI assay identifies MPER bnAb precursors by incorporating CD64 (FcγRI) into TZM-bl cells that binds MPER antibody via Fc and enhances neutralization potency when the neutralizing antibody paratope binds the MPER bnAb epitope ^32^.

An additional factor that limits bnAb development is the requirement for bnAbs to accumulate high levels of appropriate functional improbable mutations—mutations that are essential for broad neutralization activity in a given lineage but are not frequently made by somatic hypermutation ^10^. During HIV-1 infection, bnAb B cell lineages only develop after years, due in part to the requirement of multiple rounds of Env escape from coevolving bnAb lineage intermediates for selecting B cells with B cell receptors encoding the combination of improbable and probable mutations that are required to achieve heterologous neutralization breadth ^11^. B cell lineage immunogen design seeks to circumvent this problem by designing immunogens capable of rapidly selecting for improbable bnAb B cell receptor mutations via vaccination, but to date this has not been accomplished in humans.

Here, we demonstrate that an MPER peptide-liposome that binds to a bnAb unmutated ancestor (UA) antibody could induce in humans polyclonal MPER epitope-targeted antibodies that neutralize heterologous HIV-1 strains. The most potent vaccine-induced MPER clonal B cell lineage underwent affinity maturation after three immunizations to acquire neutralization of 15% of global or heterologous tier 2 HIV-1 strains, and 35% of clade B HIV-1 strains. Moreover, we demonstrate that the MPER peptide-liposome rapidly selected antibody functional improbable mutations that were essential for heterologous HIV-1 neutralization and we determine the contributions of the individual mutations to MPER and lipid binding for mediating neutralization breadth and potency.

## Results

### HVTN 133 Clinical Trial

The immunogen used in the HVTN 133 trial was a MPER peptide-liposome designed to bind to the UA of prototype proximal MPER bnAb, 2F5 ^47, 49, 50^. The HVTN 133 (NCT03934541) clinical trial tested the MPER peptide-liposome in 20 HIV-1 naïve individuals; five vaccinees in a 500 mcg vaccine low dose group and 15 vaccinees in the 2000 mcg vaccine high dose group ^51^. The trial enrolled 20 vaccinees and 4 placebo recipients with the aim of immunizing each individual four times. However, after 20 vaccinees received two immunizations and five vaccinees received three immunizations, one vaccinee developed an anaphylaxis reaction 4 hours after their third immunization that required antihistamine and steroid treatment for symptom resolution ^51^. This reaction was determined to be most likely due to allergy to polyethylene glycol (PEG), because of a vaccine-induced rise in titer to PEG, and sensitization of a vaccinee’s neutrophils by PEG antibodies to degranulate upon exposure to vaccine-matched peptide-liposomes ^51^.

Nonetheless, the HVTN 133 clinical trial was successful in inducing a 95% serum response rate to the immunogen and 100% blood CD4+ T cell response rate after two immunizations ^51^. Moreover, 13/20 (65%) vaccinees had serum responses after two immunizations that mapped to the MPER bnAb epitope ^51^. Finally, weak heterologous tier 2 HIV-1 neutralizing activity was detected in serum of 2 of 5 individuals who received three immunizations before the trial was stopped (49). Here, we have performed a B cell repertoire analysis in the five individuals in HVTN 133 that received 3 immunizations and determined the characteristics and mechanisms of neutralization of induced tier 2 heterologous neutralizing antibodies.

### Frequency of MPER-reactive B Cells in Blood

First, we studied the peripheral blood B cell repertoire of the 5 vaccinees and one placebo recipient who received three immunizations for the presence of MPER-reactive (MPER+) B cells. We isolated 510 B cells from the 5 HVTN133 vaccine trial participants after the 3^rd^ immunization via antigen-specific single B cell sorting with fluorophore-labeled MPER-peptide tetramers (**Figures 1a-c**). After expression of B cell receptors (BCRs) as recombinant IgG1 antibodies, we identified 87 MPER+ antibodies that bound to vaccine-matched MPER656 (^656^NEQELLELDKWASLWNWFNITNWLW^680^) or MPER.03 (KKK^656^NEQELLELDKWASLWNWFDITNWLWYI^682^RKKK) peptides (**Table S1**). For identification of antibodies that had the highest probability of membership in bnAb B cell lineages, we assayed for MPER-differential (Δ) binding antibodies that bound to a wild-type (WT) MPER.03 peptide and had ≥2.5-fold reduction in binding to a mutant MPER.03 peptide with knockout (KO) mutations of MPER bnAb epitopes (D664A, W672A) ^36^ (**Tables S1-S3**). MPERΔ antibodies constituted 86% (75/87) of MPER+ antibodies (**Tables S1-S3**). While all five vaccinees who received three immunizations (133-23, 133-30, 133-33, 133-35 and 133-39) were responders with regard to induction of serum responses to the immunizing peptide liposome ^51^, only vaccinees 133-23 and 133-39 in the low dose group were high responders regarding isolation of MPER+ memory B cells (**Tables S1-S2**). Immunization with the MPER peptide-liposome boosted the frequency of MPER+ antibodies from ≤ ∼1 in 1,000,000 B cells at baseline to ∼1 in 11,100 B cells in vaccinee 133-23 and to ∼1 in 25,000 B cells in vaccinee 133-39 after three immunizations (**Table S2**).

**Figure 1.**
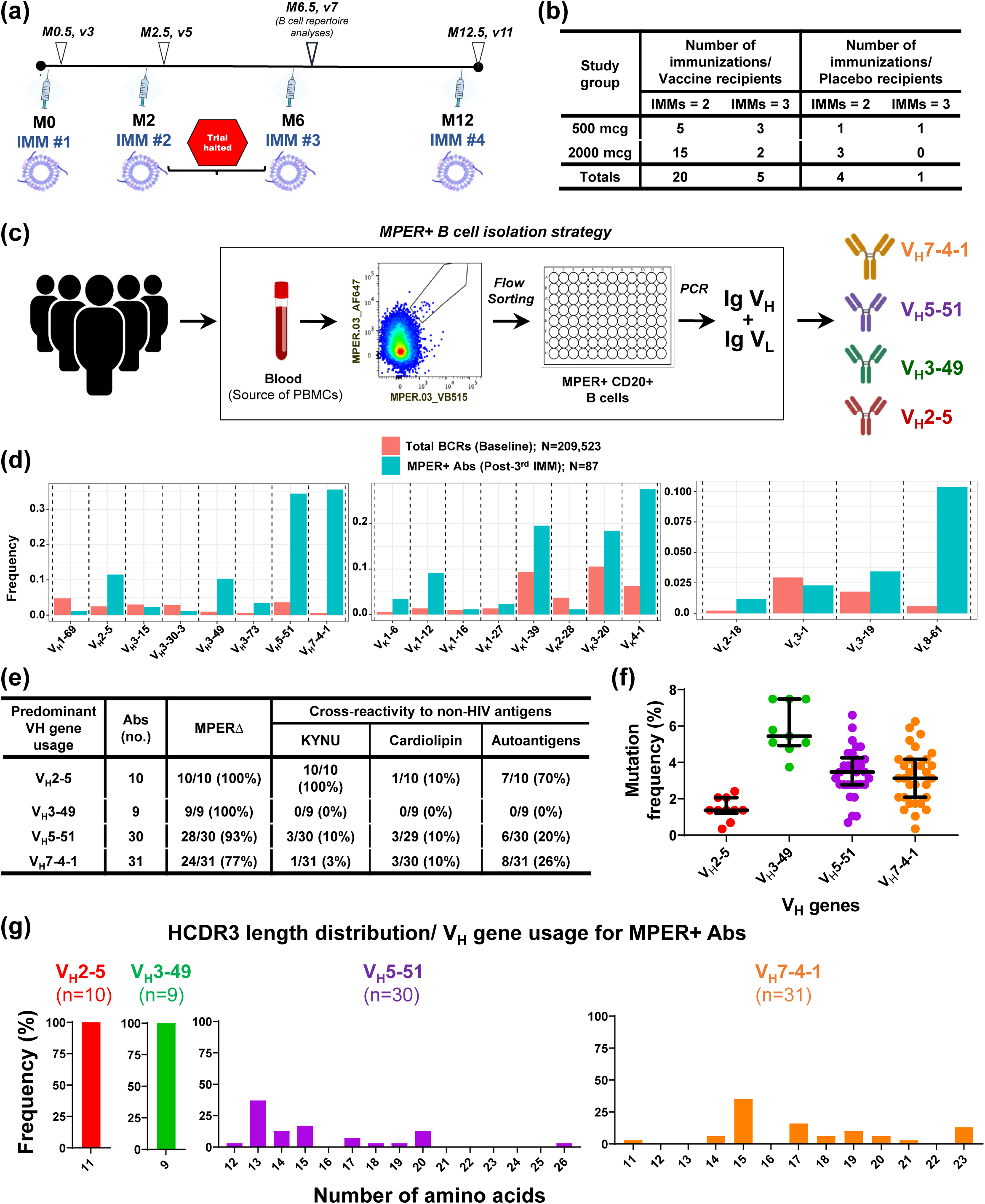
HVTN 133 Clinical Trial Elicited a Polyclonal MPER+ Antibody Response. **(a)** Immunization (IMM) schedule for HVTN 133. The timeline for IMMs in months (M) is indicated. Peripheral blood was collected from the participants at two weeks after each IMM for characterization of vaccine-induced responses. B cell repertoire analyses reported in this study was done on peripheral blood cells (PBMCs) collected 2 weeks post-3^rd^ IMM. The trial was halted after one vaccine recipient developed an anaphylactic reaction 4 hours post-3^rd^ IMM. All participants received the 2^nd^ IMM, but only five vaccine recipients received the 3^rd^ IMM. **(b)** Vaccine recipients were divided into two groups based on Alum dosing of 500 or 2000 mcg. There were four placebo recipients across both groups. **(c)** Schema for B cell repertoire analysis performed on PBMCs from 5 HVTN 133 vaccinees who received three IMMs. Boxed area: MPER+ B cells were flow sorted and Ig VH and VL genes recovered from each single sorted B cells to generate recombinant antibodies (Abs) for binding and functional screens. The top 4 V_H_ genes used by MPER+ Abs were V_H_2-5, V_H_3-49, V_H_5-51 and V_H_7-4-1, characteristic of a polyclonal response. **(d)** Frequency of immunoglobulin (Ig) genes used by MPER+ Abs isolated from blood in HVTN 133 vaccine recipients post-3^rd^ IMM. For comparison, we studied antigen-unbiased B cell receptor (BCR) repertoire pre-IMM in all five vaccinees. Data shown reflect individual Ig gene usages; unpaired heavy and light chain genes. From our reference dataset of antigen-naïve total BCR repertoires, we report only the Ig genes used by MPER+ Abs in the HVTN 133 vaccinees. The total number of Abs studied per group is listed in the figure key. Not shown: immunogenetics of two MPER+ Abs at baseline from 2/5 vaccinees studied (133-30 and 133-23). **(e)** Tally of Abs isolated post-3^rd^ immunization that were categorized based on predominant V_H_ gene used by MPER+ Abs, and frequency of MPER+ Abs tested for different binding specificities; MPERΔ bnAb precursor phenotype and cross-reactivity to non-HIV antigens (Kynu, Cardiolipin and autoantigens) – see also Tables S3 and S6. All Abs were tested for MPER binding phenotypes and Kynu-reactivity, and a majority of the Abs were tested for cross-reactivity with cardiolipin and autoantigens based on Ab availability. **(f)** Mutation frequency of the predominant V_H_ genes used by MPER+ Abs. Each dot represents a single Ab. Error bars represent the median and interquartile range (GraphPad Prism, v9.0). **(g)** CDR3 length distributions of predominant V_H_ genes used by MPER+ Abs. HCDR3 length is reported in amino acids (x-axis) and the frequency (%) of Abs with the indicated amino acid length is shown on the y-axis.

### Vaccine-induced MPER+ Antibodies

The variable heavy (V_H_) genes used in MPER+ antibodies isolated from MPER+ B cells post-3^rd^ immunization were polyclonal and predominantly V_H_7-4-1 (31/87, 36%) (V_H_ usage not previously described for MPER bnAbs); V_H_5-51 (30/87, 34%) (used by prototype proximal MPER bnAbs, m66 and m66.6) ^30^; V_H_2-5 (10/87, 11%) (used by prototype proximal MPER bnAb, 2F5) ^52^; and V_H_3-49 (9/87 (10%) (not previously described for MPER bnAbs) (**Figures 1d-1e; Table S4**). These V gene frequencies were higher than those observed in 209,523 paired IgH sequences of antigen-unbiased antibodies in pre-vaccination blood samples from the same five HVTN 133 participants (**Figure 1d; Table S5**). A diversity of light chains was also observed in post-vaccination MPER+ antibodies with the most common Vκ genes being Vκ4-1 (24/87, 28%), Vκ1-39 (17/87, 19%) and Vκ3-20 (16/87, 18%) (**Figure 1d; Table S4**). Additionally, while vaccine-induced MPER+ antibodies had higher median heavy and light chain mutation frequencies compared with antigen-unbiased antibodies in the same vaccinees prior to vaccination (**Figures 1f and S1A**), they were not as mutated as prototype MPER bnAbs (8). MPER+ antibodies using V_H_2-5 and V_H_3-49 genes had the shortest HCDR3 lengths of 11 and 9 amino acids (aa), respectively, while those using V_H_7-4-1 and V_H_5-51 genes had longer HCDR3 lengths ranging from 11 to 26 amino acids (**Figures 1g and S1b**).

Polyreactivity is a common trait of MPER bnAbs ^13, 31, 53^. MPER bnAb 2F5 cross-reacts with human protein kynureninase (KYNU) that contains a linear peptide sequence that matches identically to the core epitope gp41 sequence of 2F5, ^662^ELDKWA^667^ ^53, 54^. Of 87 MPER+ antibodies isolated after the 3^rd^ immunization, 29/87 (33%) demonstrated binding to either KYNU, clinical autoantigens and/or cardiolipin (**Table S6**). Thirteen of 16 KYNU-reactive antibodies demonstrated ≥3-fold reduction in binding to a KYNU-KO (D664E) ^54^ protein that disrupts the 2F5 epitope on KYNU (referred to as KYNU-Δ binders), and 10/13 KYNU-Δ antibodies used V_H_2-5 also found in 2F5 (**Table S6**). V_H_2-5 MPER+ antibodies were most polyreactive; 10/10 KYNU-Δ and 7/10 were reactive with either cardiolipin or clinical autoantigens (**Figure 1e; Table S6**). However, in contrast to the 22 aa length of the 2F5 bnAb, the V_H_2-5-using putative 2F5-like Abs had HCDR3s of only 11 aa (**Figure 1g**) and thus were unlikely to progress to bnAbs status ^3^.

The 2F5 UA and mature bnAb target the proximal-MPER epitope that includes ^664^DKW^666^ in SP62 peptide (^652^QQEKNEQELLELDKWASLWN^671^) ^47^. Based on alanine-substituted peptide mutation analysis, we found that the majority of MPER+ vaccine-induced antibodies (67/87, 77%) bound this MPER bnAb epitope with heterogeneous patterns of epitope reactivity within ^662^ELDKW^666^ (**Table S7**). There were no vaccine-induced antibodies isolated that bound to the distal-MPER bnAb epitope, ^671^NWFNIT^676^, although this aa sequence was present in the MPER peptide-liposome.

### HIV-1 Neutralization Activity of MPER+ Antibodies

Next, we assayed MPER+ and MPERΔ recombinant antibodies for their ability to neutralize both easy-to-neutralize tier 1 (W61D, HXB2) and difficult-to-neutralize tier 2 (JRFL, SC44, and the transmitted-founder virus WITO) clade B HIV-1 strains in both the TZM-bl/FcγRI and TZM-bl neutralization assays. **Figure 2a** outlines the maturation pathway of MPER bnAbs and the use of the TZM-bl/FcγRI and TZM-bl neutralization assays to define precursors with limited breadth in contrast to mature antibodies with bnAb status. From 87 vaccine-induced MPER+ antibodies that constituted 38 unique clonal lineages, we selected 80 mAbs representative of the 38 clones and tested them for HIV-1 neutralization in TZM-bl and TZM-bl/FcγRI cells (**Figure 2b**). Forty-nine of 80 MPER+ antibodies (61%) neutralized tier 1 and/or tier 2 HIV-1 strains in the TZM-bl/FcγR1 neutralization assay (**Table S8**). These 49 MPER+ neutralizing antibodies constituted 24 clonal lineages from either vaccinee 133-23 or 133-39 and used six different heavy chain genes (V_H_7-4-1, V_H_5-51, V_H_3-49, V_H_2-5, V_H_1-69, and V_H_3-73) (**Figures 2c, S2; Tables S8-S9**), with HCDR3 lengths ranging from 9-26 amino acids (**Figure S1c; Tables S8-S9**). Within the 14 B cell clonal lineages that neutralized tier 1 HIV-1 strains in the TZM-bl assay, two V_H_7-4-1 clonal lineages, the single member DH1425 nAb and an 11-member clonal lineage, DH1317, also neutralized heterologous HIV-1 tier 2 strains in the TZM-bl assay (**Figure 2c; Tables S8-S9**).

**Figure 2.**
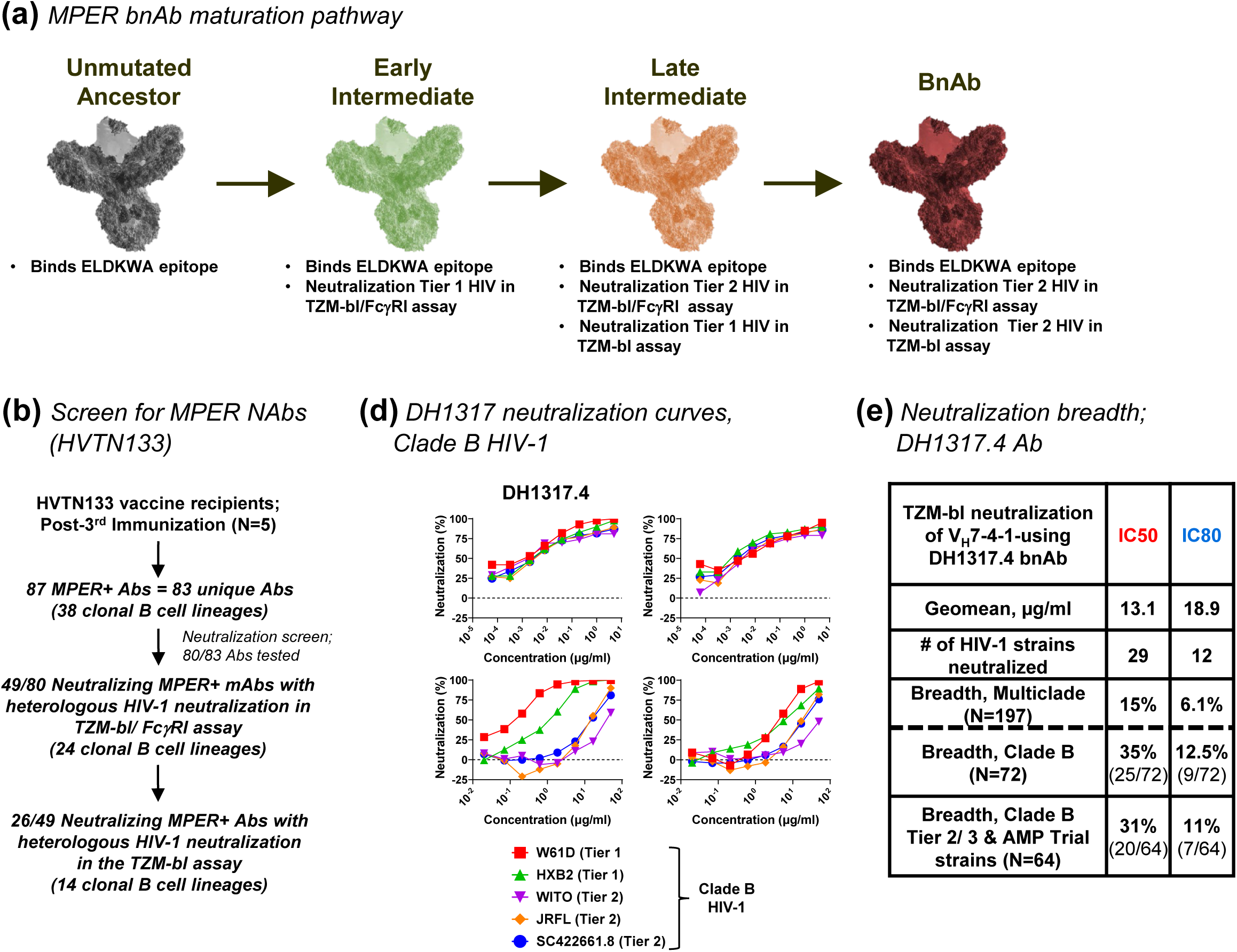
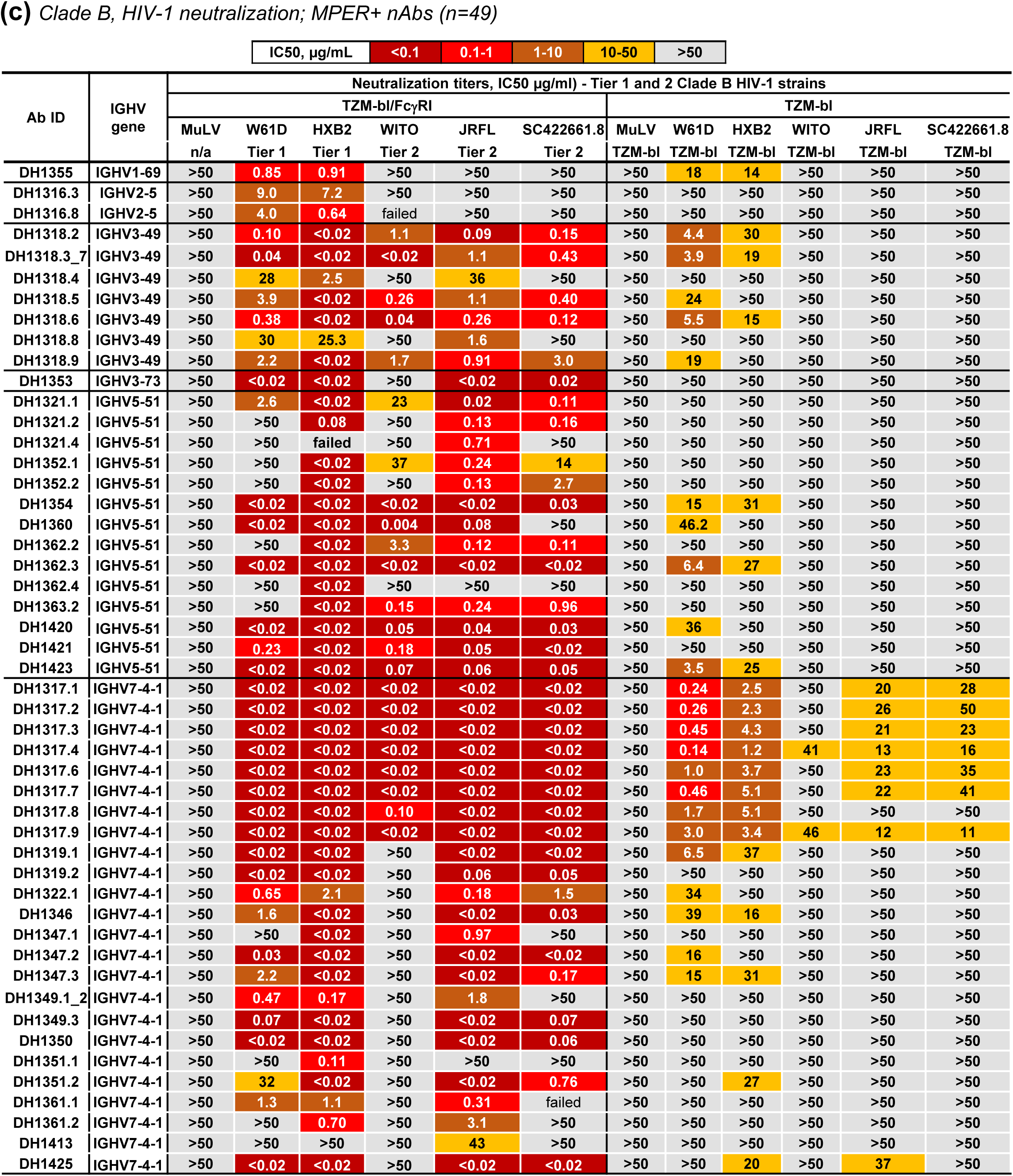
Neutralization Breadth Profile of Vaccine-induced DH1317 BnAb. **(a)** Schematic outlining maturation pathway for MPER bnAbs. **(b)** Flowchart summarizing the neutralization screens for MPER+ recombinant Abs isolated from HVTN133 vaccine recipients post-3^rd^ immunization. Eighty unique Abs representative of 83 total MPER+ Abs were tested for neutralization. Three additional MPER+ Abs were not tested for neutralization due to low Ab yields following 293i cell transfections. **(c)** Neutralization profile of 49/80 MPER+ Abs that neutralized tier 1 or 2 clade B HIV-1 strains in TZM-bl/FcγRI or TZM-bl assays. Neutralization titers were measured in IC50 (µg/ml). **(d)** Neutralization curves against tier 1 and 2 clade B HIV-1 strains for representative DH1317 lineage Abs; DH1317.4 and DH1317.9. Neutralization was performed in the TZM-bl and TZM-bl/FcgRI assays. Data shown are from single or duplicate experiments; the mean titers are shown for results from duplicate experiments. **(e)** Summary of neutralization breadth for one representative member (DH1317.4) of a V_H_7-4-1-using DH1317 clonal lineage of Abs that demonstrated neutralization of heterologous tier 1 and 2 HIV-1 strains in panel c.

### TZM-bl Assay Heterologous Tier 2 HIV-1 Neutralization Activity

V_H_7-4-1 is an uncommonly used heavy chain gene expressed at less than 2% in the B cell repertoire (**Table S5**), although it was the most common V_H_ used among the MPER+ B cell receptors (BCRs) elicited (**Figure 1d; Tables S3-S4**). Of two alleles, V_H_7-4-1*02 is the most frequently used, and occurs in ∼43% of individuals ^55^. Poor expression of the V_H_7-4-1*01 allele is due to an unpaired cysteine at heavy chain gene position 92 that compromises Ab expression ^55^. Of the 5 HVTN 133 vaccinees who received three immunizations, only the two best vaccine-responders, 133-23 and 133-39, had V_H_7-4-1 transcript frequencies >1% suggestive of these vaccine recipients having V_H_7-4-1*02 allele. Both vaccinees 133-23 and 133-39 elicited V_H_7-4-1-using tier 2 virus-neutralizing clonal lineages (**Figure S1d; Table S8**).

The V_H_7-4-1 neutralizing B cell clonal lineage DH1317 contained 11 IgG MPER+ antibodies with a 15 aa-long HCDR3 and nucleotide mutation frequencies of ∼2-6%, paired with the Vκ1-12 gene (**Figure S2a; Table S9**). Analysis of the DH1317 clonal lineage members using the SP62 MPER alanine scanning peptide set demonstrated binding dependence on the D^664^ of the ^662^ELDKWA^667^ epitope with >50% reduction in binding to D664A peptide (**Figure 3a; Tables S7 and S10**). Five of eleven DH1317 mature clonal lineage members were polyreactive for host antigens (**Table S6**).

**Figure 3.**
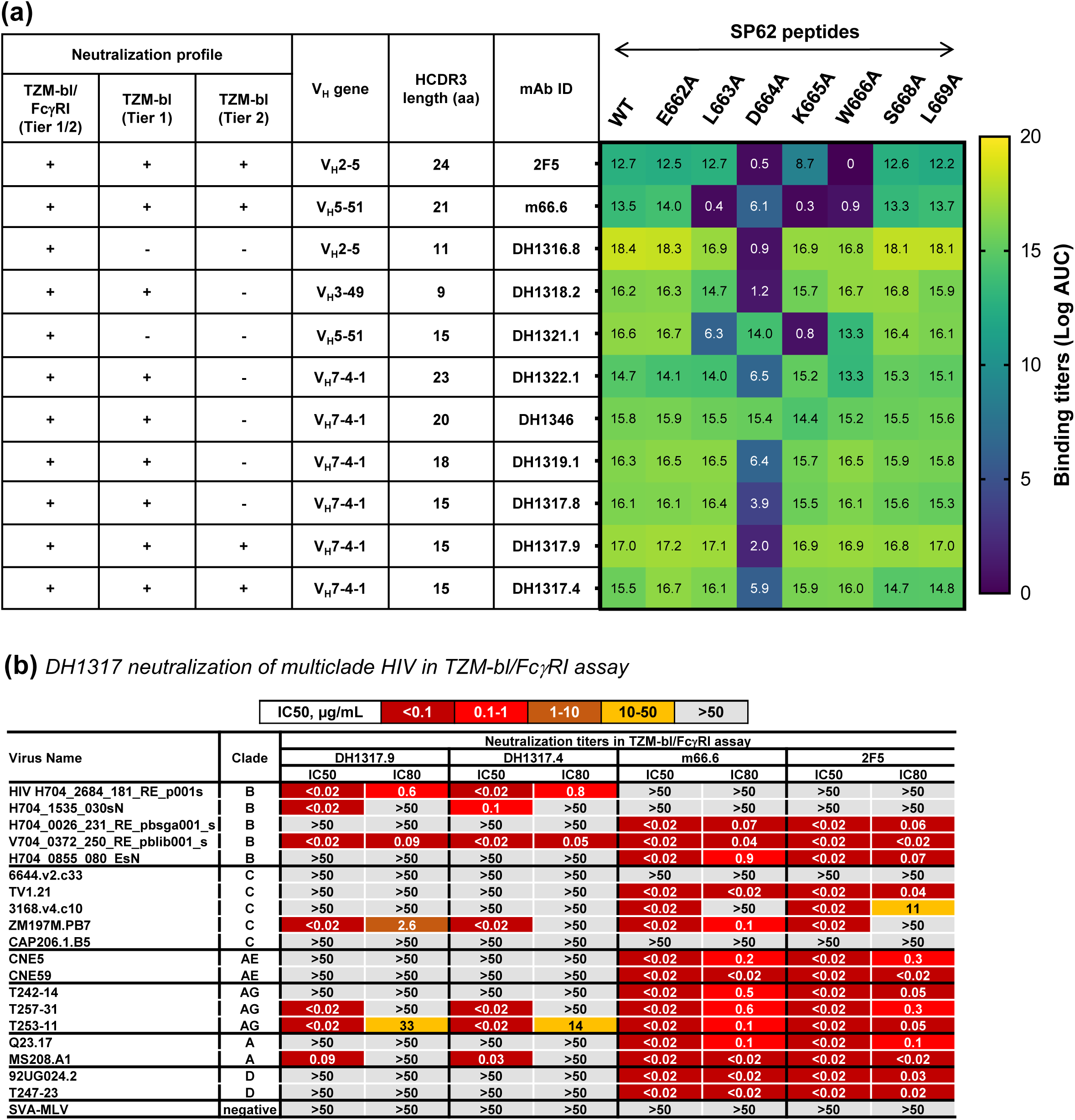
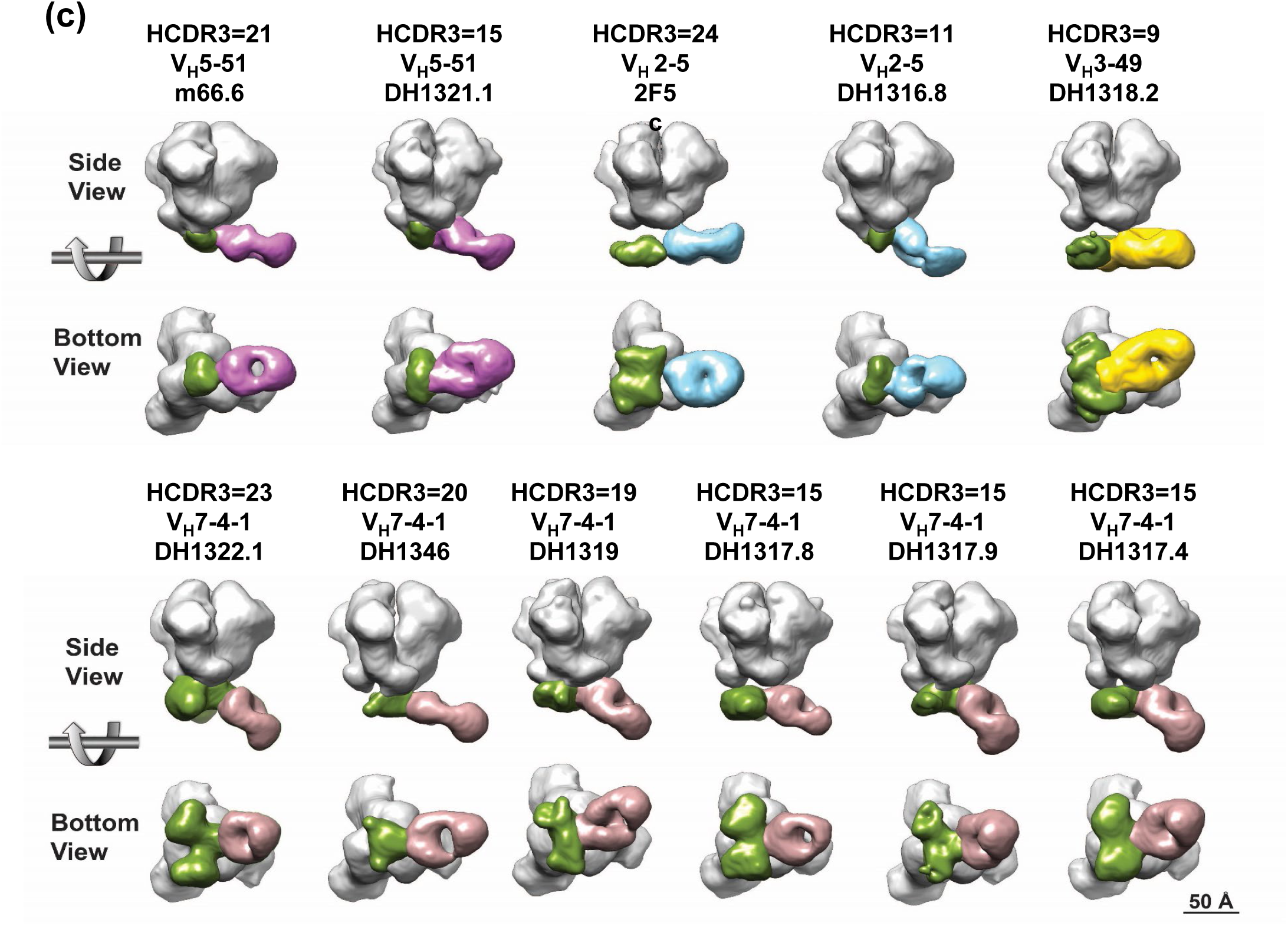
MPER+ NAbs Mapped to Proximal BnAb Epitopes. **(a)** *(Left)* A summary of neutralization profile and heavy chain gene used by the MPER+ neutralizing antibodies (nAbs). *(Right)* Mapping of MPER+ nAbs to proximal MPER bnAb epitope in SP62 peptides. Binding was measured in ELISA and reported as Log AUC as shown in the heatmap. Binding sensitivity or epitope mapping to a specific residue in SP62 tested were based on ≥50% reduction in binding to single alanine-scanned mutant peptides compared to the wild-type. **(b)** Neutralization titers of DH1317.4, DH1317.9 and MPER proximal bnAbs m66.6 and 2F5 tested against a panel of clade B HIV-1 strains from the AMP trial and non-clade B HIV-1 strains. Neutralization titers shown were measured in the TZM-bl/FcγRI assay and titers reported as IC50 and IC80 (µg/ml). DH1317.4 and DH1317.9 did not neutralize these viruses with titers <50µg/ml in the TZM-bl assay (see Figure S3). **(c)** NSEM maps for 11 MPER-directed Abs shown in side and bottom views. The Env trimer, excluding the MPER domain is colored gray, the MPER region with partial Fab density is colored green, and the MPER Fabs are colored according to their heavy chain gene as follows: V_H_5-51 = pink; V_H_2-5 = sky blue; V_H_3-49 = gold, and V_H_7-4-1 = brown.

In the standard HIV-1 pseudovirus assay in TZM-bl cells, we found that 8/11 DH1317 B cell lineage member antibodies and 5 inferred intermediate antibodies neutralized both tier 1 and 2 heterologous HIV-1 strains (**Table S11**), with neutralization potency and breadth enhanced in TZM-bl/FcγRI cells by 2 orders of magnitude (**Figures 2c-2d, 3b; Table S11**). The DH1317 UCA and 5 additional intermediate antibodies neutralized tiers 1 and 2 HIV-1 strains in TZM-bl/FcγRI cells, but not in TZM-bl cells (**Table S11**). Thus, the stages of vaccine-induced affinity maturation of proximal-MPER neutralizing antibody activity include primary acquisition of the ability to neutralize tier 1 and tier 2 HIV-1 in the TZM-bl/FcγRI assay, followed by neutralization of tier 1 HIV-1 in the standard TZM-bl assay, and then acquisition of neutralization in the standard TZM-bl assay of tier 2 HIV-1 strains (**Figure 2a**).

DH1317.4 and DH1317.9 Abs demonstrated the most potent neutralization of tier 2 clade B HIV-1 strains in TZM-bl cells (**Figure 2c; Table S11**). DH1317.4 neutralized 29/197 (15%) multiclade HIV-1 strains in the standard TZM-bl assay, including 35% of clade B HIV-1 strains (**Figures 2e and S3**). Of 125 non-clade B HIV-1 strains, DH1317.4 neutralized two clade G viruses and one recombinant clade CD virus (**Figure S3**).

To determine if lack of neutralization of non-clade B strains by antibodies DH1317.4 and DH1317.9 in the standard TZM-bl neutralization assay was due to lack of MPER binding to the antibody fragment of antigen binding (Fab) paratope or to insufficient virion lipid binding, we tested neutralizing antibodies DH1317.4 and DH1317.9 for their ability to neutralize non-clade B viruses in the TZM-bl/FcγRI assay ^32^. Both DH1317.4 and DH1317.9 neutralized 7/19 HIV-1 strains in the TZM-bl/FcγRI assay selected for their neutralization resistance in the TZM-bl assay (**Figure 3b**). The tier 2 HIV-1 strains neutralized in the TZM-bl/FcγRI assay included clades A (1/2), C (1/3), recombinant AG (2/3) and B (3/5) pseudoviruses, the latter from placebo recipients who acquired HIV-1 infection in the recent Antibody Mediated Protection (AMP) trials (NCT02716675, NCT02568215)] (**Figure 3b**). These data demonstrated that the paratope of DH1317 neutralizing antibodies was in a conformation capable of binding to the neutralizing gp41 epitope in a subset of resistant HIV-1 strains, but lacked sufficient antibody reactivity with virion lipid for TZM-bl cells to neutralize these viruses.

One additional single member V_H_7-4-1-using lineage, DH1425, was also isolated that neutralized the tier 2 HIV-1 JRFL strain in the TZM-bl assay with IC50 of 37 µg/ml (**Figure 2c**). DH1425 has 14 aa-long HCDR3 and uses a Vκ3-20 light chain (**Table S9**). These data indicated that while bnAb maturation in vaccinated humans in the HVTN 133 trial had proceeded further towards bnAb breadth and potency than in any previous study in humans, full neutralizing antibody B cell maturation after three immunizations was incomplete.

### Mechanisms of Neutralization By Vaccine-induced Antibodies

#### The effect of HIV-1 diversity

When compared to prototype bnAbs m66.6 and 2F5, DH1317.4 had similar neutralization profiles both in terms of correlation of neutralization potency as well as with overlap of commonly sensitive or resistant viruses (**Figure S4)**. An analysis of Env amino acid signatures of neutralization of 197 HIV-1 strains by DH1317.4 compared to the prototype proximal MPER bnAb, 2F5, demonstrated that DH1317.4 had similar patterns of neutralization resistance as 2F5, but that 2F5 tolerated more amino acid diversity within the MPER compared to DH1317.4 (**Figures S5a-S5d**). While 2F5 could neutralize more viruses containing amino acids ^658^K, ^659^D, ^662^A, ^667^N and ^668^N than DH1317.4, both bnAbs could not neutralize viruses with ^665^S or ^667^K (**Figure S5d**). DH1317 neutralization resistance to non-clade B strains (**Figure S5e**) could in part be explained by the inability of DH1317 to recognize MPER bnAb epitope amino acid variability since these clades were enriched for DH1317.4 resistant amino acid signatures (**Figure S5c**). For example, the most common amino acid in each of these positions in the B clade, which was most sensitive, were the amino acids which matched the immunogen; Q658, E659, E662, A667, and S668, while the most common amino acids in the C clade in these positions all differ, and were: K658, D659, A662, N667 and N668. B clade viruses were most sensitive to 2F5 and DH1317 antibodies, while C clade viruses were resistant (**Figure S5b**) ^56^. Analyzing the impact of sequence diversity at the above signature sites in the more sensitive TZM-bl/FcγRI assay, we found that while viruses with resistance mutations D659, A662 and N668 could be neutralized by DH1317.4 and DH1317.9, viruses with K658, S665 and K667 could not. Notably, all 12 viruses resistant to DH1317.4 or DH1317.9 carried the resistance signature K658, while all 7 sensitive viruses carried the sensitivity signature Q658 or N658 (p=1.9×10^-5^, Fisher’s exact test) (**Figure S5f**). Similarly, for 2F5 and m66.6, resistance in the TZM-bl/FcγRI assay was perfectly associated with absence of the sensitivity signature K665 (p=0.0003, Fisher’s exact test). These results together suggest that, like the prototypical bnAb 2F5, sequence diversity poses a roadblock for DH1317 neutralization of some major HIV-1 clades, and that some, but not all, of the underlying resistant variants can be overcome by improved lipid binding (as mimicked in the TZM-bl/FcγRI assay).

#### The effect of lipid binding

Virion lipid reactivity is a feature of MPER+ bnAbs, with a role for the CDRH3 hydrophobic loop in lipid binding and neutralization ^32^. Crystallographic analysis of 4E10 bnAb bound to lipids led to the identification of a CDRH1 binding motif (CDRH1 site 1) that interacted with lipid head groups, in addition to the utilization of CDRH3 in binding to the hydrophobic acyl chain ^33^. Examination of the CDRH1 residues in DH1317 lineage, as well as in other V_H_7-4-1 neutralizing antibody members, revealed that the putative lipid head group-binding CDRH1 sequences were similar (**Figure S6a**). However, the V_H_7-4-1 antibodies, including members of DH1317 lineage, did not bind equally to liposomes containing phosphatidylglycerol (PG) (**Figures 4a and S6b**), a lipid ligand for the MPER bnAb, 4E10 ^33^. It is notable that antibodies with weaker PG-lipid binding had negatively charged residues adjacent to the putative CDRH1 lipid head group binding site (**Figure S6a**) that potentially could impede lipid binding due to charge hindrance. For the heterologous neutralizing antibody clonal lineage, DH1317, a significant correlation was observed between PG-lipid binding and HIV-1 strain W61D neutralization potency (Kendall’s Tau correlation coefficient =0.6363, P=0.0054) (**Figure 4b**).

**Figure 4.**
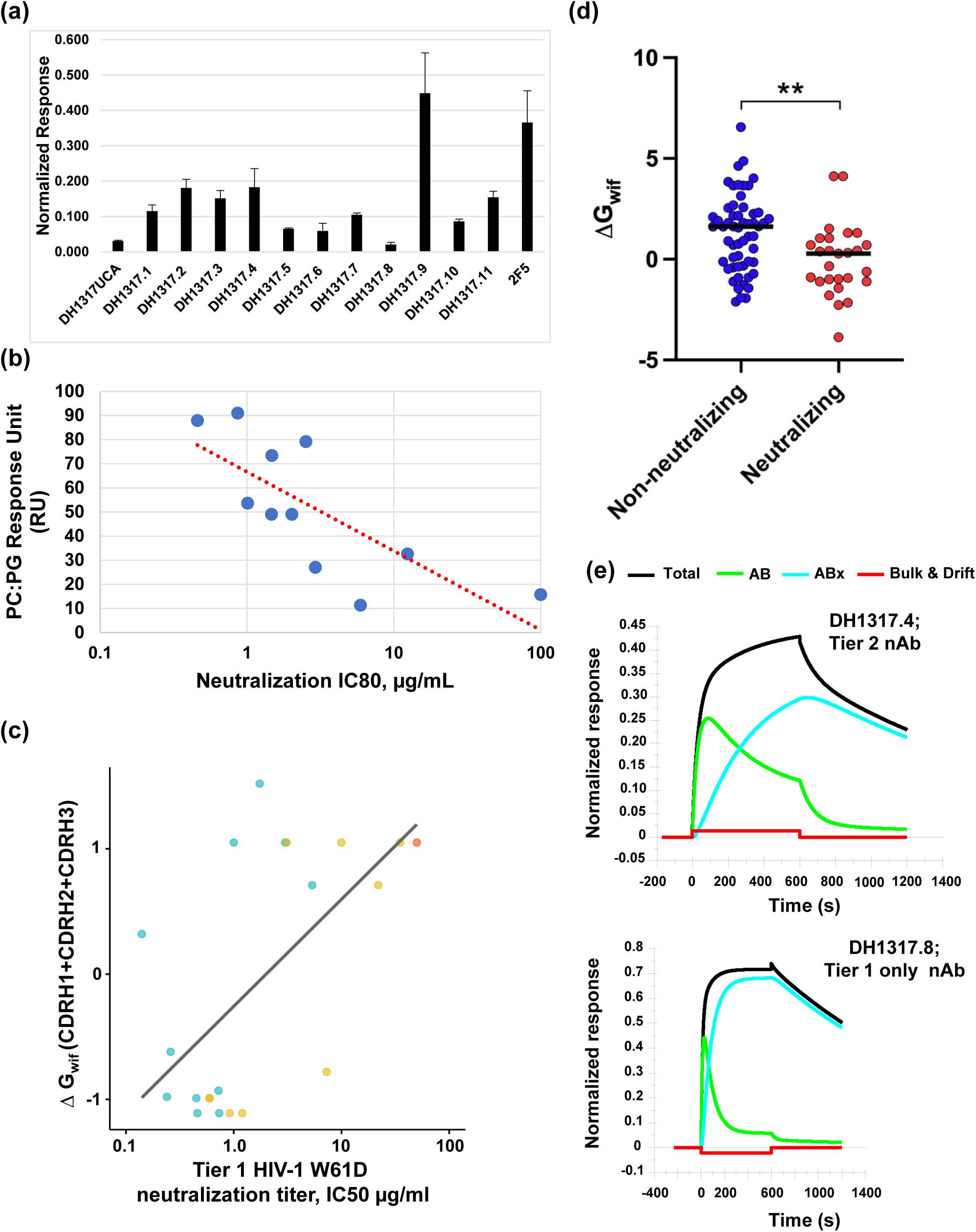
Lipid Reactivity Profiles of DH1317 Lineage Antibodies. **(a)** Binding of each indicated antibody (Ab) at 100µg/mL to POPC:DOPG (25:75) liposomes were measured by surface plasmon resonance (SPR) and plotted data show binding responses in response units (RU) normalized to control surface (lipophilic linker) and capture level of liposomes. Values and error bars represent the average and standard deviation of three measurements. **(b)** Plot of phosphatidylglycerol lipid (PC:PG) binding response in RU versus W61D pseudovirus neutralization IC80 values of each of the DH1317 lineage Abs. DH1317 UCA IC80 of >50 µg/mL is plotted with a given value of 100 µg/mL. Phosphatidylgycerol lipid containing binary liposomes were prepared using phospholipids POPC and DOPG as described in methods and binding response to DH1317 Abs were measured using SPR analysis. The plotted PC:PG binding responses are mean values of three independent experiments. Kendall’s Tau correlation coefficient =0.6363, P=0.0054 for both IC80 and IC50 values. **(c)** Computed lipid insertion propensity scores ΔG_wif_ of HCDR1, 2 and 3 (summed) versus IC50 (µg/mL) W61D neutralization in the TZM-bl assay. Computed lipid insertion propensity scores ΔG_wif_ (kcal/mol) for HCDR1, 2 and 3 (summed) is positively correlated with log W61D neutralization potency for DH1317 clonal members (R = 0.61, P = 0.0043, Pearson correlation). Lower ΔG_wif_ scores correspond to more favorable lipid insertion propensity. Isolated members of the DH1317 clone are shown in blue, inferred intermediate Abs in yellow, and inferred UCA in red. DH1317I6 and I7 appear as a single dot due to identical ΔG_wif_ scores (−0.99) and almost identical neutralization values (0.598 and 0.595, respectively). Comprehensive ΔG_wif_ scores of all antibodies within the DH1317 clonal lineage are detailed in Table S12, and corresponding neutralization values are available in Table S11. **(d)** Computed lipid insertion propensity scores ΔG_wif_ (kcal/mol) for HCDR1, 2 and 3 (summed) are significantly higher (less favorable for lipid insertion) in W61D nnAbs (blue) than nAbs (red). ** p<0.01; Mann-Whitney test. The ΔG_wif_ score is based on free energy of binding, where negative free energy is more favorable. **(e)** Biolayer interferometry (BLI) binding of DH1317.4 heterologous tier 2 nAb (top) or DH1317.8 (bottom) tier 1 only nAb Fabs to MPER liposomes fitted to the 2-step conformational change model. SPR binding curve (black) in each panel is overlayed with resolved component curves representing the encounter (first step, green) and the docked complexes (2nd step, blue).

We next computed lipid insertion propensity scores (ΔG_wif_) ^36^ of amino acids in the CDRs of each mature DH1317 lineage neutralizing antibody, as well as inferred antibody intermediates and UCA in the DH1317 lineage (**Table S12**). The ΔG_wif_ scores corresponding to more favorable lipid insertion propensity were also associated with higher HIV W61D pseudovirus neutralization potency (**Figure 4c**). We also observed more favorable ΔG_wif_ lipid insertion scores in the overall set of W61D neutralizing Abs (n=27) compared to non-neutralizing antibodies (n=56) (**Figure 4d; Table S13**). Regarding ΔG_wif_ total CDR scores, HVTN 133 induced antibodies were more similar to the prototype MPER bnAb, m66.6 than to MPER bnAb 2F5 (**Table S13**).

Each of the DH1317 neutralizing antibodies bound to MPER peptide-liposomes, and were able to recognize MPER epitopes in the context of the lipid membrane (**Figure S7**). However, the mode of binding to MPER peptide-liposomes were different when DH1317.4 that neutralized tier 2 viruses was compared to the DH1317.8 antibody that neutralized only tier 1 viruses in the TZM-bl assay. Binding of DH1317.4 to MPER liposomes could be fit to a sequential 2-step conformational change model, a binding profile previously described for MPER bnAbs that interact sequentially with lipids and MPER epitopes ^28, 31, 32, 36^, whereas the DH1317.8 could not (**Figure 4e**). Thus, the heterologous tier 2 neutralizing antibody, DH1317.4, displayed a two-step binding model similar to that previously reported for prototype proximal and distal MPER bnAbs (2F5, 4E10, DH511 and 10E8) ^28, 50^.

#### Vaccine selection of functional improbable antibody mutations

A major roadblock to bnAb development is the need for improbable bnAb lineage mutations ^10^. We computationally reconstructed the clonal tree of the DH1317 B cell lineage and identified improbable mutations (<2% probability of occurrence in germinal centers prior to antigenic selection) acquired during its maturation. We found that the MPER-peptide liposome selected for a number of improbable mutations in the DH1317 clone resulting in tier 2 heterologous HIV-1 neutralizing activity in most members of the DH1317 B cell lineage (**Figure S8**).

To assess the functional relevance of DH1317 clonal lineage mutations, we tested neutralization of mutant antibodies generated by introducing each individual mutation into the preceding intermediate (**Figure S9**). No neutralization was observed by the DH1317UCA in the TZM-bl assay prior to the second intermediate (DH1317I3) in the maturation pathway (**Figure S9; Tables 1 and S11**). We therefore measured neutralization from the UCA to the first intermediate (DH1317I4) branch using the TZM-bl/FcγRI assay (**Figure S9; Table 1**). There were two mutations going from the UCA to the first intermediate antibody; alanine to threonine at position 51 (A51T) and threonine to proline at position 99 (T99P) in the light chain, and both were estimated to be improbable. The introduction of A51T or T99P into DH1317 UCA (DH1317UCA+A51T and DH1317UCA+T99P) resulted in improvement in neutralization potency for HIV-1 W61D, HXB2 and JRFL strains compared to the UCA alone, with improvement the greatest with T99P (**Figure S9; Table 1**). The combination of both A51T and T99P (equivalent to DH1317I4) resulted in neutralization of all three strains tested in the TZM-bl assay as well as enhanced neutralization in the TZM-bl/FcγRI assay (**Figure S9; Table 1**). The emergence of the improbable mutation from tyrosine to serine at position 60 (Y60S) near the CDRH2 in the second intermediate (DH1317I3) resulted in enhanced neutralization of W61D, HXB2 and JRFL in the TZM-bl and TZM-bl FcγRI assays, thus demonstrating the critical role of Y60S for acquisition of HIV-1 neutralization breadth. While no single mutation present in the third intermediate, termed DH1317I2, (S77N, K114T, and A50G) substantially improved neutralization potency, we observed >2-fold higher potency in DH1317I2 for two out of three HIV-1 strains tested in the TZM-bl assay, indicating that smaller individual contributions of the mutations along this branch in the maturation pathway combined to increase potency (**Table 1**). Of the fourteen mutations between I2 and DH1317.4, only the heavy chain improbable mutation L72V increased neutralization potency by greater than two-fold for any of the viruses tested (**Figure S9; Table 1**). Thus, these data demonstrated that vaccination with the MPER peptide-liposome with three immunizations could select for functional improbable mutations (A51T and T99P in the light chain, and Y60S and L72V in the heavy chain) required for the development of heterologous HIV-1 neutralization by DH1317.4 bnAb.

**Table 1.** Functional Improbable Mutation Analysis in the DH1317 B Cell Clonal Lineage.

| Antibodies Tested | Neutralization Titer (IC50, µg/ml) |  |  |  | Binding Analysis |  |
| --- | --- | --- | --- | --- | --- | --- |
|  | TzM-bl |  | TzM-bl/ FcγRI |  | MPER peptide (KD, nM) | POPC-DOPG Liposome (Response units) |
|  | HXB2 | W61D | HXB2 | W61D |  |  |
| DH1317 UCA | >50 | >50 | 1.1 | >50 | 49.9 | 0.03 |
| DH1317 UCA A5IT | >50 | >50 | 1.4 | 10 | 24.8 | 0.21 |
| DH1317 UCA T99P | 29 | >50 | 0.01 | 0.02 | 6.61 | 0.06 |
| DH1317 I4 | 36 | 35 | 0.015 | 1.4 | 3.26 | 0.15 |
| DH1317 I3 | 12 | 10 | 0.006 | 0.013 | 1.12 | 0.11 |
| DH1317 I2 | 3.7 | 3.1 | 0.002 | 0.012 | 1.57 | 0.22 |
| DH1317 I2 L72V | 2.8 | 1.1 | 0.0002 | 0.0008 | 0.69 | 0.18 |
| DH1317.4 | 0.9 | 0.15 | 0.002 | 0.0018 | 0.25 | 0.18 |

Shown in the table are the results from the assessment of mutations that mediated DH1317 B cell clonal lineage maturation for heterologous HIV-1 neutralizing activity and binding kinetics. For DH1317 Ab lineage binding to phosphatidylglycerol (PG)-containing lipids (POPC-DOPG liposomes), binding of each indicated mAb at 100ug/mL to POPC:DOPG (25:75) liposomes were measured by surface plasmon resonance (SPR) and plotted data show binding responses in RU normalized to control surface (lipophilic linker) and capture level of liposomes. Values represent the average of two measurements.

Next the roles of lipid versus protein epitope binding in DH1317.4 bnAb development were determined. We found that the light chain A51T only minimally increased the binding K_D_ of the DH1317 UCA Fab to MPER.03 peptide from 49.9 nM to 24.8 nM, while it enhanced binding responses to phosphatidylglycerol (PG)-containing lipids (POPC-DOPG liposomes) by approximately an order of magnitude (**Table 1; Figures S10-S11**). In contrast, light chain T99P increased the K_D_ of binding to MPER.03 peptide by 7.6-fold (**Table 1; Figure S10**). The heavy chain mutations Y60S in I3 and the L72V in DH1317.4 improved binding to MPER peptide, but not to lipid when compared to that of A51T UCA mutant. However, each intermediate antibody bound more strongly to PG-lipid than the UCA (**Table 1; Figure S11**). Binding to immunogen MPER liposomes remained weak for the A51T mutant, while T99P mutant bound more strongly, and the binding improved in the intermediate antibodies (**Figure S12**). Thus, the predominant functional role of early light chain mutation, A51T, was for lipid binding while the roles of T99P, Y60S and L72V were for polypeptide epitope binding demonstrating the critical ability of the MPER-liposome immunogen to select for functional improbable mutations mediating the two essential recognition components shared among all MPER bnAbs: MPER peptide and viral lipid binding.

### Structural Analyses of DH1317 Neutralizing Antibodies

We next performed negative-stained electron microscopy (NSEM) on 7 neutralizing B cell clonal lineages [DH1317, DH1319, DH1322 and DH1346 (V_H_7-4-1); DH1321 (V_H_5-51); DH1318 (V_H_3-49); and DH1316 (V_H_2-5)] and 2 prototype bnAbs (m66.6 and 2F5) that mapped to residues in the MPER bnAb epitope ^662^ELDKWA^667^ (**Figures 3c, S13 and S14**). We prepared complexes with a detergent micelle-bound trimer encoding MPER (JRFL SOSIP.683) (**Figure S13)** ^57^, and the Fab fragments of the MPER-directed antibodies, and visualized the Fab-trimer complexes by NSEM (**Figure 3c)**. Both the prototypic bnAbs and HVTN 133 antibodies bound to the MPER epitope in the JRFL trimer in a similar manner. Although considerable flexibility was observed in the binding of the MPER-directed antibodies to the epitope, the DH1321.1 V_H_5-51 neutralizing antibody appeared similar to the bnAb m66.6, while the DH1318 V_H_3-49 neutralizing antibody was the most similar in orientation to the prototype bnAb 2F5. Both m66.6 and 2F5 prototype bnAbs have long HCDR3s ^30^. Similar orientations of the most potent neutralizing antibody, DH1317.4 were seen when NSEM was performed on JRFL Env with both transmembrane and cytoplasmic domain regions (**Figure S15**).

Next, we used cryo-electron microscopy (cryo-EM) to determine a higher resolution structure of the heterologous neutralizing antibody DH1317.4 in complex with a stabilized, full-length JRFL Env trimer that included the ectodomain, as well as the transmembrane and cytoplasmic domains (**Figure S16 and Table S14**). The full-length Env was purified from the cell membrane of HEK293 GNT1-cells following detergent solubilization, with VRC01 Fab and PGT145 IgG added during the purification by binding to a Protein A column, followed by on-column proteolysis to remove the PGT145 Fc and release the Env bound to VRC01 and PGT145 Fabs (see methods for details). Cryo-EM structures were determined for DH1317.4 bound to the purified JRFL Env complex. Reference free 2D classification yielded diverse views of the complex and revealed classes that showed DH1317.4 Fab moieties bound to the Env, along with VRC01 bound at the CD4 binding site and PGT145 bound to the Env trimer apex. Three distinct populations of the Env complex were identified in the cryo-EM dataset, each differed in the relative order of the bound DH1317.4 observed at each of the three binding sites. All three binding sites were occupied by DH1317.4 in the trimer populations observed. We did not observe any Env trimer populations where DH1317.4 was not bound. These results demonstrate the compatibility of DH1317.4 binding to a pre-fusion closed HIV-1 Env SOSIP trimer. Some of the bound DH1317.4 Fabs were well-ordered with the entire Fab clearly visible in the cryo-EM reconstruction, whereas others were less well ordered farther from the epitope, with some binding sites where only the epitope proximal part of the bound DH1317.4 Fab was visible. These cryo-EM reconstructions were indicative of differential mobility and flexibility between the Env ectodomain and the membrane-proximal regions, which is consistent with reports that describe tilting of the Env ectodomain in the context of the lipid membrane ^58^.

A cryo-EM reconstruction of DH1317.4 Fab bound to the HIV-1 Env trimer (also bound to VRC01 and PGT145) was resolved to 6.7 Å. The bound DH1317.4 showed a considerable interface with the transmembrane/micelle region, which appeared as a disordered fuzzy density near the trimer base, where the viral membrane would be located in a membrane-bound HIV-1 Env trimer. The approach of the bound DH1317.4 revealed proximity to the gp41 glycan N611, which would have been in the way of the approaching Fab and was displaced from its canonical position and redirected to allow binding of DH1317.4

To obtain high resolution definition of the structural basis for the epitope recognition of MPER-directed antibodies elicited in this study, we co-crystallized Fab fragments of neutralizing antibody lineages DH1317, DH1322 and DH1346 V_H_7-4-1 in complex with gp41 MPER peptides (**Figures 5 and S17; Table S15**). Structures of DH1317.8 and DH1322.1 Fabs were determined to 2.4 and 2.0 Å resolution, respectively, in complex with proximal-MPER peptides spanning gp41 residues 652-671 and 651-671, respectively. The structure of clone DH1346 was determined to 2.0 Å resolution in complex with an extended MPER peptide spanning gp41 residues 656-683 (**Figures 5-6 and S17-S18; Tables S15-S16**).

**Figure 5.**
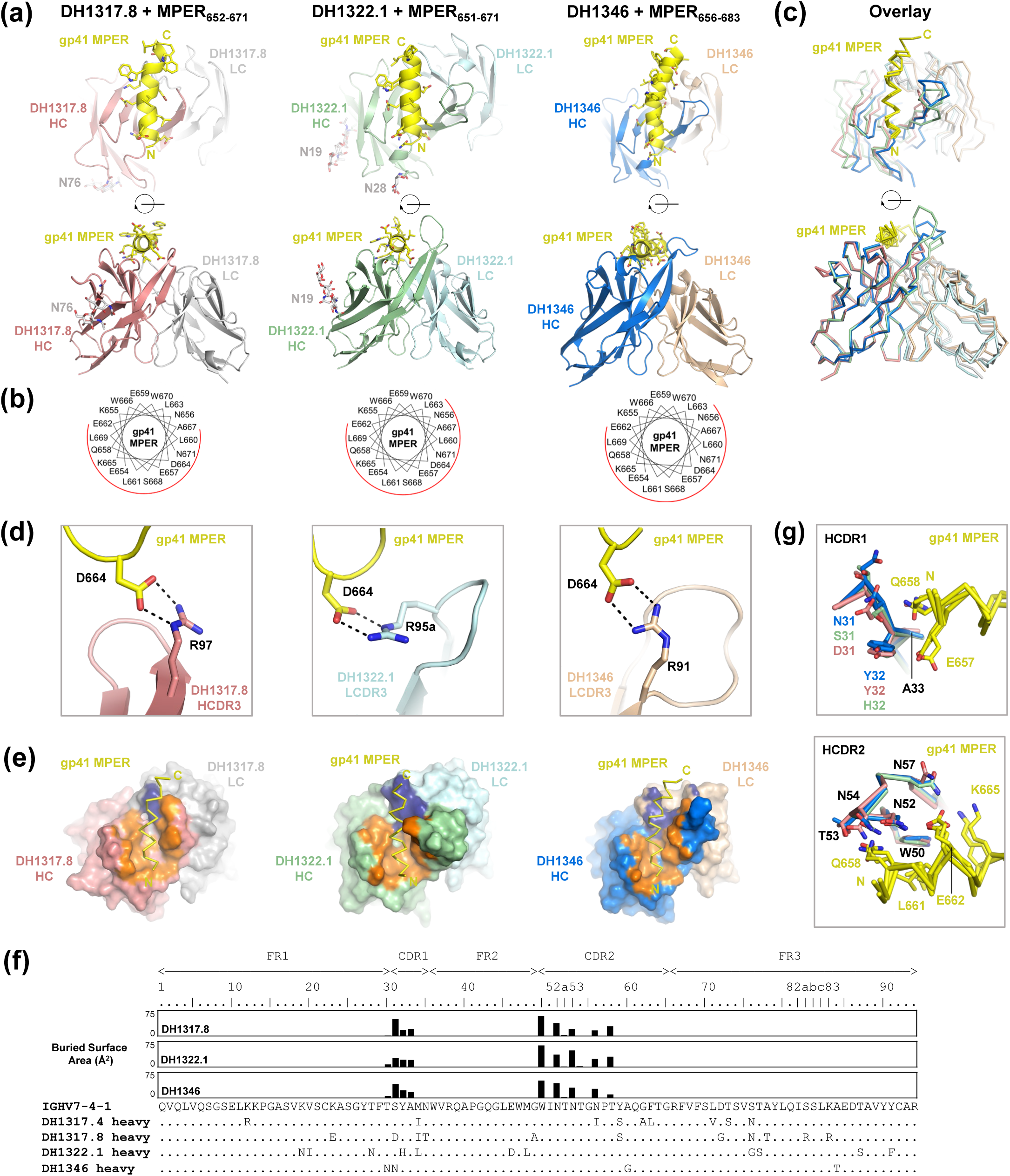
Structural Analysis of the Polyclonal V_H_7-4-1 Antibody Response to the MPER Peptide-Liposome in Vaccinee 133-23. **(a)** Structures of neutralizing antibodies (nAbs) DH1317.8, DH1322.1, and DH1346 in complex with MPER peptides, with variable regions shown in two orientations. **(b)** Helical wheel representations of MPER with nAb contacting faces shown in red. **(c).** Overlay of Ab complexes based on alignment of common V_H_7-4-1 regions, shown in same orientations and colors as in panel a. **(d)** Salt bridge interactions between MPER residue D664 and nAbs DH1317.8, DH1322.1, and DH1346. **(e)** Paratope interfaces on DH1317.8, DH1322.1, and DH1346 mapped orange on heavy chains and dark blue on light chains, shown in same orientation as in panel a (top). **(f)** Sequence alignment of the heavy chain regions of DH1317.4, DH1317.8, DH1322.1, and DH1346 against V_H_7-4-1 germline gene, with interface residue BSAs plotted as bars above. **(g)** V_H_7-4-1-encoded HCDR1 and HCDR2 loops in DH1317.8, DH1322.1, and DH1346 adopt similar conformations and interact with the same set of MPER residues, underpinning common modes of Ab recognition.

**Figure 6.**
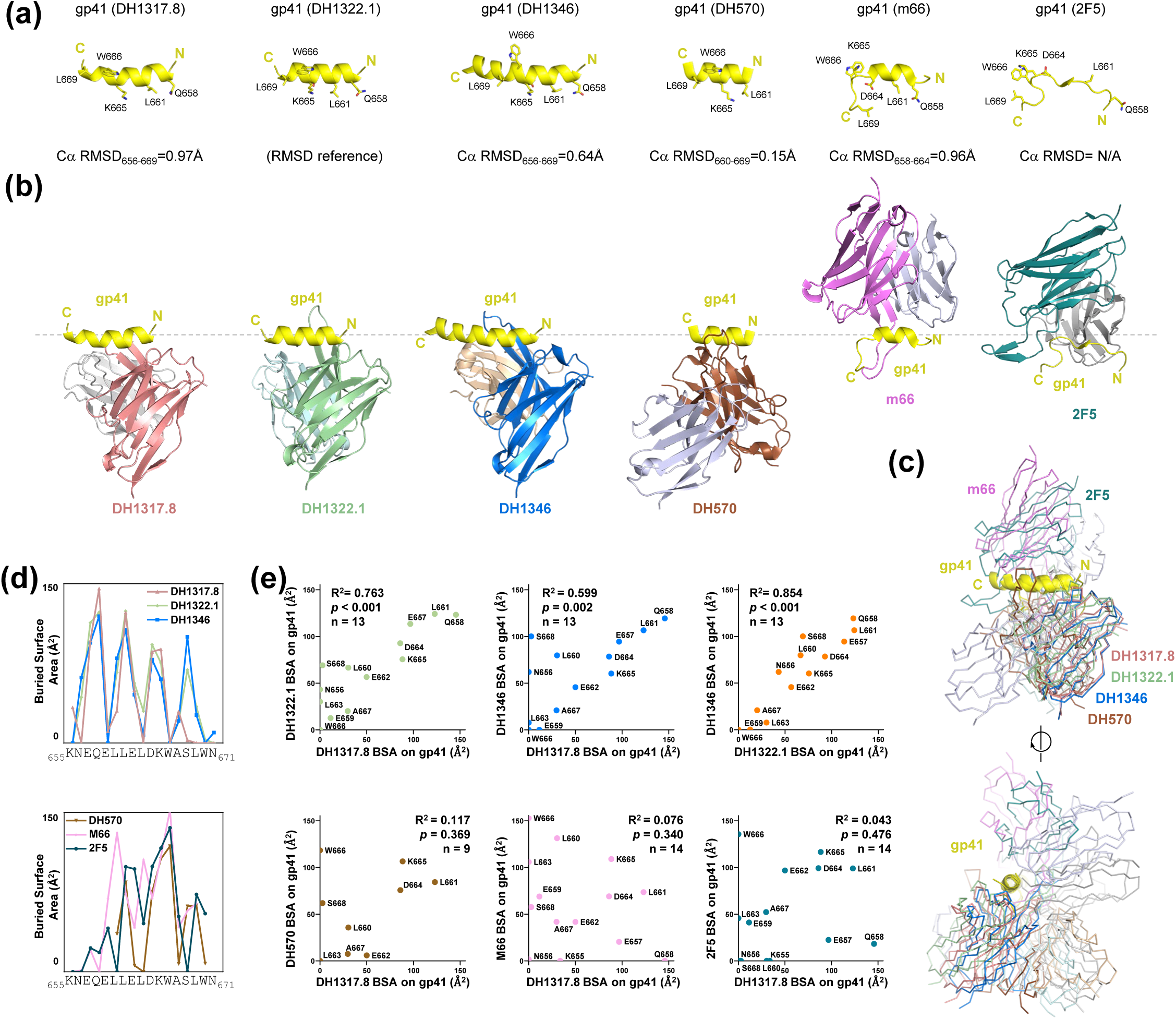
Structural Comparison of Antibody Recognition of Proximal Gp41 MPER. **(a)** DH1317.8, DH1322.1, DH1346, DH570 (5DD0), m66 (4NRX), and 2F5 (1TJI)-bound conformations of proximal gp41 MPER (yellow), oriented based on Ca alignments to reference DH1322.1-bound MPER. R.m.s.d. values shown for residue ranges used in the alignments. The epitope of 2F5 is shown based on alignment to residues 664-666 of m66-bound MPER. **(b)** Antibody (Ab) variable region structures bound to MPER, orientated based on bound MPER peptides as shown in panel a. **(c)** Overlay of MPER-bound Ab variable region structures shown in same orientations and colors as in panel b (top), and rotated ∼90° (bottom). **(d)** Plots of interface residue buried surface areas (BSA) along proximal MPER for DH1317.8, DH1322.1, and DH1346 (top) and DH570, m66, and 2F5 (bottom). **(e)** Pairwise correlation analysis of buried surface areas (BSA) of shared MPER interface residues between V_H_7-4-1-using nAbs themselves (upper panels) and with reference Abs DH570, m66, and 2F5 (lower panels).

All three V_H_7-4-1 Abs recognized a similar helical conformation of the gp41 proximal-MPER epitope (**Figures 5a, 6a and S17**). Ordered regions of the bound peptides commenced with residues N656 or E657 and extended to residues W670, N671, and K683 for DH1317.8, DH1322.1, and DH1346, respectively. In total, buried interfaces on MPER ranged from 709-865 Å^2^, while those on the antibodies ranged from 615-798 Å^2^, with the heavy and light chains accounted for 70-80% and 20-30% of the interfaces, respectively **(Figures 5f and 6d-6e; Table S16)**. Superposition of the three complexes by aligning their common V_H_7-4-1 regions revealed that the three neutralizing antibodies approached MPER from similar angles and recognized the same face of the proximal-MPER helix, one that included residue D664 (**Figure 5d**). Closer examination of D664 interactions revealed that it nestled into a pocket formed between the antibody heavy and light chains in all three neutralizing antibodies **(Figure S18a)**. D664 interactions within this pocket were anchored by a salt bridge in all three complexes, although in each case mediated by distinct antibody arginine residues: HCDR3 R97 in DH1317.8, LCDR3 R95a in DH1322.1, and LCDR3 R91 in DH1346 **(Figures 5d-5e, and S18a)**.

Since the disparate light chains and HCDR3 loops of DH1317.8, DH1322.1 and DH1346 did not alter their common modes of MPER recognition, we next assessed the contribution of their shared V_H_7-4-1 genes to epitope recognition, and found a nearly identical set of residues across the three antibodies (**Figure 5f**). These interfacial residues fell predominantly within V_H_7-4-1-encoded HCDR1 and HCDR2 loops, and exhibited similar patterns of residue buried surface areas (**Figure 5f**). While ∼4 out of 9 of these V_H_7-4-1 residue positions exhibited somatic mutations, contacts between them and gp41 were nonetheless conserved (**Figures 5f-5g**). Specifically, HCDR1 residues contacted gp41 residues E657 and Q658 in all three complexes, while HCDR2 residues commonly contacted gp41 residues Q658, L661, E662, and K665 in all three neutralizing antibodies (**Figure 5g**). At these sites, common amino acid variants Q658K, E662A, and K665S were associated with resistance to DH1317 bNAbs (**Figure S5**). Thus, the x-ray crystallographic structural analysis of a polyclonal set of V_H_7-4-1-using neutralizing antibodies revealed a novel shared mode of proximal-MPER recognition that is permissive for use of diverse light chains, and diverse heavy chain-encoded CDR3 loops.

We next compared the modes of proximal-MPER recognition of the V_H_7-4-1-using neutralizing antibodies against those of previously characterized antibodies that target this region; an MPER bnAb-precursor antibody, DH570, derived from a rhesus macaque immunized with the MPER peptide-liposome that neutralized HIV-1 in the TZM-bl/FcγRI assay (PDB 5DD0) ^36^, and the prototype proximal MPER bnAbs m66 (PDB 4NRX) and 2F5 (PDB 1TJI) (**Figure 6**) ^30, 59, 60^. DH570-bound MPER adopted an exclusively helical conformation, similar to that of V_H_7-4-1-using neutralizing antibodies, while m66-bound MPER adopted a structurally hybrid conformation that is helical between residues 658-664 but a coil otherwise (**Figure 6a**) (PDB 4NRX) ^59^. 2F5-bound MPER exclusively adopted an extended coil conformation (**Figure 6a**) (PDB 1TJI) ^60^. Superposition of the helical regions of DH570- and m66-bound epitopes to that of V_H_7-4-1 neutralizing antibody DH1322.1, which was used as a reference, revealed that DH570 approached from a similar direction as the V_H_7-4-1 using neutralizing antibodies, while m66 approached a face of the proximal-MPER helix that was opposite to the V_H_7-4-1 using neutralizing antibodies **(Figures 6b-c)**. Alignment of structurally homologous regions of bound 2F5 peptide to bound m66 peptide (namely residues 664-666) suggested that 2F5 approached the gp41 MPER from a direction similar to that of m66 when oriented in this manner (**Figures 6a-c**). Indeed, analysis of contact buried surface areas (BSA) along each MPER residue for the three V_H_7-4-1-using neutralizing antibodies as well as for prototype proximal-MPER antibodies DH570, m66, and 2F5 revealed the three V_H_7-4-1-using neutralizing antibodies mediated interactions with a nearly identical set of residues on gp41 and with similar degrees of buried surface per residue, while MPER interface residues of DH570, m66, and 2F5 exhibited a pattern of interaction distinct from that observed for the V_H_7-4-1-using neutralizing antibodies (**Figures 6d-6e**). Thus, while DH570 and m66 were similar to V_H_7-4-1 antibodies in that they recognized fully or partially helical conformations of proximal MPER, and in the case of DH570 also approached from a similar direction, the two antibodies, as well as 2F5, interacted with sets of residues on MPER that were distinct from those bound by V_H_7-4-1-using neutralizing antibodies (**Figure 6d-6e**).

Thus, our structural studies provide a comprehensive visualization across different resolution scales for how the MPER-directed antibodies that were elicited by vaccination in this study interact with their Env epitope.

### Lineage Tracing of MPER+ Neutralizing B Cells

Of the 24 vaccine-induced MPER+ neutralizing B cell clones detected post-3^rd^ immunization in the two vaccine responders 133-23 and 133-39 (**Table S9**), we found clonal relatives for 20 of these clones over time via Illumina NGS (**Tables S17-S18**). Two clones were detected as early as post-1^st^ immunization (visit 3), 11 clones were detected post-2^nd^ immunization (visit 5) and 7 clones were detected only post-3^rd^ immunization (visit 7) (**Table S18**). In vaccinee 133-23, we found a total of 66 unique VH reads that were clonally-related to DH1317 at both post 2^nd^ and 3^rd^ immunizations (**Table S18**). Of 66 DH1317-like VH reads, 45 (68%) had Y60S, and 2 (3%) had L72V after three immunizations, improbable mutations that were associated with DH1317.4 bnAb maturation (**Table S19**). These data demonstrated that vaccine-induced MPER+ bnAb B cell lineages such as DH1317 can be initiated after two immunizations.

## Discussion

Here, we have demonstrated that a vaccine containing HIV-1 gp41 MPER peptide-liposomes induced in humans a polyclonal MPER-directed B cell repertoire response made up of both heterologous neutralizing antibody B cell clones and their precursors. Each of the neutralizing B cell clones were active against heterologous tiers 1 or 2 HIV-1 strains in TZM-bl/FcγRI cells and 58% (14/24) of these neutralizing clonal lineages also were active against heterologous HIV-1 strains in the standard TZM-bl neutralization assay. In one of five individuals who received 3 immunizations before the trial was stopped due to a PEG-associated anaphylaxis reaction, the most potent B cell clonal lineage was primarily targeted to clade B with 35% of clade B viruses neutralized in the TZM-bl assay (**Figure S3**). Lack of neutralization of clade A, C, and AG HIV-1 strains (**Figure 3b**) was due to either insufficient lipid tethering or to MPER sequence diversity. Thus, in the HVTN 133 clinical trial, the induced MPER bnAb lineages were at early to late intermediate stages of MPER HIV-1 bnAb development (**Figure 2a**).

There are multiple important implications from this study. We and others have previously demonstrated in MPER UA or mature bnAb knock-in (KI) mice that full maturation of MPER bnAbs is limited by immune tolerance, both in the bone marrow and in peripheral B cell development stages ^19, 36, 38, 53^. While the distal-MPER bnAb 4E10 maturation in 4E10 KI mouse was limited by tolerance to lipids, the proximal MPER bnAb in 2F5 was limited by tolerance to the tryptophan enzyme, kynureninase ^662^ELDKWA^667^ sequence ^36, 53, 54^. That MPER antibodies that bound to lipids were induced in HVTN 133 suggests that vaccine-induced lipid-reactive antibodies are not controlled by tolerance to lipids at least to the stage of maturation of neutralizing antibody development reported here. Thus, one hypothesis is that reformulation of the MPER peptide-liposome without PEG and performance of additional boosting may increase neutralizing antibody affinity and acquisition of additional heterologous neutralization breadth and potency. Reformulation without PEG has been accomplished and clinical trial preparations are underway.

The MPER peptide-liposome was designed to target the 2F5 UA to induce 2F5-like MPER bnAbs ^47^. While 11% of MPER+ antibodies selected by the MPER peptide-liposome used the V_H_2-5 gene also used by bnAb 2F5 and some cross-reacted with kynureninase, none had HCDR3s longer than 11 aa, directly demonstrating that vaccine induced V_H_2-5, 2F5-like Abs with long HCDR3s were disfavored. In contrast, V_H_7-4-1 heterologous neutralizing antibodies were positively selected by the MPER peptide-liposome. That tier 2 virus-neutralizing V_H_7-4-1 B cell clonal lineages had a degree of lipid membrane binding that correlated with neutralization potency, raised the hypothesis that lipids may be required to be in subsequent boosting immunogens. Only a subset of individuals are predicted to make V_H_7-4-1 utilizing MPER bnAb lineages ^55^, thus it is important that the MPER peptide-liposome induced a polyclonal neutralizing antibody response utilizing multiple heavy chain genes.

The field of HIV-1 vaccine development is working with several UCA-targeting immunogens to induce putative precursors of bnAbs ^41, 44, 61, 62^ in order to use sequential boosting immunogens ^15^ to select for improbable mutations ^10^ and induce full bnAb maturation. However, since only putative precursors have been induced thus far in these cases, it will not be certain such precursors have the capability to mature to potent and broad heterologous neutralization capacity until this has been accomplished in humans. In contrast, here we demonstrate that the putative MPER bnAb precursors in the DH1317 B cell clonal lineage that neutralized only in the TZM-bl/FcγRI assay, underwent affinity maturation to neutralize tier 2 heterologous HIV-1 in the TZM-bl assay, thus proving their bnAb precursor status (**Figure 2a**).

### Implications for future vaccine design

This study has important implications for future vaccine design. First, it has provided insights into the sequence of events that transpire when MPER bnAbs develop. Second, given the concern that MPER bnAbs are autoreactive and have long HCDR3 regions and thus will be disfavored for their induction and lineage maturation, it was encouraging to observe that while very autoreactive antibodies with long HCDR3 regions were not induced, other heterologous neutralizing B cell lineages were readily induced. Third, it has provided templates upon which to design new immunogens to boost such MPER bnAbs once induced, since the induced bnAbs in this study, while neutralizing tier 2 HIV-1 strains, do so with less breadth and potency than that will be needed by a successful HIV-1 vaccine. Thus, work is underway to design mRNAs that encode an MPER immunogen that can bind to the DH1317 lineage members and thus are likely to boost such memory B cells. Fourth, this study demonstrated the clinical limitations of the use of PEG to stabilize MPER peptide-liposomes and has stimulated the development of PEG-less immunogen versions that will be used to prime B cell responses in a new clinical trial. Fifth, prototype bnAbs 2F5, m66.6, and the antibodies induced in HVTN 133 shared breadth limitations and clade specificity, because of the distinctive variability of this epitope in different clades. The Env resistance signatures identified in this study provide a roadmap for design of boost immunogens that can improve upon the breadth of currently induced responses by exposing such resistance signatures in sequential immunogens. Alternatively, separate lineages of proximal MPER targeting antibodies may need to be induced tailored to the very distinctive form of the epitope in the C clade that dominates much of the HIV-1 pandemic, or this proximal MPER region epitope can be used in conjunction with strategies that also induce antibodies to the distal MPER region epitope that confers more global breadth. Finally, a major roadblock to induction of mature bnAbs is the length of time it takes for a HIV-1 induced bnAb B cell lineage to acquire the functional improbable mutations required to achieve bnAb breadth in the setting of HIV-1 infection. Here, we have demonstrated that a vaccine can indeed select for functional improbable mutations in bnAb B cell lineages and do so after only 2-3 immunizations. Thus, this study has provided important insights into the design and feasibility of HIV-1 vaccine development efforts to induce bnAbs.

### Limitations of the study

This study was limited by the interruption of the HVTN 133 trial and thus by the limited number of trial participants who received three immunizations. Secondly, only 2 of the 5 vaccinees were high responders to the vaccine, and they both had the VH7-4-1*02 allele that allowed them to make VH7-4-1 neutralizing antibody B cell lineages. Thus, it appears that not every vaccinee will be able to make a VH7-4-1 MPER bnAb response. Another potential limitation of the HVTN 133 clinical trial was that in a companion paper, we only observed weak serum neutralizing antibody responses that required affinity purification of the MPER+ antibodies ^51^. However, this is not necessarily a negative finding, given the necessity of keeping Env-stimulated memory B cells in germinal centers for long periods of time for accumulation of levels of mutations required for neutralizing antibody breadth and potency, rather than stimulating B cell end stage differentiation to plasma cells with secretion of high levels of serum antibody.

In summary, HIV-1 envelope-reactive B cell lineages with heterologous neutralizing breadth have been induced in humans by an MPER-targeted HIV-1 immunogen. While most of the induced neutralizing antibodies did not broadly neutralize tier 2 HIV-1 strains, some antibodies unequivocally neutralized tier 2 heterologous HIV-1, thus proving that such antibodies can indeed be induced in humans by vaccination. Moreover, these data suggested that a similar lipid-gp41 complex may be designed to complete bnAb maturation and provides a pathway for iterative development of HIV-1 vaccine immunogens that can induce broad and potent MPER bnAbs.

## Acknowledgments

Supported by the HHS, NIH, NIAID, Division of AIDS, Center for HIV/AIDS Vaccine Immunology-Immunogen Discovery (CHAVI-ID) grant AI100645, by the HHS, NIH, NIAID, Division of AIDS Consortia for HIV/AIDS Vaccine Discovery (CHAVD) grant AI144371, by the HHS, NIH, NIAID, Center for HIV Structural Biology grant U54 AI170752, and by a Collaboration for AIDS Vaccine Development (CAVD) grant, OPP1094352/INV-007688, from the Bill & Melinda Gates Foundation. We thank the DHVI program and finance teams for grant management, including Kelly Cuttle, Jordan Cocchiaro, Daniel Tonkin and Whitney Edwards-Beck. We would like to thank Ken Cronin for technical support with Fab preparation, SPR epitope mapping and affinity measurements; Jongln Hwang, Aria Arus-Altuz, Xiaozhi Lu, and Dapeng Li for technical support with flow cytometry B cell isolation and PCR amplification of Abs from sorted B cells; Sommer Holmes for technical support on recombinant Ab expression; and Ryan Tuck for support with making figures and tables. BCR repertoire sequencing (10X Genomics) was done by the DHVI Viral Sequencing Analysis Core facility, and Duke University Genomic and Computational Biology Core Facility. X-ray crystallography data were collected at Southeast Regional Collaborative Access Team (SER-CAT) 22-ID beamline at the Advanced Photon Source, Argonne National Laboratory. Use of the Advanced Photon Source was supported by the U.S. Department of Energy, Office of Science, Office of Basic Energy Sciences, under Contract No. W-31-109-Eng-38. Use of the Stanford Synchrotron Radiation Lightsource, SLAC National Accelerator Laboratory, was supported by the U.S. Department of Energy, Office of Science, Office of Basic Energy Sciences under Contract No. DE-AC02-76SF00515.

## Author Contributions

BFH conceived study, designed immunogen, reviewed all data, and wrote the paper; SMA designed immunogen, performed experiments, and edited the paper; WW oversaw isolation of Abs and BCR sequencing and repertoire analysis, collated data from across different labs, and wrote the paper; GO oversaw crystallography structural analysis; PA oversaw cryo-EM structural analyses and Env protein production for NSEM and cryo-EM; NE, SRW, and LRB were PIs of HVTN 133; DM, MS, and AE performed neutralization assays; RJE and KM performed NSEM; DWC and MM performed assays for B cell isolation; IM, TE and YC performed assays for BCR sequencing; RP, MB, and AF performed serum experiments and/or characterized Abs for binding and epitope mapping; KA performed immunogen antigenicity testing, preparation of phospholipid containing liposomes, and SPR/BLI measurements and analysis of Ab binding to lipids and to MPER liposomes; PP prepared the Fabs and performed the affinity measurements curve fitting analyses. SS, XH, JL and SF produced Envs for NSEM and cryoEM analysis; RP and KJ performed specimen optimization and screening, and collected cryo-EM data; AN, BMJ, and AA performed x-ray crystallography; MB, HK, EV and KW performed computational analysis; KOS oversaw recombinant protein expression; CBF provided PEGylated liposome for vaccine immunogen; KWC, MJE and LC oversaw the clinical trial and lab analyses by the HVTN.

## STAR Methods

### Vaccine Participants

In the HVTN 133 clinical trial (NCT03934541) twenty-four participants, including 20 vaccine and 4 placebo recipients, were enrolled and scheduled to receive four immunizations of the MPER-656 peptide/liposome immunogen. Vaccinees received either a low (group T1, 500 mcg) or high (group T2, 2000 mcg) MPER peptide-liposome dose regimen (**Figures 1A-1B**). Before completion of the scheduled vaccinations, however, the study was halted due to safety concerns. 20 participants received 2 MPER peptide liposome doses, and 5 received 5 MPER peptide-liposome doses prior to trial being halted. Informed consent was obtained by all participants and the trial was approved by the Patient Review Safety Committee at NIAID.

#### Ethical oversight

All ethical oversight and monitoring for implementation/running of the trial itself was through HVTN and Sponsor (DAIDS) standard operating procedures. The protocol was reviewed and approved by the institutional review boards at all sites (Brigham and Women’s Hospital, Fenway, Columbia University, New York Blood Center, University of Alabama Birmingham, and Fred Hutchinson Cancer Research Center) and written informed consent was obtained from each participant prior to enrollment. When the trial was halted, the event was reviewed by the Protocol Safety Review Team (PSRT)—which includes the investigators (protocol Chairs), protocol leadership, and clinical safety specialists, as well as by the HVTN Data and Safety Monitoring Board (DSMB).

#### Sample/patient identifiers

The numbers provided in the publication are not sample or patient IDs. They are specifically HVTN-provided “PubID” (“Publication ID”) numbers-generated by HVTN as an extra layer of coding, beyond the already coded PTID #s. They are intended for use in presentations and publications.

### Flow Cytometry Sorting of MPER+ B Cells

#### B Cell Sorting

Sorts of MPER-specific memory B cells were conducted as previously described ^36, 63^. Briefly, cryopreserved PBMCs were thawed and counted. Cells were labeled with optimized concentrations of the following fluorochrome-mAb conjugates: PerCP-Cy5.5 anti-human IgM (clone G20-127, BD Biosciences), PE anti-human IgD (clone IA6-2, BD Biosciences), PE-CF594 anti-human CD10 (clone HI10A, BD Biosciences), PE-Cy5 anti-human CD3 (HIT3a, BD Biosciences), PE-Cy5 anti-human CD235a (clone GA-R2, BD Biosciences), PE-Cy7 anti-human CD27 (clone O323, ThermoFisher), Alexa Fluor 700 anti-human CD38 (clone HB7, BD Biosciences), APC-Cy7 anti-human CD19 (clone SJ25C1, BD Biosciences), BV570 anti-human CD16 (clone 3G8, Biolegend), and BV605 anti-human CD4 (clone M5E2, Biolegend). Cells were also labeled with fluorochrome-labeled antigen baits, prepared as 4:1 molar mixtures of biotinylated MPR.03 peptide and Streptavidin-VioBright 515 (Miltenyi Biotec) or Streptavidin-Alexa Fluor 647 (ThermoFisher). Labeling with LIVE/DEAD Aqua (ThermoFisher) allowed identification and exclusion of dead cells. The gating strategy identify to memory B cells was: Aqua^-^CD14^-^CD16^-^CD3^-^CD235a^-^CD19^+^IgD^-^. Memory B cells that bound both MPR.03-VB515 and MPR.03-AF647 baits were sorted as single cells into wells of a 96-well plate containing PCR lysis buffer. Cells were sorted on a FACSAria IIu (BD Biosciences) using FACSDiva software version 8. Data were analyzed using FlowJo version 10 (BD Biosciences).

#### Fluorophor-conjugated MPER-peptide production

In order to produce enough conjugated peptide to stain at least 10 aliquots of 5 million cells each with excess for quality testing, 200μL of 0.2mg/mL biotinylated MPER.03 peptide (KKK^656^NEQELLELDKWASLWNWFDITNWLWYIRK^684^KK) (MW = 4678 Da) was measured out and divided into two parts (1.07nmol or 100μL each) to be conjugated to either VB515 streptavidin fluorophore (MW = 61,040 Da, commercially obtained from Millitenyi Biotec Cat#: 130-108-993) or AF647 streptavidin fluorophore (MW = 59,485 Da, commercially obtained from ThermoFisher Cat#: S21374) at a 4:1 molar ratio. To achieve a 4:1-part molar ratio of peptide to fluorophore, 53.5pmol (16.3μL) per addition of VB515 was measured for a total of 265pmol (81.5μL) and 53.5pmol (12.7μL) per addition of AF647 for a total of 265pmol (63.6μL). In an amber Eppendorf tube, the biotinylated MPER.03 peptide was incubated for 15 minutes on a continuous mixing device at room temperature with one addition of fluorophore. After the incubation, subsequent additions of fluorophore were added with 15-minute continuous mixing in between until all 5 additions were added. The conjugated peptide mixture was then diluted using 1X PBS +0.02% NaN_3_ to a concentration of 3000nM. Similarly, the biotinylated MPER.03_D664AW672A mutant peptide was conjugated to the BV421 streptavidin fluorophore (MW = 133,400 Da, commercially obtained from BioLegend, Cat#: 405225). The amount of BV421 used was 73.8μL split into 5 additions of 14.8μL. The three conjugated peptides were stored at 4°C. To confirm usability for flow studies, the peptides were tested for binding to antibody-coated beads against mixes of 20% positive beads (2F5, 13H11) and 80% negative beads (CH65). The conjugated peptides were serially diluted, mixed with the bead mixtures, incubated for 30 minutes at 4°C, gently aspirated and washed a total of 3 times using 1X PBS +0.02%NaN_3_ and assessed for binding via flow cytometry.

### PCR Isolation of Antibodies from Antigen-specific B Cell Sorts

Single sorted B cells were obtained from HVTN133 vaccine recipients using previously described flow cytometry-based sorts ^36, 63^. Cells were individually sorted into 96 well plates containing lysis buffer and immediately stored at −80. Human V_H_DJ_H_ and V_L_J_L_ segments were isolated by single-cell PCR ^22, 63, 64^. Antibody sequences were analyzed using a custom-built bioinformatics pipeline for base-calling, contig assembly, quality trimming, immunogenetic annotation with Cloanalyst (https://www.bu.edu/computationalimmunology/research/software/), VDJ sequence quality filtering, functionality assessment, and isotyping as described ^65^. The sequences of the heavy and light chain variable regions for the MPER+ antibodies isolated from antigen-specific B cell sorts were provided in **Table S4**.

### Recombinant Antibody Production

Following amplification of antibody heavy and light chain genes per B cell in our high-throughput flow cytometry sorts, we used an aliquot of the purified heavy and light chain PCR amplicon per B cell to generate a linear expression cassette bearing each amplicon as described^22, 63^. The expression cassettes were transfected into Expi293F cells (ThermoFisher, Cat No. A14527) with Expifectamine (ThermoFisher, Cat No. A14526) to generate microgram quantities of IgG. Three days after transfection, cell culture media was cleared of cells and secreted recombinant antibodies in the cell culture media were tested for binding to HIV-1 envelope via ELISA. In this initial binding screen, our positivity cutoff was OD_450_≥0.5. From the ELISA binding screen, antibodies were selected for gene synthesis in milligram quantities.

Recombinant antibodies were generated in large quantities by using commercially-obtained (GeneScript, Piscataway, NJ) plasmids with antibody heavy and light chain genes to transfect Expi 293i cells using ExpiFectamine 293 transfection reagents (Life Technologies, GIBCO; Cat#A14524) as described ^11^. Briefly, the synthesized heavy and light chain genes were cloned into gamma, kappa, or lambda expression vectors (GenScript). Plasmids were prepared for transient transfection of Expi293F cells using the MididPrep or MegaPrep plasmid plus kit (Qiagen). The purified recombinant antibodies were stored at 4°C in a citrate buffer (pH = 6.0). All recombinant antibodies were expressed from plasmids containing a human or macaque IgG constant region and were QC’ed in Western Blot and Coomassie SDS PAGE for appropriate heavy and/or light chain protein expression.

From the initial binding screen, we identified 87 MPER+ antibodies. We expressed 83 recombinant mAb that were representative of the 87 MPER+ antibodies, given that we expressed a single mAb where two antibodies had identical amino acid sequences.

### Site-Directed Mutagenesis

Fifty nanograms of antibody heavy or light chain plasmid was mutated using the QuikChange Lightning Multi Site-Directed Mutagenesis Kit (Agilent). DNA oligonucleotide primers introducing site-specific mutations were designed with the QuikChange Primer Design Tool (Agilent) and synthesized and purified via standard desalting by Integrated DNA Technologies. The manufacturer’s protocol was followed to mutate each plasmid except DpnI digestion was conducted for 1 hour. XL-10 Gold cells were transformed per the manufacturer’s protocol with 1 microliter of the mutagenesis reaction. Mutated plasmids were purified by MidiPrep (Qiagen) and Sanger sequenced to confirm introduction of mutations. Purified plasmids were used for transient transfection of 100 mL of Expi293F cells (ThermoFisher) using the Expifectamine transfection system.

### VDJ Sequencing of Single B Cells from Peripheral Blood

#### VDJ library construction and sequencing

Single Cell VDJ library construction was processed on 10X Chromium Controller (PN-1000204) using the Chromium Next GEM Single Cell 5l Kit v2, (PN-1000263) and BCR Amplification Kit (PN-1000253) following the manufacturer’s protocols. In brief, between 16,000 and 18,000 sorted B cells were loaded onto the Chromium controller to target the recovery ∼10,000 cells for each sample. Final libraries were quantified using a TapeStation 2200 (Agilent) and Qubit 3 Fluorometer (Thermo Fisher). For BCR repertoire sequencing at baseline in HVTN133 vaccine recipients, we performed negative B cell enrichment from ∼30M PBMCs in each experiment using a commercially available B cell enrichment kit according to manufacturer’s protocol (StemCell Technologies; Vancouver, BC, Canada). The VDJ libraries were sequenced on the Illumina NovaSeq 6000 platform with Novaseq S4 Reagent kit (200 cycle kit) using the read parameters: 26 cycles for Read1, 10 cycles for i7 index, 10 cycles for i5 index, and 150 cycles for Read 2. We targeted 5,000 reads/cell for the VDJ libraries.

#### VDJ analysis

Illumina NGS generated BCR sequencing data was processed using the Cell Ranger single cell gene expression software provided by 10X Genomics. Reads were demultiplexed and quality filtered, followed by assembly and annotation using Cell Ranger 6.0.0 with the GRCh38 human V(D)J gene segment reference provided by 10X Genomics. Contigs assembled by Cell Ranger were re-annotated and analyzed using Cloanalyst (https://www.bu.edu/computationalimmunology/research/software/) with the default human Ig library to determine B cell immunogenetics and clonality as previously described ^65^.

The softwares used for computational analysis of B cell receptor sequencing includes MPEX version v3.3.0, ANARCI (https://github.com/oxpig/ANARCI), Cloanalyst version 1.0.6419.16890, Figtree version 1.4.4, ARMADiLLO (https://github.com/WieheLab/ARMADiLLO) and Cell Ranger version 6.0.0. Custom script used in this manuscript for single B cell VDJ sequencing analysis has been deposited to GitHub (https://github.com/WieheLab/HVTN133).

### Next Generation Deep-Sequencing and Analysis of Antibody Genes

Illumina NextSeq sequencing of antibody heavy chain VDJ sequences was performed on peripheral blood cells using the commercially available ArcherDX BCR Repertoire kit (ARCHER, Boulder, Colorado). Removal of duplicated sequences and error correction were initially performed via Archer Analytical pipeline (https://analysis.archerdx.com/). Subsequently, paired-end sequences were merged using FLASH ^66^, quality filtered (Q score ≥30 for ≥95% of sequence) and deduplicated. V, D, and J gene segment, clonal relatedness testing and reconstruction of clonal lineage trees were performed using the Cloanalyst software package ^67^. Immunogenetics information of human antibody sequences were assigned by Cloanalyst using Cloanalyst’s default libraries of human immunoglobulin genes (https://www.bu.edu/computationalimmunology/research/software/). B cell clonality was determined based on similar heavy chain rearrangements and CDR3 length as described ^68^.

### Indirect Binding ELISA

Antibody binding was measured by ELISA in 384 well ELISA plates (Costar #3700) coated with 2mcg/ml streptavidin (Thermo Fisher Scientific Inc., Cat. No. S-888) or protein antigen in 0.1M sodium bicarbonate overnight at 4C. Plates were washed with PBS/0.1% Tween-20 and blocked for one hour with assay diluent (PBS containing 4%(w/v) whey protein/15% Normal Goat Serum/0.5% Tween-20/ 0.05% Sodium Azide). Streptavidin coated plates were washed and followed by 10ul biotinylated peptide at 2mcg/ml in assay diluent for one hour. All plates were then washed and antibodies added in 10µl volumes for 1 hour in three-fold serial dilutions beginning at 100mcg/ml. Plates were washed and 10µl HRP conjugated goat anti-human secondary Ab (Jackson ImmunoResearch C:109-035-008) diluted to 1:15,000 in assay diluent without azide was incubated at for 1 hour, washed again and detected with 20µl SureBlue Reserve (Seracare 5120-0081) for 15 minutes. The antibody binding reactions were stopped with the addition of 20µl HCL stop solution and plates read at 450nm. ELISA binding data were collected by Spectramax Plus384 plate reader, analyzed using Softmax Pro version 5.3, and collated or graphed using excel and GraphPad Prism version 9.

### Polyreactivity Binding Assays

#### AtheNA

MAb reactivity to nine autoantigens was measured using the AtheNA Multi-Lyte ANA kit (Zeus scientific, Inc, #A21101). Abs were serially diluted for four 2-fold steps starting at 300µg/mL. The Abs were then diluted 1:6 in assay beads resulting in a final serial dilution of four 2-fold steps starting at 50 µg/mL in 50ul of beads. The kit SOP was followed for the remainder of the assay. Samples were analyzed using AtheNA software. An individual well was positive if >120 AU (Athena Units), negative if <100 and considered indeterminate if between 100-120. Any given Ab needed to be positive for two consecutive wells (i.e. to at least 25µg/ml) to be considered positive for autoreactivity; see values with yellow fill in **Table S6**.

#### Anti-cardiolipin reactivity

MAb reactivity to cardiolipin was measured using the Quanta Lite ACA IgG III kit (Inova Diagnostics, Inc.) by diluting Abs to final concentrations of 100, 50, 25 and 12.5µg/ml in sample diluent. 100ul of each sample was added to assay plates along with kit controls and the kit SOP was followed for the remainder of the assay. IgG phospholipid units (GPL) were calculated against a linear standard curve per kit instructions. An individual well was positive if GPL was >20, negative if <15 and indeterminate if between 15-20. Any given antibody needed to be positive for two consecutive wells (i.e. to at least 50µg/ml) to be considered cardiolipin positive; see values with yellow fill in **Table S6**.

### Neutralization Assays

Neutralizing antibodies were measured in either TZM-bl cells ^69^ or TZM-bl/FcγRI cells ^35^ as a function of reductions in luciferase (Luc) reporter gene expression as described ^70^. The NIH AIDS Reagent Program provided TZM-bl cells. In both assays, a pre-titrated dose of virus was incubated with serial 3-fold or 5-fold dilutions of either serum (heat-inactivated 56°C, 30 minutes), purified IgG or mAbs in duplicate in a total volume of 150 µl for 1 hr at 37°C in 96-well flat-bottom culture plates. Freshly trypsinized cells (10,000 cells in 100 µl of growth medium containing 75 µg/ml DEAE dextran) were added to each well. One set of 8 control wells received cells + virus (virus control) and another set of 8 wells received cells only (background control). After 48 hours of incubation, 100 µl of cells was transferred to a 96-well black solid plate for measurements of luminescence using the Bright-Glo Luciferase Assay System (Promega). Neutralization titers are the dilution (serum samples) or concentration (purified IgG and mAbs) at which relative luminescence units (RLU) were reduced by 50% or 80% compared to virus control wells after subtraction of background RLUs. Assay stocks of molecularly cloned Env-pseudotyped viruses were prepared by transfection in 293T/17 cells (American Type Culture Collection) and titrated in TZM-bl cells as described ^70^. The software used to collect data for neutralization assays is Promega GloMax Navigato. The software used to analyze neutralization data is LabKey Server NAb Tool.

Forty-nine MPER+ neutralizing antibodies constituted 24 clonal lineages from either vaccinee 133-23 or 133-39 (**Table S8**). The 24 neutralizing B cell clonal lineages neutralized heterologous tier 1 HIV-1 in the TZM-bl/FcγRI assay with geometric mean IC50 neutralization titers for W61D (0.18 µg/ml) and HXB2 (0.05 µg/ml), as well as heterologous tier 2 HIV-1 in the in the TZM-bl/FcγRI neutralization assay with geometric mean IC50 neutralization titers for JRFL (0.11 µg/ml), WITO (0.11 µg/ml) and SC422661.8 (0.09 µg/ml). Of the 24 B cell clonal lineages that neutralized in the TZM-bl/FcγRI neutralization assay, 14 clones also neutralized heterologous tier 1 HIV-1 in the standard TZM-bl assay with geometric mean IC50 neutralization titers for W61D (4.2 µg/ml) and HXB2 (10 µg/ml). Geometric mean titers were calculated in excel using the positive neutralization values shown in **Table S8**; geometric mean titers were calculated for each virus, separately.

### Signature Analyses and Neutralization Profile Comparison

Env neutralization signature analyses for DH1317.4 and DH1317.9 were performed as previously described ^56^. Briefly, binary phenotypes of DH1317.4 and DH1317.9 sensitivity or resistance (IC50 >50 or <50µg/ml) were used. To correct for multiple testing, a false discovery rate (q-value) of <0.2 was used, and to correct for clade effects, phylogenetically corrected signatures were also calculated. Robust signature sites were identified by using the criteria that signatures at these sites had q < 0.2 and such sites were either in epitope sites (658 to 668) or had a phylogenetically corrected signature, or both. Signatures were visualized using sequence logos for the sensitive vs. resistant group of viruses (**Figure S5**) calculated using AnalyzeAlign tool on the Los Alamos HIV Databases (https://www.hiv.lanl.gov/content/sequence/ANALYZEALIGN/analyze_align.html). For calculating inter-subtype variation at key signature sites in database strains, the “Filtered web” sequence alignment in AnalyzeAlign was used. For comparison of neutralization profiles between bNAbs, linear regression and Fisher’s exact test modules from SciPy were used (www.scipy.org) ^71^. The 20 HVTN 704 subtype B pseudoviruses used for the analysis reported in **Figure S5c** were from Gilbert *et al.* ^72^.

### Antibody Binding to Binary Phospholipids

PC-PG binary lipids were prepared by mixing 1-palmitoyl-2-oleoyl-glycero-3-phosphocholine (POPC, Avanti) with 1,2-dioleoyl-sn-glycero-3-phospho-(1’-rac-glycerol) (18:1 (Δ9-Cis) PG, Avanti) that had been solubilized in chloroform to a concentration of 10mM with a 25:75 ratio of PC to PG. The PC-PG lipid mixture was dried under a steady stream of nitrogen and then placed in a vacuum for 3-4 hours to remove any trace amounts of chloroform. The lipids were rehydrated in 500uL of PBS 1X pH7.4 and incubated at 37°C for 45 minutes. After incubation, the lipid preparation was vortexed and sonicated for 45s followed by extrusion through a 0.1um polycarbonate membrane using a 1-mL mini-extruder (Avanti).

The binding of Abs was performed by surface plasmon resonance (SPR) using the Biacore3000 platform (Cytiva) in PBS 1X pH7.4 running buffer. A lipophilic L1 chip (Cytiva) was docked and prepared by immobilizing BSA 1% to a level of 2000-3000RU to reduce non-specific binding of mAbs. A 1:5 dilution of PC-PG in PBS 1X was prepared and then manually captured on the flow cells of the L1 chip at 5uL/min to a level of 400-500RU. At least one flow cell had no captured lipid and would be used for reference subtraction. After a series of wash steps, mAb diluted down to 100ug/mL was injected over the sensor surface at 20uL/min for 2 minutes followed by a dissociation of at least 5 minutes. 13H11 and 2F5 were used as negative and positive control mAbs, respectively. The lipid was then regenerated from the sensor using a 60s pulse of 40mM OGP at 100uL/min. Results were analyzed using BiaEvaluation Software (Cytiva). Binding to blank flow cells with no lipid capture was used for reference subtraction and to account for non-specific binding. Binding values were measured at the end of association period and then normalized with respect to the variable lipid capture values. Reported results are an average of at least two measurements.

### Lipid Insertion Propensity Scores

Hydropathy analysis tool MPEx ^73^ (Membrane Protein Explorer) v3.3.0 with the water-interface scale was used to calculate lipid insertion propensity scores as the sum of Δ*G*_wif_, the free energy of transfer of an amino acid from water to POPC interface ^74^, over all amino acids in the CDR regions of both light and heavy chains. CDR amino acid positions were defined using the software ANARCI ^75^ with the IMGT scheme. Custom R scripts were used to compute scores for all CDRs individually. The custom script used in this manuscript for Ab lipid propensity scoring calculations has been deposited to GitHub (https://github.com/WieheLab/HVTN133). Calculation and significance test for the Pearson correlation coefficients were performed using the R function stat_corr from package *ggpubr;* https://rpkgs.datanovia.com/ggpubr/).

### Antibody and Fab Binding and Titrations against MPER Liposome Immunogen

The binding of antibodies to the MPER656 peptide-liposome immunogen (IDRI, Lot: 18P001) was performed using biolayer interferometry (BLI) and the OctetRed 96 system (Sartorius) in PBS 1X pH7.4 buffer. The sequence of MPER656-GTH1 peptide used in the MPER peptide-liposome is ^656^NEQELLELDKWASLWNWFNITNWLWYIK^683^YKRWIILGLNKIVRMYS). MPER peptide-liposomes were prepared at a 1:200 dilution in PBS and captured using APS (aminopropysilane) sensor tips to a level of approximately 1.0-1.5nm. The duration of liposome capture was 600s. The liposome loaded sensor tips were then coated with 0.01% BSA for 600s to block any non-specific interaction of the antibody with the sensor tips. The sensor tips were then washed with PBS for 120s. After washing with PBS 1x, the sensor tips were submerged into the mAbs (or Fabs) diluted down to 100µg/mL for an association length of 600. 13H11 was used as a negative control reference mAb while 2F5 and 2F5UA were used at positive control mAbs. Antibody binding analysis was performed using the ForteBio Data Analysis 10.0 software (Sartorius). The Y-axis was aligned to the baseline from 175s to 179.8s and the inter-step correction was aligned to dissociation. 13H11 binding was subtracted from the binding of each antibody for reference subtraction and to account for any non-specific binding. Binding curves are representative of one dataset.

For the Fab and mAb titrations against MPER656 peptide-liposomes, the same BLI procedure was performed as in the screening analysis. However, the samples were diluted from 1.25ug/mL to 50ug/mL and then individual MPER peptide-liposome captured sensors were submerged into these wells. Curve fitting analyses were performed in the BiaEvaluation Software (Cytiva) after 13H11 subtraction using the 1:1 model for most samples. For DH1317.4 Fab and DH1317.8 Fab, the two state conformational change model was used for fitting. Kinetic results and affinities are representative of one dataset.

### F(ab) Preparation from Purified IgG Samples

F(ab) preparation was performed using a modified protocol previously described ^76^. Purified IgG samples were dialyzed into 20mM sodium phosphate, 10mM EDTA, pH 7.0 and then concentrated to 20mg/ml using centrifugal filtration (Amicon). IgGs were then digested using papain-agarose resin (Thermo Fisher Scientific) in 20mM sodium phosphate, 10mM EDTA, 20mM cysteine, pH7.4 for 5 hours at 37°C. A few select mAbs were digested with Lys-C instead of papain-agarose. F(ab) fragments were separated from undigested IgG and Fc fragments through a 1 hour incubation with rProtein A Sepharose Fast Flow (Cytiva). Finally, cysteine was removed by centrifugal filtration (Amicon) and buffer exchange to 1x PBS, pH 7.4.

Quality of F(ab) was measured via SDS-PAGE and size exclusion chromatography (SEC). SDS-PAGE was conducted by loading 2ug protein/lane on a 4% to 15% TGX stain-free gel (Bio-Rad) under reducing and nonreducing conditions. Loaded gels were run at 200V in Tris-glycine-SDS buffer. Bands were visualized using Gel Doc EZ imager (Bio-Rad), and band size was assessed with a protein standard ladder (Bio-Rad). SEC was performed by loading 15ug protein on to a Superdex 200 increase 10/300 column using a 100-μl loop and run at 0.5ml/min using an Äkta Pure system (Cytiva). F(ab) peaks were analyzed with the system’s Unicorn 7.6.0 software. Molecular weight was estimated using a linear regression calculated by running a mix of known-molecular-weight protein standards (Cytiva) on the same column.

### Titrations of F(ab) to SP62 Peptide and Mutants

Affinities of F(ab)s to SP62 peptide (^652^QQEKNEQELLELDKWASLWN^671^) and alanine-substituted mutants were measured through Surface Plasmon Resonance (SPR) using a BIAcore S200 instrument (Cytiva) in HBS-EP+ 1x running buffer at 25°C. Biotinylated SP62 peptides and mutants were diluted to 20 ug/ml and injected over a streptavidin immobilized CM3 or CM4 chip surface at 5 uL/min. 80 – 120 response units (RU) of ligand was captured over flow cells 2 – 4. Flow cell 1 was used as a blank for reference subtraction of nonspecific binding to chip surface and streptavidin. Ligand capture was followed with long dissociation period of 30 mins or until response stabilized. F(ab)s were diluted to various concentration ranges between 0.25 nM and 400 nM and injected at a flow rate of 50 uL/min over flow cells 1 – 4 using single-cycle kinetics injection type. Six 120 s injections of F(ab) at increasing concentrations with 720 s dissociation after final injection. Chip surfaces were regenerated with Glycine, pH 2.0 for 30 s between cycles. Blank running buffer titration was used to account for signal drift in addition to the blank flow cell for double reference subtraction. Curve fitting of results was performed through BIAcore S200 evaluation software (Cytiva). Fitting made use of either heterogenous ligand model or 1:1 Langmuir model with global Rmax. Titration curves are representative of 2 data sets.

### Protein Expression and Purification

Plasmid encoding JRFL.SOSIP.v6.683 or JRFL.SOSIP.v6.delCT or full-length JRFL.SOSIP.v6 was mixed with plasmid encoding furin in 1:4 ratio. This plasmid mixture was transfected in GnTI-cells (ATCC) at density of 2×10^6^ cells/ml using 293Fectin (Thermo Fisher) according to manufacturer’s protocol. The cells were harvested 38-40 hours following transfection spinning down the transfected cell culture for 30 min at 3400 x g, and the supernatant was discarded. The cell pellet was solubilized with 200 ml lysis buffer (0.5% v/v Triton X-100, 50 mM Tris pH 7.4, 150 mM NaCl) with the addition of 2mg of PGT145 IgG and Pierce protease inhibitor tablets (Thermo Fisher) and incubated at 4°C for 30 min. The cell debris were spun down again at 4347 x g for 1 hour followed by filtration through a 0.8 µm filter. The supernatant was incubated overnight with 1 ml Ultra Protein A resin (Thermo Fisher) and 2 mg of VRC01 IgG at 4°C. On day 2 of the purification, the unbound cell lysate was removed by passing the resin containing supernatant over a gravity column. The resin was washed with 10 CV of each of the following buffers, buffer 1 (0.1% w/v CHAPS, 50 mM Tris pH 7.4, 150 mM NaCl, 0.03 mg/mL sodium deoxycholate), buffer 2 (0.1% w/v *n*-Dodecyl β-D-maltoside (DDM), 50 mM Tris pH 7.4, 500 mM NaCl, 0.03 mg/mL sodium deoxycholate), and buffer 3 (0.1% w/v DDM, 50 mM Tris pH 7.4, 150 mM NaCl, 0.03 mg/mL sodium deoxycholate, 2 mM EDTA) successively. The complex of Env with the IgG bound to the Protein A resin was digested on-column using Lys-C (1 µg Lys-C for 20 µg IgG) in wash buffer 3 at room temperature for 4 hours. Following digestion, the flow through containing the Fab-bound Env followed by a 5 CV wash of the resin with the SEC buffer (0.1% w/v DDM, 50 mM Tris pH 7.4, 150 mM NaCl, 0.03 mg/mL sodium deoxycholate), and concentrated using 100,000 MWCO concentrator to ∼500 μL and further purified by size exclusion chromatography (SEC) using Superose 6 Increase 10/300 GL column (Cytiva) preequilibrated in SEC buffer.

### Surface Plasmon Resonance Measurement of Env Trimer Binding

Binding experiments for Fabs were performed using SPR on a Biacore T-200 (Cytiva, MA) with HBS buffer supplemented with 3 mM EDTA and 0.05% surfactant P-20 (HBS-EP+, Cytiva, MA). All binding assays were performed at 25 °C. DH1317.4 Fab binding to JRFL.SOSIP.v6.683 was assessed using a Series S CM5 chip (Cytiva, MA) which was coated with anti-human IgG (fc) Ab using a Human Ab Capture Kit (Cytiva, MA). 2G12 IgG was captured on the anti-human Fc chip by flowing over a 200 nM solution for 120s at 5 µL/min flow rate, followed by Env at 200 nM (120s at 5 µL/min). DH1317.4 Fab was injected at concentrations ranging from 12.5 nM to 200 nM (prepared in a 2-fold serial dilution manner) over the chip surface using the single cycle kinetics mode with 5 concentrations per cycle. The surface was regenerated three pulses of a 3 M MgCl_2_, pH 7.0 solution for 10 seconds at 100µL/min. Sensogram data were analyzed using the BiaEvaluation software (Cytiva, MA).

### Protein Crystallization

Fragments of antigen binding (Fabs) of antibodies DH1317.8, DH1322.1, and DH1346 were buffer exchanged into 50 mM Sodium Acetate pH 5.2 and loaded onto a Mono S 5/50 ion exchange column (Cytiva) equilibrated in the same buffer. Loaded Fabs were eluted on a continuous gradient using 50 mM Sodium Acetate pH 5.2 supplemented with 500 mM Sodium Chloride, followed by size exclusion chromatography using a Superdex 200 Increase 10/300 column (Cytiva) equilibrated in gel filtration buffer (GFB) composed of 2.5 mM Tris Cl pH 7.5, 150 mM Sodium Chloride, and 0.002% sodium azide.

For purified DH1317.8 Fab, complexes were prepared with MPER peptides MPR.03, KKK^656^NEQELLELDKWASLWNWFDITNWLWYIRK^684^KK, and SP62, ^652^QQEKNEQELLELDKWASLWN^671^, at 3-fold molar excess. Sitting drop vapor diffusion crystallization screens were set up with a Mosquito Nanoliter Liquid Handling System (SPT Labtech, Concord, CA) using a total of 768 conditions (384 unique conditions for each complex) obtained from commercially available screens (Rigaku Reagents, Woodlands, TX, and Hampton Research, Aliso Viejo, CA). Diffracting crystals of the DH1317.8 Fab-SP62 peptide complex were obtained in condition composed of 20.1% Isopropanol, 10.05% PEG 8000, and 0.1M Imidazole/Hydrochloric acid pH 6.5 which were optimized and cryoprotected in (2R,3R)-(−)-2,3-Butanediol prior to flash freezing in liquid nitrogen for data collection.

For purified DH1322.1 Fab, complexes were prepared with peptide K^651^NQQEKNEQELLELDKWASLWN^671^K at 3-fold molar excess. Sitting drop vapor diffusion crystallization screens were set up with a Mosquito Nanoliter Liquid Handling System (SPT Labtech, Concord, CA) using a total of 672 conditions obtained from commercially available screens (Rigaku Reagents, Woodlands, TX, and Hampton Research, Aliso Viejo, CA). Diffracting crystals were obtained in a condition composed of 25% PEG 1500 and 0.1M SPG buffer pH 8.5, which were flash frozen in liquid nitrogen for data collection.

For purified DH1346 Fab, complexes were prepared with peptide MPR.03 (see sequence above) at 3-fold molar excess. Sitting drop vapor diffusion crystallization screens were set up using Mosquito Nanoliter Liquid Handling System (SPT Labtech, Concord, CA) using a total of 672 conditions obtained from commercially available screens (Rigaku Reagents, Woodlands, TX, and Hampton Research, Aliso Viejo, CA). Diffracting crystals were obtained in a condition composed of 25% PEG 1500, 0.1M Sodium acetate/acetic acid pH4.5, 30% MPD, and 30% dextran sulfate sodium salt, which were flash frozen in liquid nitrogen for data collection.

### X-ray crystallography

X-ray diffraction data were collected at the ID-22 beamline (SERCAT) at the Advance Photon Source (Arognne, Il) or at the BM12-2 beamline at the Stanford Synchrotron Radiation Lightsource (Menlo Park, CA). Collected datasets were processed using HKL-2000 or XDS ^77, 78^. Structures were solved by molecular replacement using PHASER, with search model PDB 4NRX for DH1317.8, and the resulting DH1317.8 structure as a search model for DH132.1 and DH1346 ^79^. Refinement was carried out using PHENIX with manual iterative model building in Coot ^80, 81^. Protein interfaces and residue buried surface areas were determined using Protein Interfaces, Surfaces and Assemblies (PISA) ^82^. The PyMOL Molecular Graphics System and GraphPad PRISM were used to produce graphical images and plots.

Statistical correlations between interface residue buried surface areas of shared MPER epitope residues between the V_H_7-4-1-using nAbs themselves and against other proximal MPER-targeting antibodies were determined by testing the null hypothesis that they were independent. The null hypothesis was that the correlations were zero (no correlation), and the alternative hypothesis was that correlations were non-zero. For the three V_H_7-4-1-using nAbs tested against each other, the null hypothesis was rejected by a significant *p*-value, supporting the possibility they had a positive correlation with each other. For the correlation of representative V_H_7-4-1 nAb DH1317.8 against DH570, m66, and 2F5, the results did not reject the null hypothesis, suggesting they had no correlation. Statistical tests and results for the analysis were generated in GraphPad PRISM using the Pearson correlation function.

### Negative Stain Electron Microscopy (NSEM)

For forming complexes, 1-5 µg JR-FL SOSIP.683 trimer was mixed with 6 molar concentration of Fab and incubated at 4 °C overnight. A portion of complex was then diluted to 20-50 µg/ml of SOSIP with buffer containing 20 mM HEPES, pH 7.4, 150 mM NaCl and 0.02 g/dL Ruthenium Red. A 5-µl drop of diluted sample was then applied to a glow-discharged, carbon-coated EM grid for 8-10 second, blotted, consecutively rinsed with 2 drops of dilute buffer containing 1 mM HEPES, pH 7.4 and 7.5 mM NaCl, and immediately blotted, stained with 2 g/dL uranyl formate for 1 min, blotted, and air-dried. Grids were examined on a Philips EM420 electron microscope operating at 120 kV and nominal magnification of 49,000x, and ∼120 images for each sample were collected on a 76 Mpix CCD camera at a nominal calibration of 2.2 Å/pixel. Images were analyzed and 3D reconstructions calculated with Relion 3.0 ^83^. Data flow for symmetry expansion and focused 3D classification of the MPER Fabs is described in **Figure S14**. NSEM data were collected with software provided for the camera (Scientific Instruments and Applications, Inc) and analyzed using Relion 3.0, and UCSF Chimera.

### Cryo-Electron Microscopy (cryo-EM)

DH1317.4 Fab was added at a 6-fold molar excess to the SEC-purified complex of full-length JR-FL Env bound to PGT145 and VRC01 Fabs, and the sample was incubated for 30 minutes. 2.5 µl of the sample was deposited on a glow-discharged QUANTIFOIL holey carbon grid (R1.2/1.3 Cu 300 mesh). After a 30 second incubation in 90-95% relative humidity, excess protein was blotted away with a Whatmann filter paper. The grid was plunge frozen in liquid ethane using an EM GP2 plunge freezer (Leica, Microsystems). A total of 46,998 movies with a dose rate ranging from 63-76 e^-^/ Å^2^ Cryo-EM data were collected using a 300 kV FEI Titan Krios electron microscope (ThermoFisher) equipped with a K3 camera (Gatan) with a pixel size of 1.08 Å with a defocus range between −1 and −3 μm. Gatan Latitude software was used to collect data. All data processing steps were carried out in cryoSparc (https://www.nature.com/articles/nmeth.4169). Movie frames were aligned and dose weighted, followed by CTF estimation, particle picking, 2D classifications, 3D classifications using ab initio and heterogenous classifications, followed by 3D refinement. Cryo-EM data collection and map refinement statistics are shown in **Table S14**.

### Reagent Authentication

Expi293F and TZM-bl cell lines were provided with a certificate of analysis from their sources. Cell identity is verified by morphology or fluorescent markers expressed. Positive control abs were used in the TZM-bl neutralization assay to distinguish the cell lines and for comparison between lots when new vials of cells are cultured.

### Data Availability

The data supporting the findings of this study are available within the main article and supplementary information. Any further data relevant to the study will be made available upon reasonable request to the corresponding authors.

### Code Availability

Custom scripts used in this manuscript for single B cell VDJ sequencing analysis and Ab lipid propensity scoring calculations have been deposited to GitHub (https://github.com/WieheLab/HVTN133). Additional softwares and codes used for data collection and analyses were outlined in the methods.

#### Accession codes

PDB IDs for x-ray crystal structures – 8G8A, 8G8C and 8G8D.

