## Supplemental Online Material - Figures and Tables for "Vaccine Induction of Heterologous HIV-1 Neutralizing Antibody B Cell Lineages in Humans"

### Figure S1

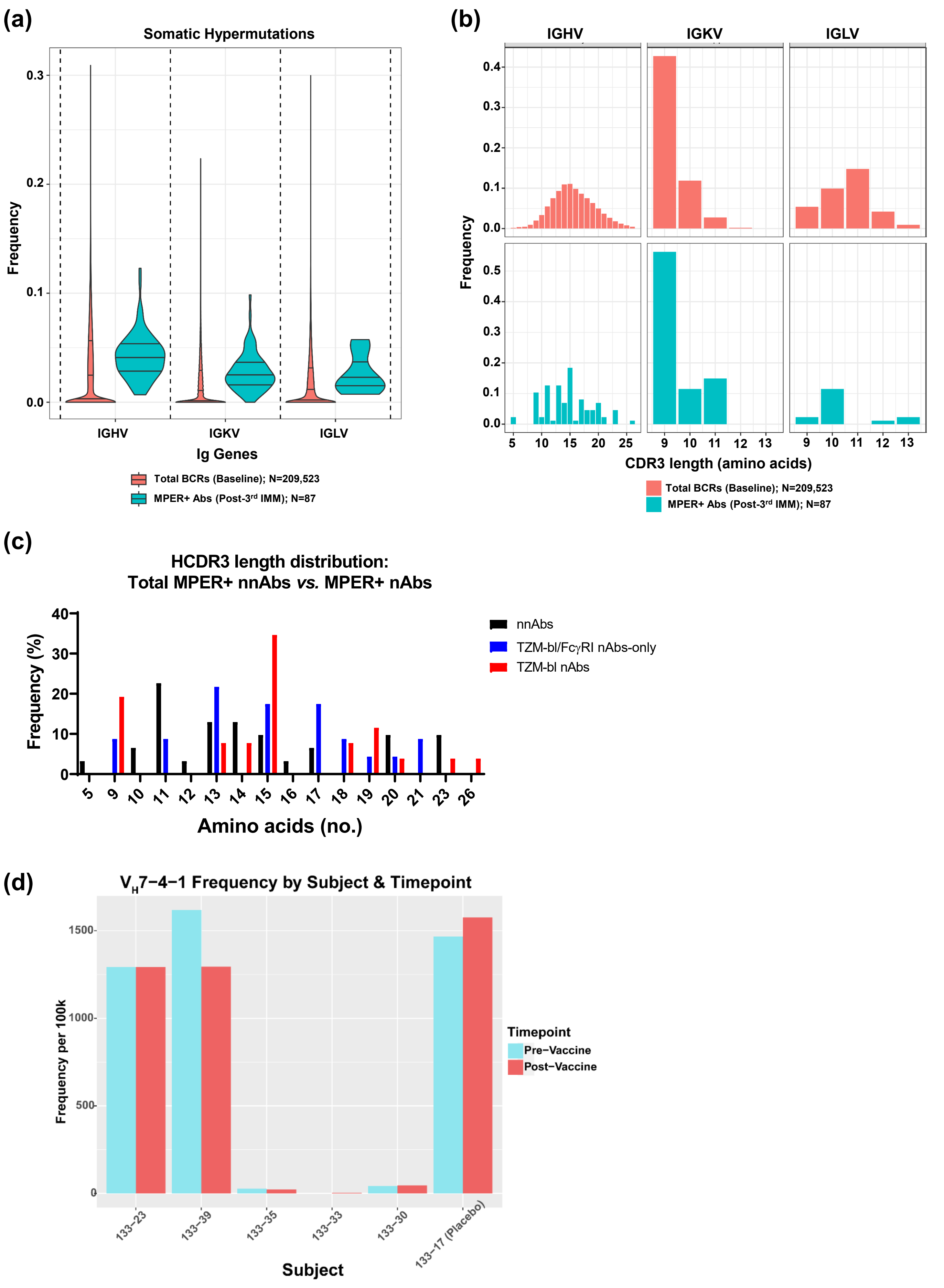
(a)
Somatic Hypermutations
Frequency
0.0
0.1
0.2
0.3
IGHV
IGKV
IGLV
Ig Genes
Total BCRs (Baseline); N=209,523
MPER+ Abs (Post-3rd IMM); N=87
(b)
IGHV
IGKV
IGLV
Frequency
0.0
0.1
0.2
0.3
0.4
0.5
5
10
15
20
25
9
10
11
12
13
CDR3 length (amino acids)
Total BCRs (Baseline); N=209,523
MPER+ Abs (Post-3rd IMM); N=87
(c)
HCDR3 length distribution:
Total MPER+ nnAbs *vs.* MPER+ nAbs
Frequency (%)
0
10
20
30
40
5
23
26
Amino acids (no.)
nnAbs
TZM-bl/FcγRI nAbs-only
TZM-bl nAbs
(d)
$V_H$7−4−1 Frequency by Subject & Timepoint
Frequency per 100k
0
500
1000
1500
133−23
133−39
133−35
133−33
133−30
133−17 (Placebo)
Subject
Timepoint
Pre−Vaccine
Post−Vaccine

**Figure S1. Immunogenetics of MPER+ Antibody Response. (a)** Somatic hypermutation (SHM) frequency in Ig genes used by MPER+ antibodies (Abs) isolated post-3rd IMM, as well as antigen-naïve Abs in our reference dataset of total BCRs. Mutation frequency is reported for nucleotide sequences. The data are shown in violin plots; the middle line is the median, and then the quartiles shown by the upper and lower lines. **(b)** HCDR3 length distributions for MPER+ Abs isolated post-3rd IMM, as well as antigen-naïve Abs in our reference dataset of total BCRs. HCDR3 length is reported in amino acids (count). (a-b) The total number of Abs studied per group is shown in the keys per panel. **(c)** Eighty MPER+ Abs representative of the 87 MPER+ Abs isolated post 3rd immunization from 5 HVTN133 vaccine recipients were tested for neutralization in the TZM-bl/Fc$\gamma$RI and standard TZM-bl neutralization assays. Shown is the frequency of MPER+ Abs with varied HCDR3 length categorized as MPER+ nnAbs (N=31; black) and those that neutralized in the TZM-bl/Fc$\gamma$RI (N=23, blue) and TZM-bl (N=26, red) assays. **(d)** Frequency of $V_H$7-4-1-using BCRs detected among antigen-naïve BCRs at baseline in the five vaccine recipients (133-23, 133-30, 133-33, 133-35 and 133-39) and one placebo recipient (133-17) who received three immunizations.

### Figure S2

**(a)** $V_H$7-4-1 nAb clones

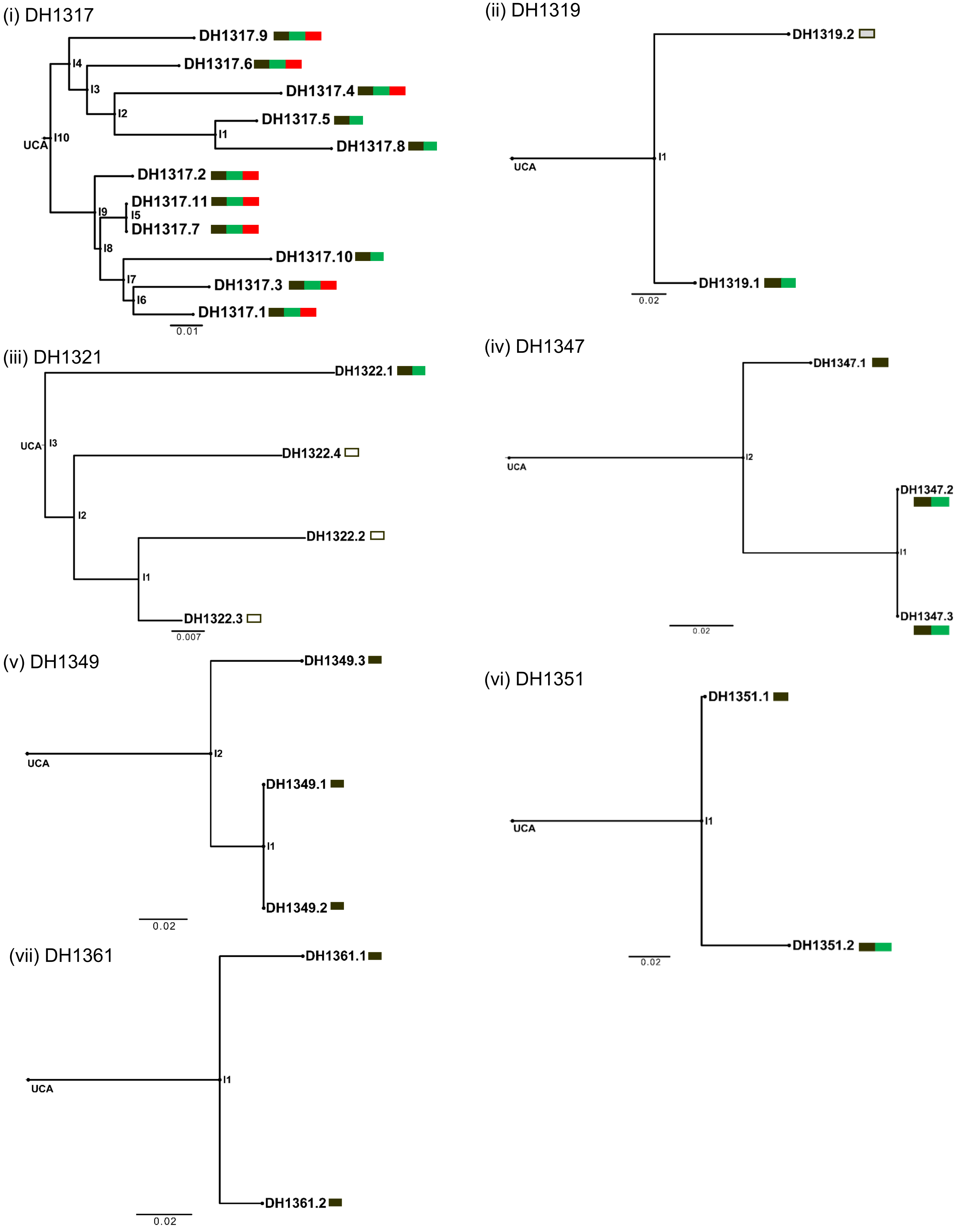

### Figure S2

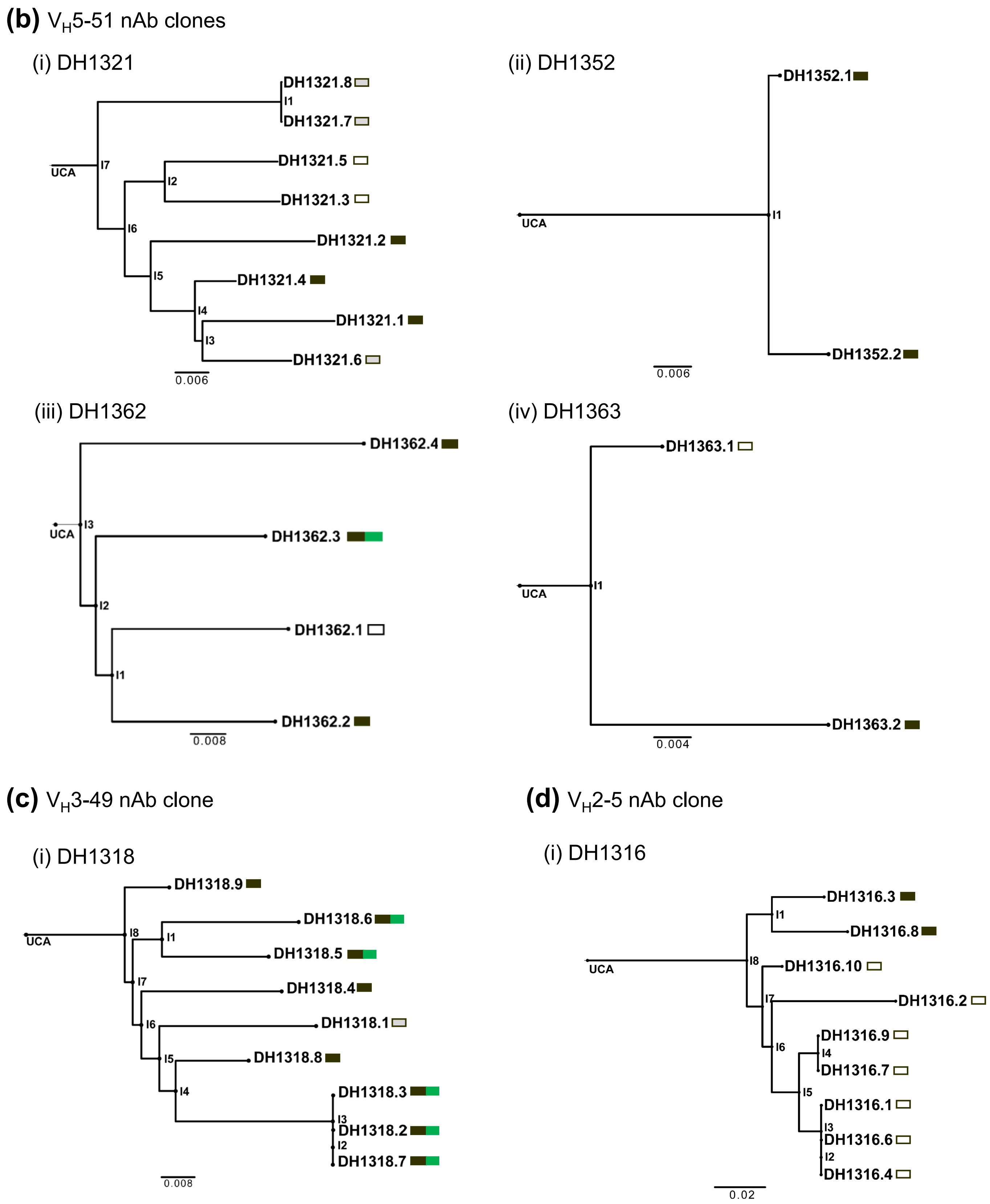

(b) $V_H$5-51 nAb clones
(i) DH1321
DH1321.8
I1
DH1321.7
UCA
I7
DH1321.5
I2
DH1321.3
I6
DH1321.2
I5
DH1321.4
I4
DH1321.1
I3
DH1321.6
0.006
(ii) DH1352
DH1352.1
UCA
I1
DH1352.2
0.006
(iii) DH1362
DH1362.4
UCA
I3
DH1362.3
I2
DH1362.1
I1
DH1362.2
0.008
(iv) DH1363
DH1363.1
UCA
I1
DH1363.2
0.004
(c) $V_H$3-49 nAb clone
(i) DH1318
DH1318.9
UCA
I8
DH1318.6
I1
DH1318.5
I7
DH1318.4
I6
DH1318.1
I5
DH1318.8
I4
DH1318.3
I3
DH1318.2
I2
DH1318.7
0.008
(d) $V_H$2-5 nAb clone
(i) DH1316
DH1316.3
I1
DH1316.8
UCA
I8
DH1316.10
I7
DH1316.2
I6
DH1316.9
I4
DH1316.7
I5
DH1316.1
I3
DH1316.6
I2
DH1316.4
0.02

**Figure S2. B Cell Clonal Expansion by MPER-peptide Liposome in HVTN133.** Shown are phylograms of clonally-related MPER+ Abs that contained an Ab that neutralized HIV-1 strains in TZM-bl/Fc$\gamma$RI or TZM-bl neutralization assay. NAb clones are grouped according to VH gene usages; **(a)** $V_H$7-4-1-using clones, **(b)** $V_H$5-51-using clones, **(c)** $V_H$3-49-using clones and **(d)** $V_H$2-5-using clones. The boxes on each phylogram summarizes the Ab neutralization profile against clade B HIV-1 tier 1 and 2 HIV-1 strains: black – tier 1 or 2 HIV-1 neutralization in TZM-bl/Fc$\gamma$RI assay; green – tier 1 HIV-1 neutralization in TZM-bl assay; red – tier 2 HIV-1 neutralization in TZM-bl assay; white – neutralization negative in both assays; and gray – not tested. Not shown are clones with a single lineage member: $V_H$7-4-1-using Abs—DH1346, DH1350, DH1413, and DH1425; $V_H$5-51-using Abs—DH1354, DH1360, DH1420, DH1421 and DH1423; $V_H$3-73-using Ab—DH1353; and $V_H$1-69-using Ab—DH1355.

### Figure S3

| IC50, μg/mL | <0.1 | 0.1-1 | 1-10 | 10-50 | >50 |
|---|---|---|---|---|---|

| Virus ID | Neutralization Tier | Clade | Titer in TZM-bl neutralization assay (μg/ml) DH1317.4 IC50 | IC80 | MPI |
|---|---|---|---|---|---|
| Q461.e2 | 2 | A | >50 | >50 | 43 |
| Q259.d2.17 | 2 | A | >50 | >50 | 32 |
| 0260.v5.c36 | 2 | A | >50 | >50 | 22 |
| 0330.v4.c3 | 2 | A | >50 | >50 | 48 |
| 191955_A11 | 2 | A | >50 | >50 | 5 |
| 9004SS_A3_4 | 2 | A | >50 | >50 | 10 |
| Q23.17 | 1B | A | >50 | >50 | 17 |
| MS208.A1 | | A | >50 | >50 | 10 |
| 6041.v3.c23 | 2 | AC | >50 | >50 | 34 |
| 6540.v4.c1 | 2 | AC | >50 | >50 | 16 |
| 246F3 | 2 | AC | >50 | >50 | 4 |
| 0815.v3.c3 | 2 | ACD | >50 | >50 | 31 |
| 3103.v3.c10 | 2 | ACD | >50 | >50 | 8 |
| CNE5 | | AE | >50 | >50 | 14 |
| CNE59 | | AE | >50 | >50 | 1 |
| T242-14 | | AG | >50 | >50 | 16 |
| T253-11 | | AG | >50 | >50 | 32 |
| 6535.3 | 1B | B | 44.0 | >50 | 56 |
| QH0692.42 | 2 | B | >50 | >50 | 13 |
| PVO.4 | 3 | B | >50 | >50 | 26 |
| TRO.11 | 2 | B | >50 | >50 | 8 |
| AC10.0.29 | 2 | B | >50 | >50 | 18 |
| RHPA4259.7 | 2 | B | >50 | >50 | 12 |
| THRO4156.18 | 2 | B | >50 | >50 | 24 |
| REJO4541.67 | 2 | B | 11.9 | >50 | 75 |
| TRJO4551.58 | 3 | B | >50 | >50 | 10 |
| CAAN5342.A2 | 2 | B | >50 | >50 | 6 |
| 3988.25 | 2 | B | >50 | >50 | 0 |
| 5768.4 | 2 | B | 4.6 | 36.2 | 91 |
| 6101.10 | 2 | B | >50 | >50 | 25 |
| 7165.18 | 2 | B | 19.8 | 48.3 | 81 |
| CNE10 | 2 | B | 22.0 | >50 | 74 |
| CNE12 | 2 | B | 13.5 | >50 | 79 |
| CNE14 | 2 | B | 26.5 | >50 | 70 |
| CNE4 | 2 | B | 26.3 | >50 | 65 |
| CNE57 | 2 | B | 9.0 | 34.4 | 88 |
| 45_01dG5 | 2 | B | 3.2 | 12.9 | 96 |
| QH0515.1 | 2 | B | 3.6 | 27.3 | 90 |
| X2278_C2_B6 | 2 | B | >50 | >50 | 0 |
| H704_0855_080_EsN | AMP trial | B | >50 | >50 | 6 |
| H704_2684_181_RE_p0001s | AMP trial | B | >50 | >50 | 53 |
| H704_0026_231_RE_pbsga001_s | AMP trial | B | >50 | >50 | 1 |
| H704_0907_130sN | AMP trial | B | >50 | >50 | 23 |
| H704_1528_240_RE_pblib_001_s | AMP trial | B | >50 | >50 | 0 |
| H704_0726_080sN | AMP trial | B | >50 | >50 | 1 |
| H704_1535_030sN | AMP trial | B | >50 | >50 | 15 |
| H704_0911_150sN | AMP trial | B | >50 | >50 | 5 |
| H704_2767_070sN | AMP trial | B | >50 | >50 | 4 |
| H704_1706_050sN | AMP trial | B | >50 | >50 | 1 |
| H704_0644_060sN_prelimSeq | AMP trial | B | >50 | >50 | 1 |
| H704_0847_030_EsN_01T | AMP trial | B | >50 | >50 | 0 |
| H704_0575_060_RE_p002s | AMP trial | B | >50 | >50 | 0 |
| H704_1991_230_RE_p001s_1194T | AMP trial | B | 12.6 | >50 | 74 |
| H704_1835_150_RE_p001s_2484A | AMP trial | B | >50 | >50 | 7 |
| H704_3008_040EsN | AMP trial | B | >50 | >50 | 27 |
| H704_1180_070EsN | AMP trial | B | >50 | >50 | 41 |
| H704_2541_080EsN | AMP trial | B | >50 | >50 | 45 |
| H704_2448_240_RE_cs | AMP trial | B | >50 | >50 | 0 |
| H704_2095_130_RE_cs | AMP trial | B | >50 | >50 | 0 |
| V704_0372_250_RE_pblib001_s | AMP trial | B | >50 | >50 | 48 |
| WEAU_d15_410_787 | 2 | B | 15.0 | >50 | 77 |
| 1006_11_C3_1601 | 2 | B | >50 | >50 | 0 |
| 1054_07_TC4_1499 | 2 | B | >50 | >50 | 0 |
| 1056_10_TA11_1826 | 1B | B | 46.4 | >50 | 52 |
| 1012_11_TC21_3257 | 1B | B | >50 | >50 | 0 |
| 6240_08_TA5_4622 | 2 | B | >50 | >50 | 4 |
| 6244_13_B5_4576 | 2 | B | >50 | >50 | 5 |
| 62357_14_D3_4589 | 2 | B | >50 | >50 | 44 |
| SC05_8C11_2344 | 2 | B | >50 | >50 | 0 |
| H022.7 | 2 | B | >50 | >50 | 0 |
| H029.12 | 1B | B | >50 | >50 | 0 |
| H030.7 | 2 | B | 34.6 | >50 | 62 |
| H031.7 | 2 | B | 3.3 | 19.7 | 91 |
| H035.18 | 3 | B | >50 | >50 | 45 |
| H061.14 | 3 | B | 43.8 | >50 | 52 |
| H079.2 | 3 | B | >50 | >50 | 11 |
| H086.8 | 1B | B | >50 | >50 | 49 |
| H077.31 | 1B | B | 32.7 | >50 | 57 |
| H078.14 | 3 | B | >50 | >50 | 0 |
| H080.23 | 2 | B | >50 | >50 | 6 |
| 700010040.C04520 | 2 | B | >50 | >50 | 5 |
| PRB926-04.A9.4237 | 2 | B | 47.8 | >50 | 51 |
| 9021-14.B2.4571 | 2 | B | 44.7 | >50 | 52 |
| W61D(TCLA).71 | 1 | B | 0.1 | 0.5 | 100 |
| HXB2 | 1 | B | 1.2 | 3.9 | 100 |
| WITO4160.33 | 2 | B | 40.7 | >50 | 59 |
| JR-FL | 2 | B | 13.1 | 34.4 | 90 |
| SC422661.8 | 2 | B | 16.0 | >50 | 81 |
| TRO11 | 2 | B | >50 | >50 | -1 |
| CNE17 | 2 | BC | >50 | >50 | 1 |
| 25710 | 2 | C | >50 | >50 | 8 |
| CE0217 | 2 | C | >50 | >50 | 9 |
| Du422.1 | 2 | C | >50 | >50 | 10 |
| ZM214M.PL15 | 2 | C | >50 | >50 | 0 |
| ZM233M.PB6 | 2 | C | >50 | >50 | 8 |
| ZM135M.PL10a | 2 | C | >50 | >50 | 5 |
| CAP45.2.00.G3 | 2 | C | >50 | >50 | 0 |
| CAP244.2.00.D3 | 3 | C | >50 | >50 | 18 |
| ZM215F.PB8 | 2 | C | >50 | >50 | 19 |
| ZM106F.PB9 | 2 | C | >50 | >50 | 9 |

| Virus ID | Neutralization Tier | Clade | Titer in TZM-bl neutralization assay (μg/ml) DH1317.4 IC50 | IC80 | MPI |
|---|---|---|---|---|---|
| 0041.v3.c18 | 2 | C | >50 | >50 | 4 |
| 0984.v2.c2 | 3 | C | >50 | >50 | 8 |
| 6040.v4.c15 | 2 | C | >50 | >50 | 20 |
| 6631.v3.c10 | 3 | C | >50 | >50 | 14 |
| 933.v4.c4 | 3 | C | >50 | >50 | 17 |
| 6980.v0.c31 | 2 | C | >50 | >50 | 9 |
| 3426.v5.c17 | 2 | C | >50 | >50 | 6 |
| Ce1172_H1 | 2 | C | >50 | >50 | 6 |
| Ce703010131_1E2 | 2 | C | >50 | >50 | 0 |
| Ce703010054_2A2 | 2 | C | >50 | >50 | 0 |
| Ce0665_F2 | 2 | C | >50 | >50 | 9 |
| Ce2052_G10 | 2 | C | >50 | >50 | 16 |
| Ce704810053_2B7 | 2 | C | >50 | >50 | 21 |
| CeCAP210_TA5 | 2 | C | >50 | >50 | 8 |
| CeCAP200_B8a | 1B | C | >50 | >50 | 12 |
| CeCAP188_1_D1_14(Rev-) | 2 | C | >50 | >50 | 0 |
| CeCAP177_1A3 | 2 | C | >50 | >50 | 5 |
| ZM249M.PL1 | 2 | C | >50 | >50 | 0 |
| 7060101641A7(Rev-) | 2 | C | >50 | >50 | 5 |
| 235080_3G7env2(Rev-) | 1B | C | >50 | >50 | 6 |
| 704010042 | 2 | C | >50 | >50 | 0 |
| 705010185 | 2 | C | >50 | >50 | 4 |
| 1245045 | 2 | C | >50 | >50 | 0 |
| 19157834_V1 | 2 | C | >50 | >50 | 5 |
| 2969249 | 2 | C | >50 | >50 | 17 |
| 3514597 | 2 | C | >50 | >50 | 0 |
| 20258279_V2 | 2 | C | >50 | >50 | 0 |
| 20915593 | 2 | C | >50 | >50 | 0 |
| 20927783 | 3 | C | >50 | >50 | 1 |
| 20965238 | 2 | C | >50 | >50 | 5 |
| 21197826_V1 | 3 | C | >50 | >50 | 10 |
| 21283649 | 3 | C | >50 | >50 | 1 |
| 2768732_C5_16 | 2 | C | >50 | >50 | 14 |
| 18814602_H8_F3 | 1B | C | >50 | >50 | 5 |
| 19252094_A5_G2 | 2 | C | >50 | >50 | 0 |
| 19707346_E8_C6 | 3 | C | >50 | >50 | 1 |
| 20104663_E11_D2 | 2 | C | >50 | >50 | 0 |
| 20198102_E9_G1 | 3 | C | >50 | >50 | 6 |
| 20286961_C1_H8 | 2 | C | >50 | >50 | 0 |
| 20883229_C9_H6 | 3 | C | >50 | >50 | 0 |
| 21203310_G7_C3 | 2 | C | >50 | >50 | 10 |
| 21492713_B11_E3 | 2 | C | >50 | >50 | 0 |
| 21502011_F12_E2 | 2 | C | >50 | >50 | 9 |
| 21561324_D3_B5 | 2 | C | >50 | >50 | 0 |
| 19314479_A2_5 | 2 | C | >50 | >50 | 5 |
| 234-F1-16-57 | 2 | C | >50 | >50 | 0 |
| 541-F1_A7_2 | 3 | C | >50 | >50 | 0 |
| 569-F1_37_10 | 2 | C | >50 | >50 | 1 |
| 556_F2_3_25 | 2 | C | >50 | >50 | 0 |
| CAP69.1.12_TA7.1 | 2 | C | >50 | >50 | 0 |
| CAP136.1.16_E6_1 | 2 | C | >50 | >50 | 0 |
| CAP174.1.06_F3_1B | 1B | C | >50 | >50 | 2 |
| CAP225.1.06_A2_18 | 3 | C | >50 | >50 | 0 |
| CAP266.2.00_E9_h6 | 2 | C | >50 | >50 | 0 |
| TRP290.2.00_23_6 | 2 | C | >50 | >50 | 0 |
| TRP307.2.00_24_1 | 2 | C | >50 | >50 | 0 |
| TRP363.2.00_10_3 | 2 | C | >50 | >50 | 3 |
| 722_G4_16 | 2 | C | >50 | >50 | 6 |
| So431_C1_1 | 2 | C | >50 | >50 | 0 |
| So405_T24_5 | 2 | C | >50 | >50 | 0 |
| CT072_56_7 | 1B | C | >50 | >50 | 0 |
| CT966_E1-7 | 2 | C | >50 | >50 | 0 |
| CT140_140_B6 | 2 | C | >50 | >50 | 0 |
| Ko426_T78_10 | 2 | C | >50 | >50 | 5 |
| Ko459_T68_4 | 2 | C | >50 | >50 | 30 |
| Ko870_C2_10 | 2 | C | >50 | >50 | 8 |
| 6644.v2.c33 | 2 | C | >50 | >50 | 18 |
| TV1.21 | | C | >50 | >50 | 7 |
| 3168.v4.c10 | | C | >50 | >50 | 15 |
| ZM197M.PB7 | 2 | C | >50 | >50 | 26 |
| CAP206.1.B5 | | C | >50 | >50 | 10 |
| CE1176 | 2 | C | >50 | >50 | 10 |
| 3817.v2.c59 | 2 | CD | >50 | >50 | 17 |
| 6952.v1.c20 | 2 | CD | 10.0 | 33.4 | 91 |
| C1080.c03 | 2 | CRF01_AE | >50 | >50 | 6 |
| R2184.c04 | 2 | CRF01_AE | >50 | >50 | 22 |
| R1166.c01 | 2 | CRF01_AE | >50 | >50 | 10 |
| C2101.c01 | 2 | CRF01_AE | >50 | >50 | 13 |
| C4118.c09 | 2 | CRF01_AE | >50 | >50 | 3 |
| CNE55 | 2 | CRF01_AE | >50 | >50 | 5 |
| T257-31 | 3 | CRF02_AG | >50 | >50 | 17 |
| 928-28 | 2 | CRF02_AG | >50 | >50 | 48 |
| T250-4 | 2 | CRF02_AG | >50 | >50 | 36 |
| T251-18 | 3 | CRF02_AG | >50 | >50 | 18 |
| T278-50 | 3 | CRF02_AG | >50 | >50 | 8 |
| BJOX2000 | 2 | CRF07_BC | >50 | >50 | 20 |
| CH119 | 2 | CRF07_BC | >50 | >50 | 9 |
| 3016.v5.c45 | 2 | D | 20.7 | >50 | 77 |
| A07412M1.vrc12 | 2 | D | >50 | >50 | 11 |
| 231965.c01 | 2 | D | >50 | >50 | 17 |
| 92UG024.2 | | D | >50 | >50 | 5 |
| T247-23 | | D | >50 | >50 | 1 |
| X1193_c1 | 2 | G | 15.9 | 48.9 | 81 |
| X1254_c3 | 2 | G | 14.7 | 48.9 | 81 |
| X2088_c9 | 2 | G | >50 | >50 | 3 |
| X1632_S2_B10 | 2 | G | >50 | >50 | 26 |
| X1632 | 2 | G | >50 | >50 | 20 |

**Figure S3. Neutralization Breadth of Vaccine-induced DH1317 BnAb.** Neutralization titers of DH1317.4 bnAb against 197 multiclade HIV-1 strains in the TZM-bl neutralization assay. Neutralization titers were measured in IC50 and IC80 (µg/ml). MPI: maximum percent inhibition.

### Figure S4

**a**

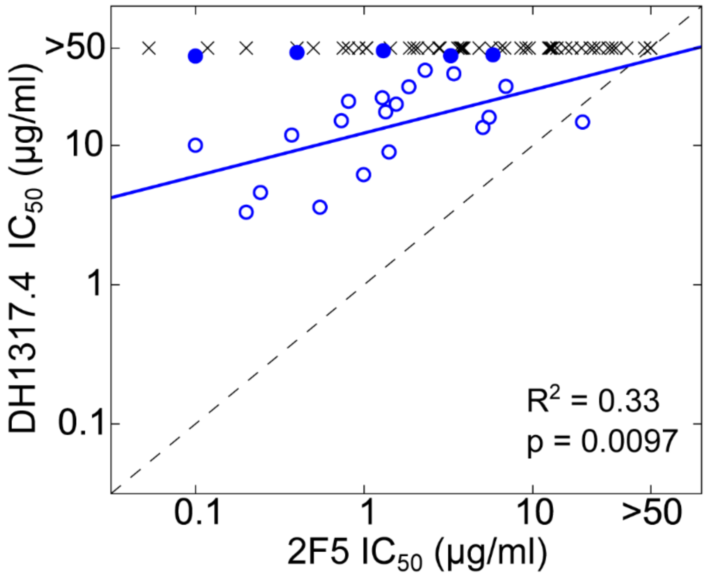

| | | DH1317.4 | |
|---|---|---|---|
| | | sen. | res. |
| 2F5 | sen. | 24 | 48 |
| | res. | 0 | 79 |

Accuracy = 0.68
Odds ratio = inf
p = 1.7 x $10^{-9}$

**b**

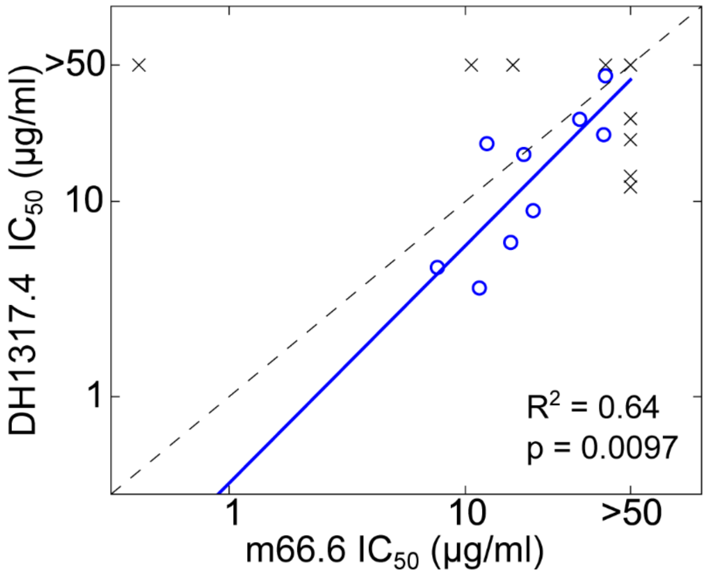

| | | DH1317.4 | |
|---|---|---|---|
| | | sen. | res. |
| m66.6 | sen. | 9 | 4 |
| | res. | 4 | 31 |

Accuracy = 0.73
Odds ratio = 17.4
p = 0.0002

**c**

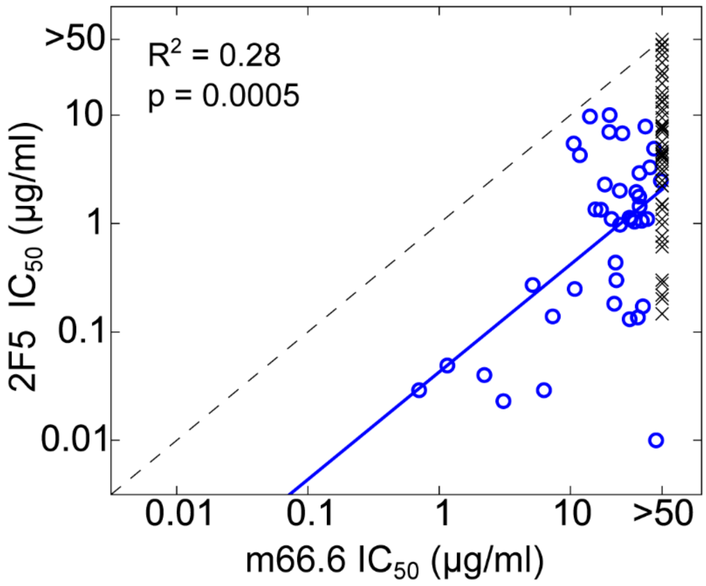

| | | 2F5 | |
|---|---|---|---|
| | | sen. | res. |
| m66.6 | sen. | 40 | 0 |
| | res. | 50 | 74 |

Accuracy = 0.70
Odds ratio = inf
p = 2.5 x $10^{-13}$

**Figure S4. Comparison of Neutralization Profiles for DH1317.4 vs. 2F5 (a), DH1317.4 vs. m66.6 (b) and 2F5 vs. m66.6 (c)**. The scatter plots on the left show IC50 titers for each heterologous virus tested. Blue circles indicate commonly sensitive viruses (IC50 < 50μg/ml for both bnAbs), black crosses indicate viruses sensitive to one or both bnAbs. Black dashed line shows identity, and blue line is the best-fit linear regression line fit using only viruses sensitive to both bnAbs being compared. Squared Pearson correlation coefficient (R2) and p-values from Pearson correlation test are indicated. For (a), filled blue circles indicate viruses that had IC50 > 40μg/ml for DH1317.4 and included some outlier points. These were excluded for linear regression. In each panel, the contingency table on the right shows the number of viruses sensitive (sen) to both bnAbs (1st row, 1st column), sensitive to either one but resistant to other (off-diagonal) or resistant to both bnAbs being compared (2nd row, 2nd column). These contingency tables were subjected to Fisher's exact test to calculate the statistical significance of the overlap of sensitive/resistant viruses for each pair of bnAbs. The *p*-values from these are indicated below each table with odds ratio and the metric accuracy that is defined as the fraction of viruses for which both bnAbs show sensitive or resistance phenotype simultaneously.

### Figure S5

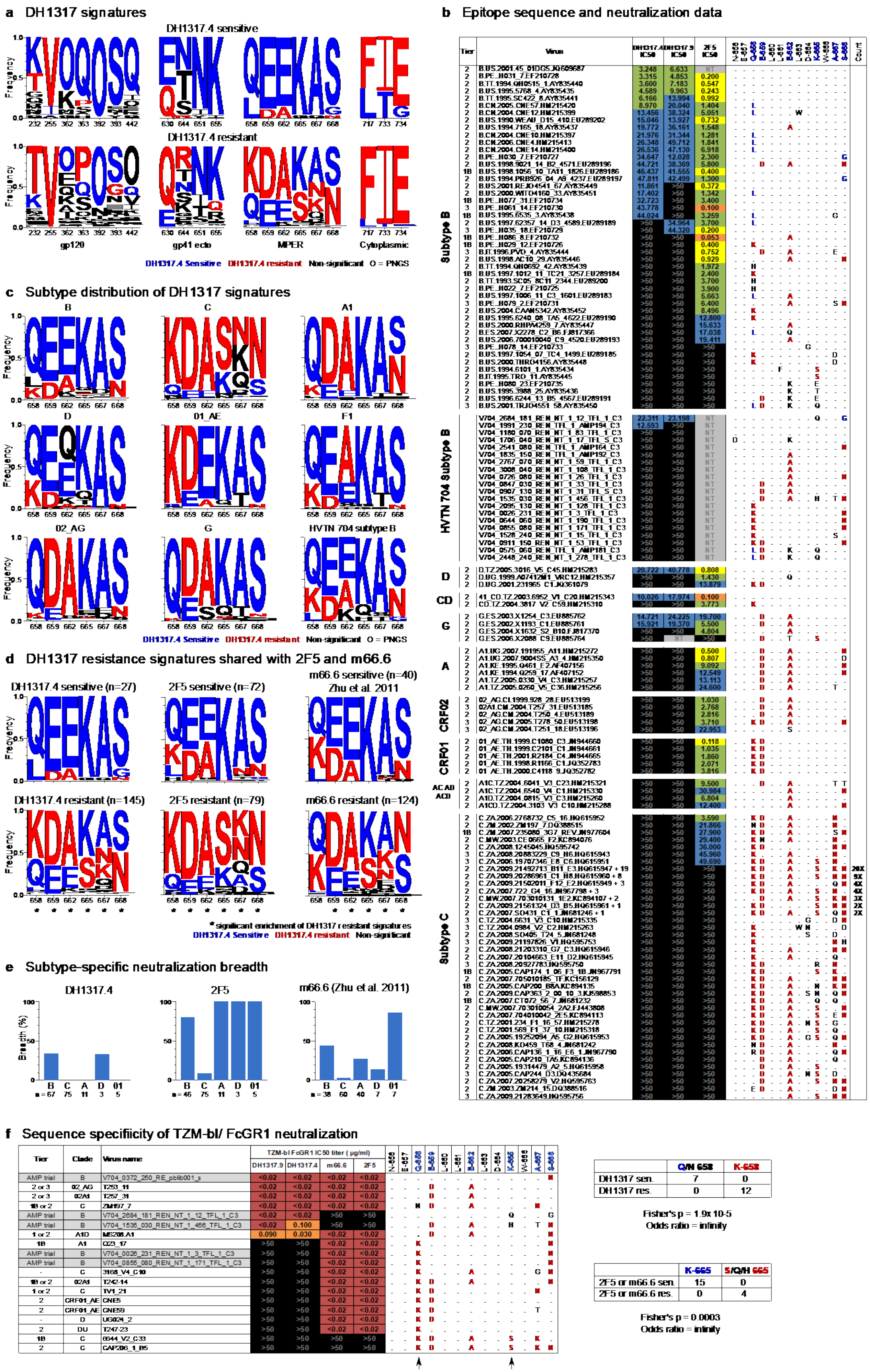

| | Q/N 658 | K-658 |
|---|---|---|
| DH1317 sen. | 7 | 0 |
| DH1317 res. | 0 | 12 |

Fisher's p = 1.9x 10-5
Odds ratio = infinity

| | K-665 | S/Q/H 665 |
|---|---|---|
| 2F5 or m66.6 sen. | 15 | 0 |
| 2F5 or m66.6 res. | 0 | 4 |

Fisher's p = 0.0003
Odds ratio = infinity

**Figure S5. (a) DH1317 signatures.** Amino acid frequencies in key Env signature sites identified for DH1317.4 neutralization are shown as sequence logos for the group of heterologous viruses that were sensitive to DH1317.4 on the top row and the resistant viruses on the bottom row. Signature sites are categorized according to the Env region they fall in. For each logo, the height of the letter corresponds to the frequency of that amino acid or of a potential N-linked glycosylation site (denoted as "O") in the group of viruses. The amino acids and glycans are color-coded according to their association with sensitivity to neutralization DH1317.4 (blue) and with resistance (red). Black letters indicate no significant association with either sensitivity or resistance. The signature sites were identical for DH1317.9 and sequence logos were virtually identical to DH1317.4 and hence are not shown. **(b) Epitope sequence and neutralization data.** Viruses are grouped according to the subtype, with more contemporary HVTN 704 subtype B viruses grouped separately from other subtype B viruses. Each row shows the tier (when known), name and IC50 titers for DH1317.4, DH1317.9 and 2F5. $IC_{50}$ titer cells are color coded from warmer to cooler shades indicating higher to lower neutralization sensitivity, and black cells indicate above the threshold of detection, $IC_{50}$ > 50μg/ml. The next columns show the sequence of the virus for HXB2 sites from 656 to 683. The sequence of the MPER peptide is used as reference and matching amino acids to the peptide are indicated by dots. Color-coding for amino acids from panel (a) is used. For subtype C viruses that are fully resistant to all 3 antibodies, recurring identical patterns that are shared by multiple sequences are indicated by the number of times they are repeated in the data in the rightmost column (e.g. "20X" means 20 distinct viruses hand shared this pattern of amino acids, and all were above the threshold of detection). **(c) Subtype distribution of DH1317 signatures.** The sequence variation at MPER signature sites from (a) are shown for different subtypes from the LANL Filtered Web reference sequence dataset using logos similar to panel (a). The last logo panel shows the sequence distribution for the 20 HVTN 704 subtype B pseudoviruses from our neutralization dataset [72]. These more contemporary B clade viruses show a enrichment for resistance amino acid signatures relative to the older B clade viruses in the standard panel. **(d) DH1317 resistance signatures shared with 2F5 and m66.6.** The top row of logos shows amino acid frequency in the viruses sensitive to DH1317.4, 2F5 and m66.6 from left to right, and the bottom row shows these in viruses that are resistant to each of these antibodies. Logos follow the same color-coding as panel (a). Resistant signatures for DH1317.4 were also significantly enriched in the resistant viruses for 2F5 and m66.6 as shown by asterisks indicating significant enrichment ($p < 0.05$, Fisher's exact test). The m66.6 neutralization data for heterologous viruses were obtained from Zhu *et al.* [30]. **(e) Subtype-specific neutralization breadth.** The breadth of neutralization at $IC_{50}$ < 50μg/ml against viruses from major subtypes is shown for DH1317.4, 2F5 and m66.6, from left to right. CRF01_AE is indicated as "01". Owing to similarity in env genes, CRF02_AG viruses are grouped with subtype A, and CRF07 / 08 are grouped with subtype C. The number of tested viruses in each subtype is shown below each subtype. **(f) Sequence specificity of TZMbl-FcγRI neutralization.** The table on the left shows the tier, clade and IC50 titers for the 19 viruses assayed in the TZMbl-FcγR1 assay (Figure 3b). Next to these are shown the amino acid sequence for each virus using the same formatting as panel (b). The contingency tables on the right show the distribution of amino acids at site 658 for DH1317.4 or DH1317.9 sensitive and resistant viruses, and at site 665 for 2F5 or m66.6 sensitive and resistant viruses. Below each, Fisher's exact test p-value and odds ratio are reported. These two sites were the only significant associations ($p < 0.05$) with TZMbl-FcGR1 neutralization sensitivity/resistance at these Env sites for either of the bnAbs.

### Figure S6

## (a)

| | HCDR1 | HCDR2 | HCDR3 | HCDR $\Delta G_{wif}$ Sum |
|---|---|---|---|---|
| IGHV7-4-1 | SGYTFTSYAMN | WINTNTGNPTYAQGFTG | RVRRGGYNN---------YYMDV | 1.05 |
| DH1317.4 | .........I. | .......I..S..AL.. | R.RR.....---------..... | 0.32 |
| DH1317.8 | ......D..IT | ..........S...... | R.RL.....---------..... | 1.52 |
| DH1351.1 | ........... | ................. | R.RRVPARGGWFDP--------- | 2.16 |
| DH1347.1 | .....IN.... | ................. | RGRAAAAGTFHIFSYFDL---- | -1.93 |
| DH1322.1 | ...N...H.L. | ................. | R.RFH.SGSYPQPADYY..... | 0.71 |
| DH1346 | .....NN.... | ...........G..... | RFRVAPAAQRGYF---Y..... | -0.89 |
| DH1319.1 | ......D...S | .............A... | RFRR.VGVLYAP-----..S.Y | -0.35 |
| IGHV5-51 | | | | |
| m66 | SGYNFASEWIG | IIYPGDSDTKYSPSFQG | RQNHYGSGSYFYRTAYYYAMDV | -1.21 |

## (b)

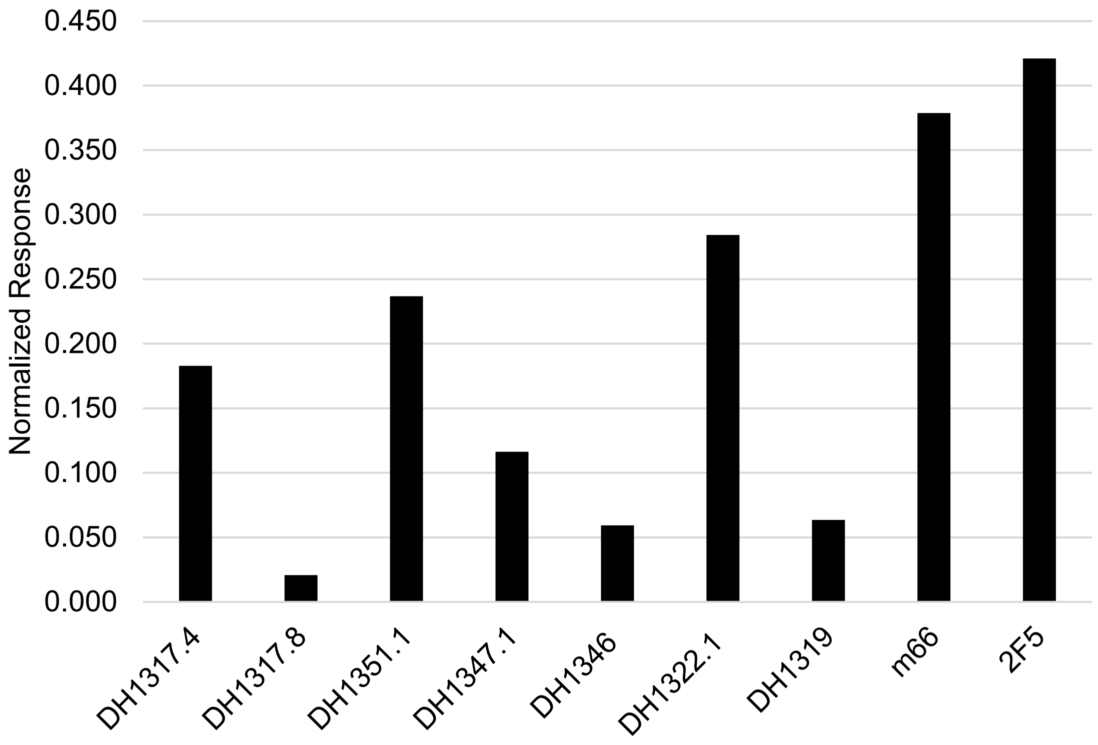

**Figure S6. HVTN133 $V_H$ 7-4-1 Abs $V_H$ CDR Sequences and Binding to Phosphatidylglycerol (PG) Liposomes (POP:-DOPG). (a)** HCDR sequences and $\Delta G_{wif}$ (free energy change of lipid insertion) of the sum of HCDRs of IGHV7-4-1 neutralizing antibody members. The putative lipid head group binding CDRH1 motif (4E10 CDRH1 site 1) [33] is underlined. HCDR3 Arg (R)-residues including R95 (a critical residue for 2F5 neutralization) [32] are in red. HCDR sequences of m66 (IGHV5-51) and $\Delta G_{wif}$ sum value are shown for comparison. **(b)** Binding responses of each of the representative $V_H$7-4-1 mAbs to PG containing liposomes measured on BLI sensor tips immobilized with POPC:DOPG (PC:PG) liposomes. Binding responses were normalized to liposome capture level (Normalized Response) and non-specific responses subtracted on each sensor tips.

### Figure S7

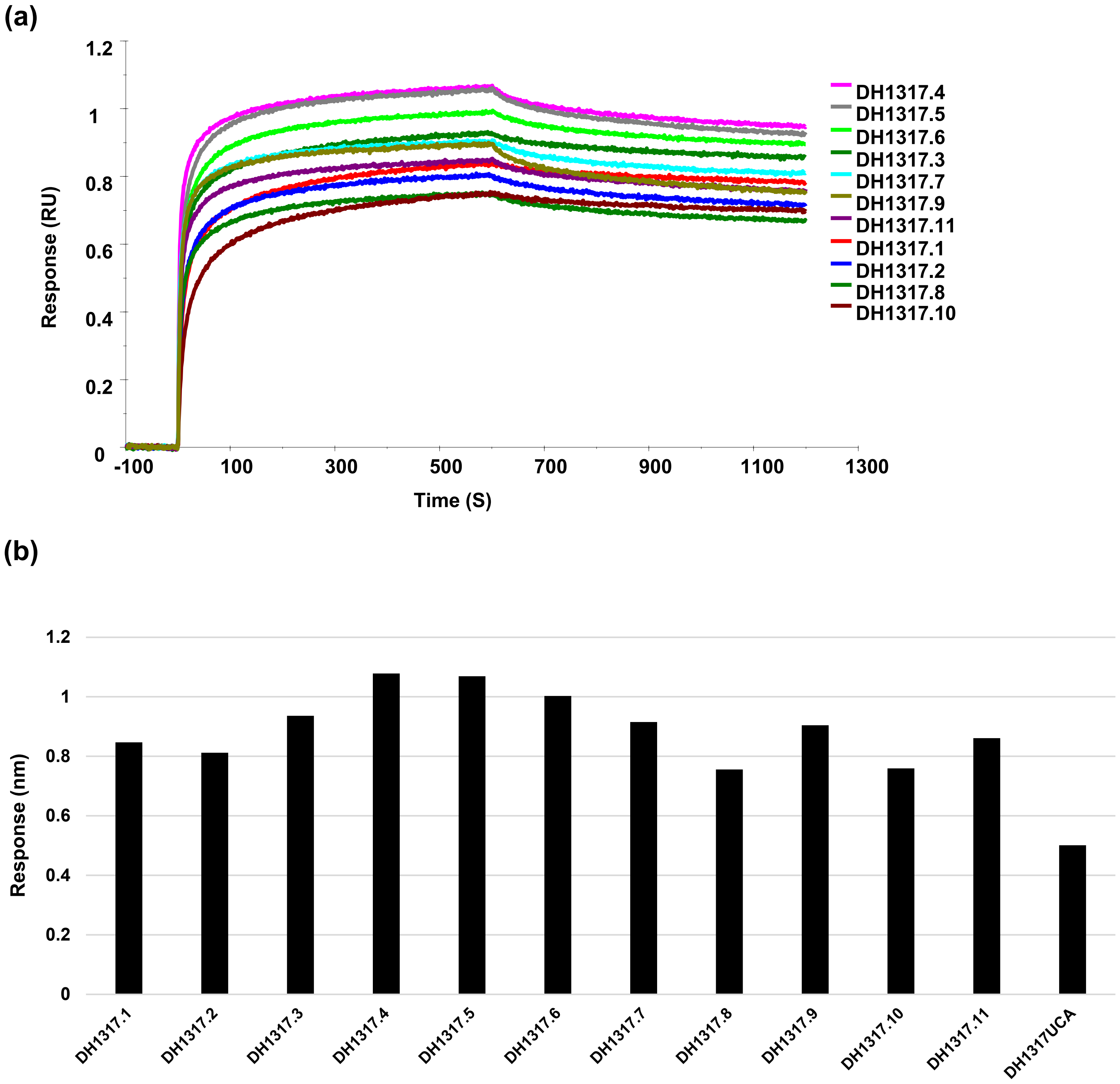

**Figure S7. Binding of DH1317 Lineage Antibodies to MPER Liposomes**. (**a**) Overlay of BLI binding curves of each indicated antibody (Ab) injected over sensors immobilized (hydrophobic APS) with MPER liposomes and following subtraction of non-specific signal over a control Ab surface. (**b**) Plot of binding responses measured at the end of the association phase in each of binding curves shown.

### Figure S8

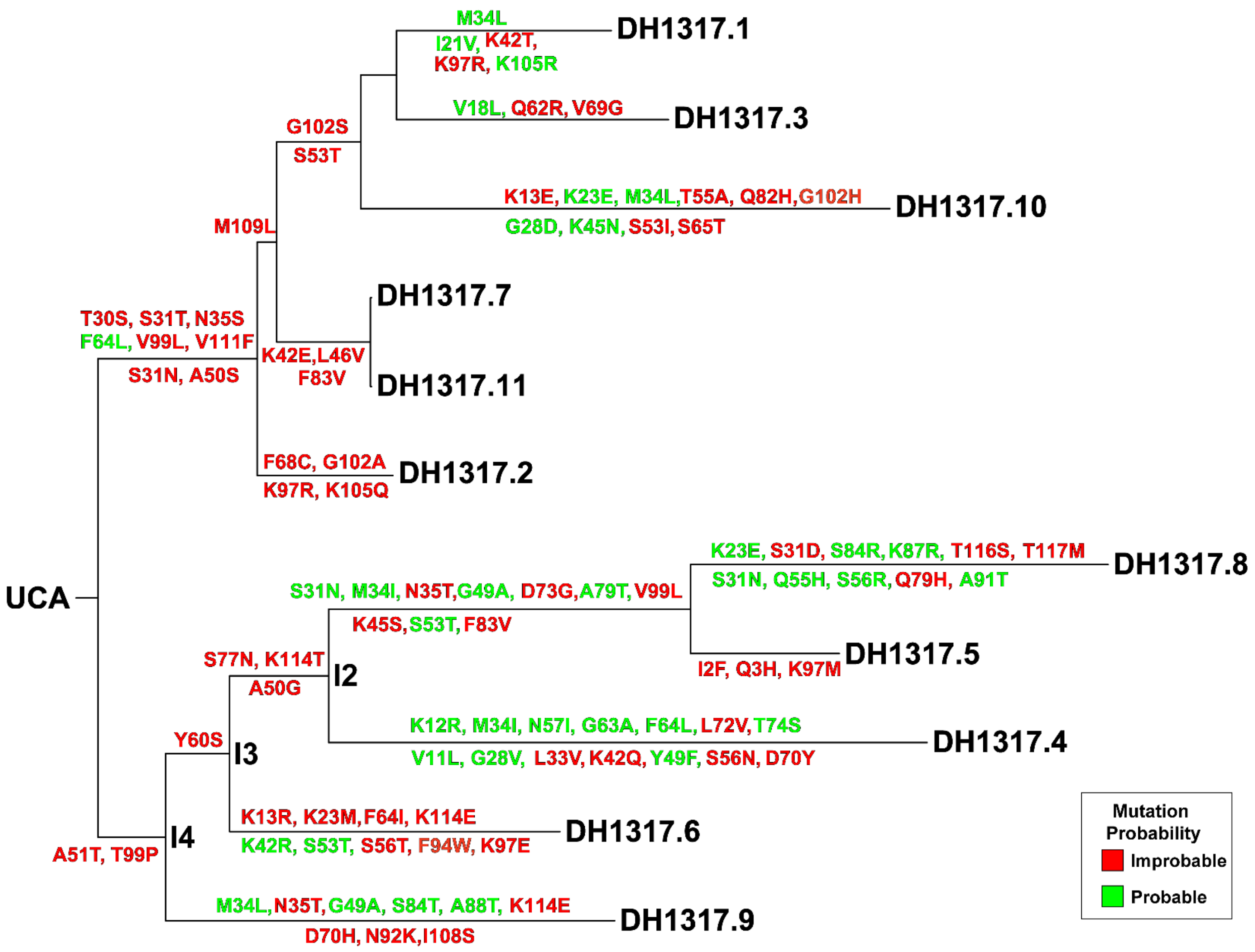

**Figure S8. Improbable Mutations in DH1317 Lineage Antibodies.** The computationally reconstructed genealogical tree of the DH1317 clone is shown with each amino acid mutation labeled along each branch of the tree and colored based on its estimated probability. Mutation probability is estimated using the computational program ARMADiLLO with the probability conditioned on the DH1317 UCA sequence and the total number of nucleotide sites mutated in the intermediate or mature DH1317 member in which the mutation occurs. Improbable mutations are defined as mutations with estimated probability of <2% and colored red. Probable mutations are estimated at >=2% probability and shown in green. The functions of the improbable mutations acquired in the intermediate antibodies (I4 to I2) listed along the evolutionary pathway to mature DH1317.4 bnAb are shown (see Figure S9).

### Figure S9

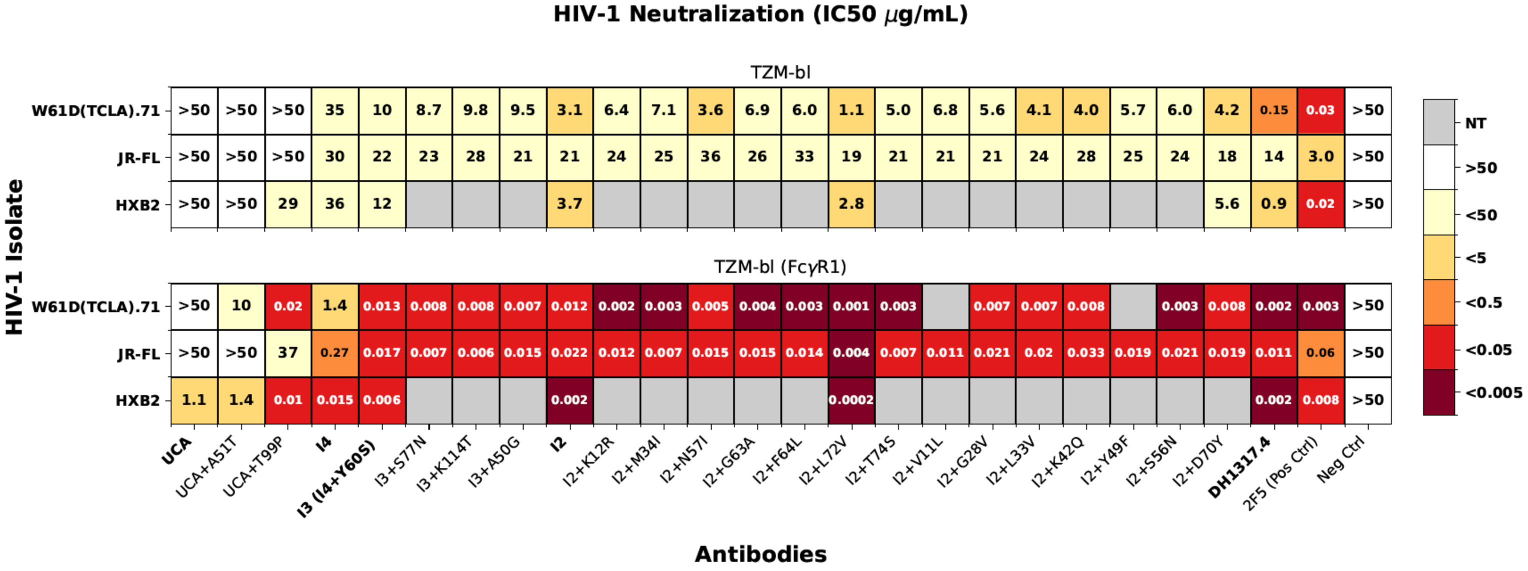

**Figure S9. Neutralization titers of wild-type and mutant DH1317 antibodies.** Recombinant mAbs bearing DH1317 genes and mutations that were acquired along the maturation pathway from the UCA to the mature DH1317.4 were tested for neutralization in the standard TZM-bl (top panel) and TZM-bl/FcγRI (bottom panel) assays. Neutralization titer was reported in IC50, μg/ml as shown in the key for the heatmap. NT: not tested.

### Figure S10

(a)

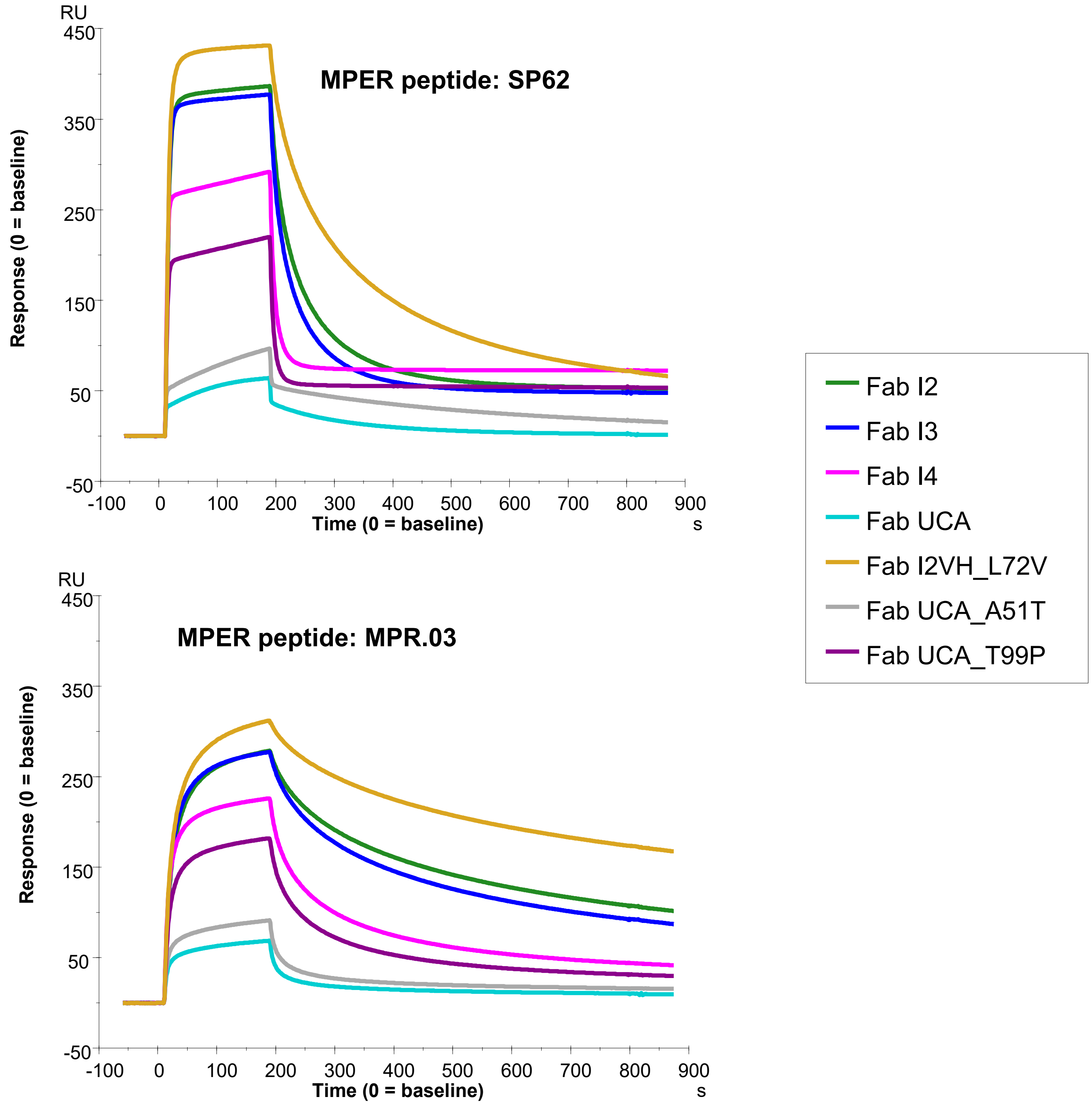

(b)

| | SP62 | | | MPR.03 | | |
|---|---|---|---|---|---|---|
| Fab DH1317… | $k_a$ (1/Ms) | $k_d$ (1/M) | $K_D$ (nM) | $k_a$ (1/Ms) | $k_d$ (1/M) | $K_D$ (nM) |
| UCA | 4.9E+05 | 9.0E-01 | 1,830 | 1.8E+06 | 9.2E-02 | 49.9 |
| UCA A51T | 6.2E+05 | 7.1E-01 | 1,140 | 3.9E+06 | 9.6E-02 | 24.8 |
| UCA T99P | 4.2E+06 | 2.4E-01 | 56.5 | 3.5E+06 | 2.3E-02 | 6.61 |
| I4 | >1.0E+07 | 2.2E-01 | 21.7 | 6.5E+06 | 2.1E-02 | 3.26 |
| I3 | 9.0E+06 | 4.1E-02 | 4.52 | 6.8E+06 | 7.6E-03 | 1.12 |
| I2 | 3.9E+06 | 2.8E-02 | 7.26 | 5.0E+06 | 7.8E-03 | 1.57 |
| I2 L72V | 3.7E+06 | 1.4E-02 | 3.74 | 5.8E+06 | 4.0E-03 | 0.689 |
| DH1317.4 | 3.5E+06 | 8.8E-03 | 2.50 | 8.0E+06 | 2.0E-03 | 0.246 |

**Figure S10. DH1317 Lineage and Mutant Fabs Binding to MPER Peptides. (a)** Surface Plasmon Resonance (SPR) measurements of binding of Fabs to biotinylated MPER peptides, MPER.03 (right) and SP62 (left) immobilized on sensor chip. DH1317UCA and listed intermediate (I) and mutant Fabs were injected at 5 ug/ml and binding curves were overlaid. **(b)** Affinity ($K_D$) and kinetic rates (association rate, $k_a$; dissociation rate, $k_d$) of binding of DH1317 Fabs to MPER peptides (SP62- left panel; MPR.03- right panel).Tabulated affinity data is representative of 2 independent titrations. Titrations were performed through single cycle kinetics at concentrations ranging from 0.25 – 64 nM (2-fold dilution series). Heterogeneous ligand model was used for curve fitting and faster kinetic parameters are reported.

### Figure S11

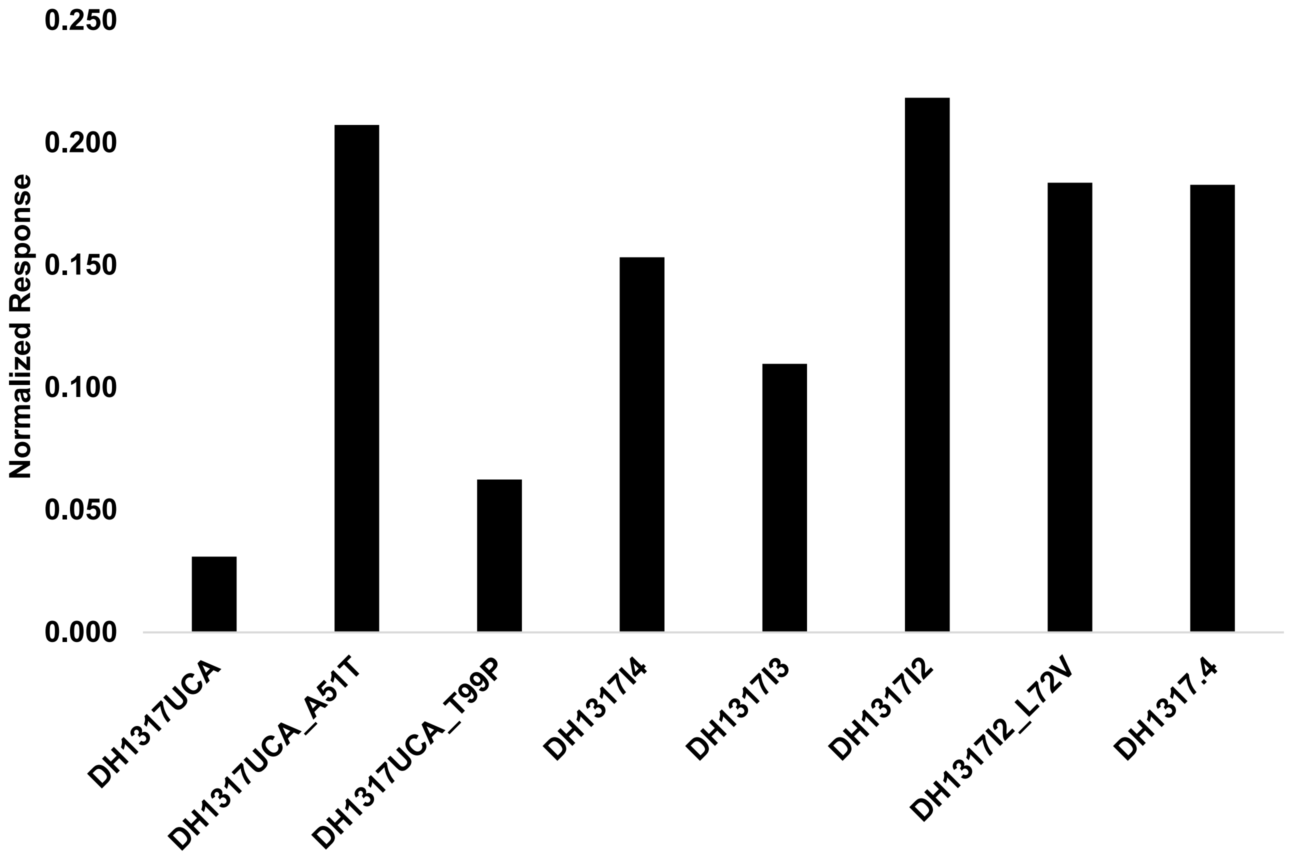

**Figure S11. DH1317 Lineage and Mutant MAbs Binding to Phosphatidyl Glycerol (PG) containing Liposomes.** Binding of each indicated mAb at 100ug/mL to POPC:DOPG (25:75) liposomes were measured by surface plasmon resonance (SPR) and plotted data show binding responses in RU normalized to control surface (lipophilic linker) and capture level of liposomes. Values represent the average of at least two measurements.

### Figure S12

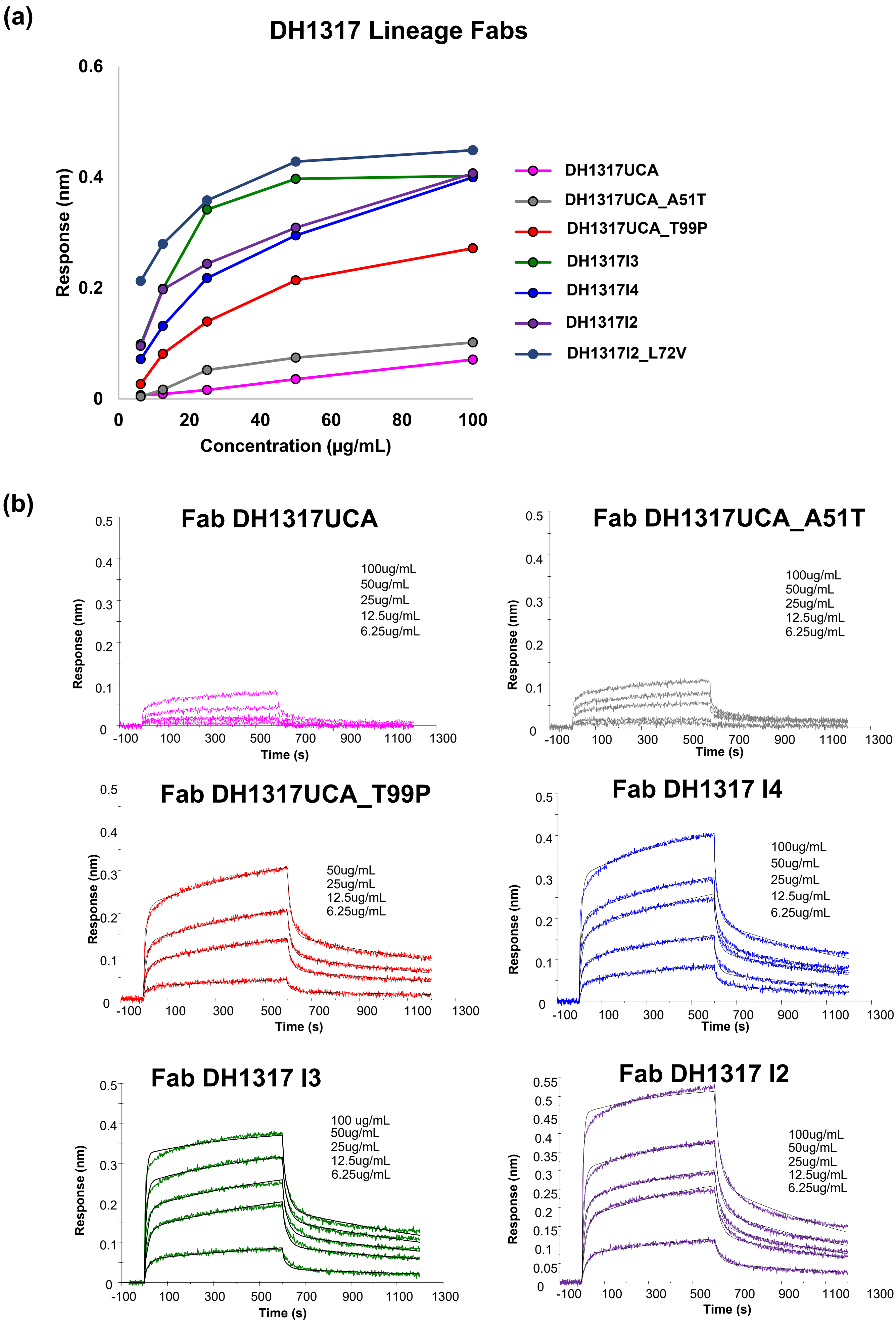

(a)
DH1317 Lineage Fabs
Response (nm)
Concentration (µg/mL)
DH1317UCA
DH1317UCA_A51T
DH1317UCA_T99P
DH1317I3
DH1317I4
DH1317I2
DH1317I2_L72V
(b)
Fab DH1317UCA
100ug/mL
50ug/mL
25ug/mL
12.5ug/mL
6.25ug/mL
Fab DH1317UCA_A51T
Fab DH1317UCA_T99P
Fab DH1317 I4
Fab DH1317 I3
100 ug/mL
Fab DH1317 I2
Response (nm)
Time (s)

### Figure S12 (cont.d)

**(b)**

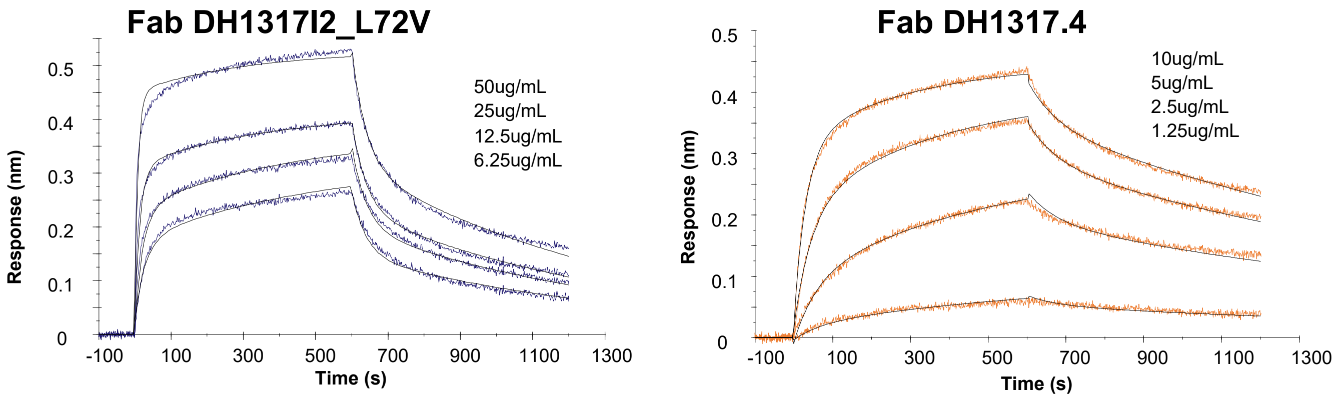

**Figure S12. DH1317 Lineage and Mutant Fabs Binding to MPER Liposomes. (a)** Biolayer interferometry (BLI) binding responses to MPER liposomes of indicated DH1317 lineage and mutant Fabs at concentrations ranging from 6.25ug/mL to 100ug/mL (2-fold dilution series). Binding responses (nm) were measured at the end of the Fab association phase from 590-595s. **(b)** BLI binding curves of DH1317 lineage and mutant Fabs from 6.25ug/mL to 100ug/mL to MPER liposomes. DH1317UCA_T99P, DH1317I4, DH1317I3, DH1317I2, DH1317I2_L72V, and DH1317.4 Fabs were all fitted to the 2-step conformation change model (overlaid black fit) and are from a single measurement.

### Figure S13

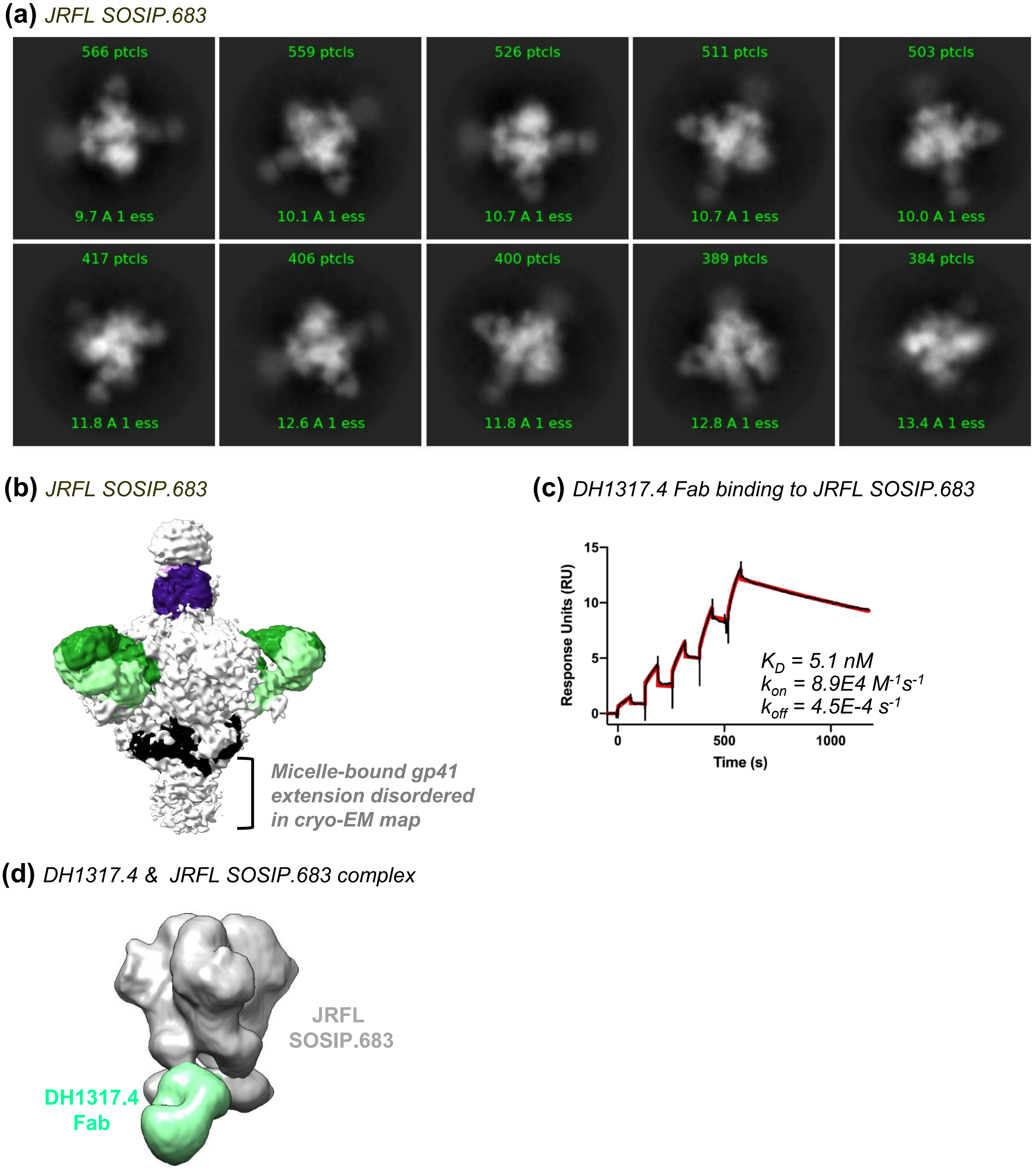

**Figure S13. DH1317 Lineage Abs Binding to JRFL Gp150 in Detergent Micelles in SPR and NSEM. (a)** Raw cryoEM images and low resolution class average of the JRFL SOSIP+ MPER trimer in bound to PGT145 and VRC01 bnAbs for stabilization. **(b)** 3D reconstruction of JRFL gp150 trimer expressed in lipid bicelles and stabilized by PGT145 and VRC01 bnAbs. **(c)** SPR sensorgram of dose titration of tier 2 nAb DH1317.4 Fab binding to JRFL trimer with affinity of 5.1 nM. **(d)** DH1317.4 bound to JRFL SOSIP bearing MPER in detergent micelles in negative stained EM (NSEM) class average.

### Figure S14

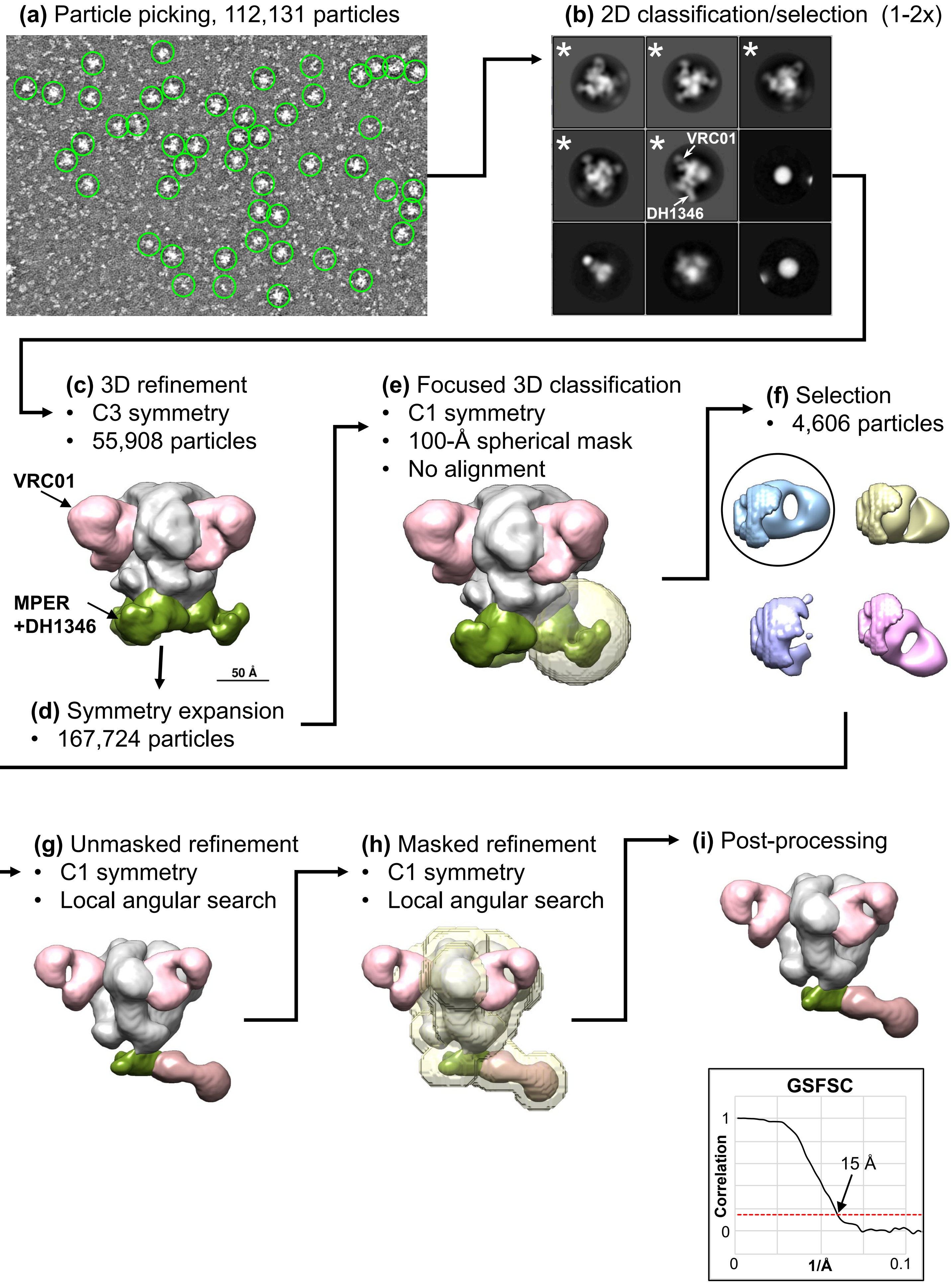
(a) Particle picking, 112,131 particles
(b) 2D classification/selection (1-2x)
VRC01
DH1346
(c) 3D refinement
• C3 symmetry
• 55,908 particles
VRC01
MPER +DH1346
50 Å
(d) Symmetry expansion
• 167,724 particles
(e) Focused 3D classification
• C1 symmetry
• 100-Å spherical mask
• No alignment
(f) Selection
• 4,606 particles
(g) Unmasked refinement
• C1 symmetry
• Local angular search
(h) Masked refinement
• C1 symmetry
• Local angular search
(i) Post-processing
GSFSC
Correlation
1
0
15 Å
0
1/Å
0.1

**Figure S14. Sample Data Processing Flow for NSEM Structures of MPER-directed Antibodies Complexed with JRFL SOSIP.683. (a)** Sample electron micrograph from the DH1346 complex showing automatically picked particles (green circles). **(b)** Sample 2D class averages from classification of extracted particles. The center class shows a SOSIP side view, in which densities corresponding to VRC01 and the MPER-directed mAb DH1346 can be identified (arrows). 2D classification was used to select the particles in classes showing Fab-bound SOSIPS (stars). For some samples, a second round of 2D classification/selection was performed. **(c)** Selected particles were then subjected to a 3D refinement with C3 symmetry imposed. At this stage, the SOSIP trimer (gray) and the VRC01 Fabs (pink) are well-resolved. The map is shown at a low contour level to display the fainter density for the poorly-resolved MPER/DH1346 Fab region (green). **(d)** Refined particles were then symmetry expanded to give three times as many as were originally selected. From this point forward all processing was done using no alignment, or with local angular searches only, to prevent particles from reverting to their original orientation and creating duplicates. This ultimately has the effect of aligning the DH1346 Fabs onto a single protomer as will be seen in G-I. **(e)** A 100-Å diameter spherical mask with a 3-pixel soft edge (transparent sphere) was then centered on one DH1346 Fab density and used for a focused 3D classification with no alignment. **(f)** Sample classes from the focused 3D classification show density within the 100-Å spherical mask. The blue and pink classes display well-resolved Fab-shaped densities. Note, however, their highly divergent angles, presumably indicating flexibility of the MPER domain. The gold-colored class is roughly Fab-shaped, but misshapen, presumably due to poor angular sampling or low particle count. The purple class is a "junk" class that does not contain any Fab-shaped density. For samples such as this one that showed at least one well-resolved Fab-shaped classes at this stage, a single class was selected (circle) and processed as described in G-I. Some samples, generally those that showed weaker binding by NSEM, showed only poorly-resolved Fabs at this stage, in which case all classes with vaguely Fab-shaped density were selected, refined as described in G below, and then subjected to a second round of focused 3D classification as described in E and F, and then a single class chosen for further processing. **(g)** Final selected particles from the focused 3D classification were subjected to an unmasked 3D refinement using local angular searches only. **(h)** The resultant map from G was used to create a shaped, soft-edge mask that contained the SOSIP trimer and MPER Fab but excluded the VRC01 Fabs (transparent surface) and used in a second refinement using local angular searches only. **(i)** The final map was then post-processed. The graph indicates the gold-standard Fourier shell correlation of the unmasked map, with the dotted line indicating the 0.143 cutoff level.

### Figure S15

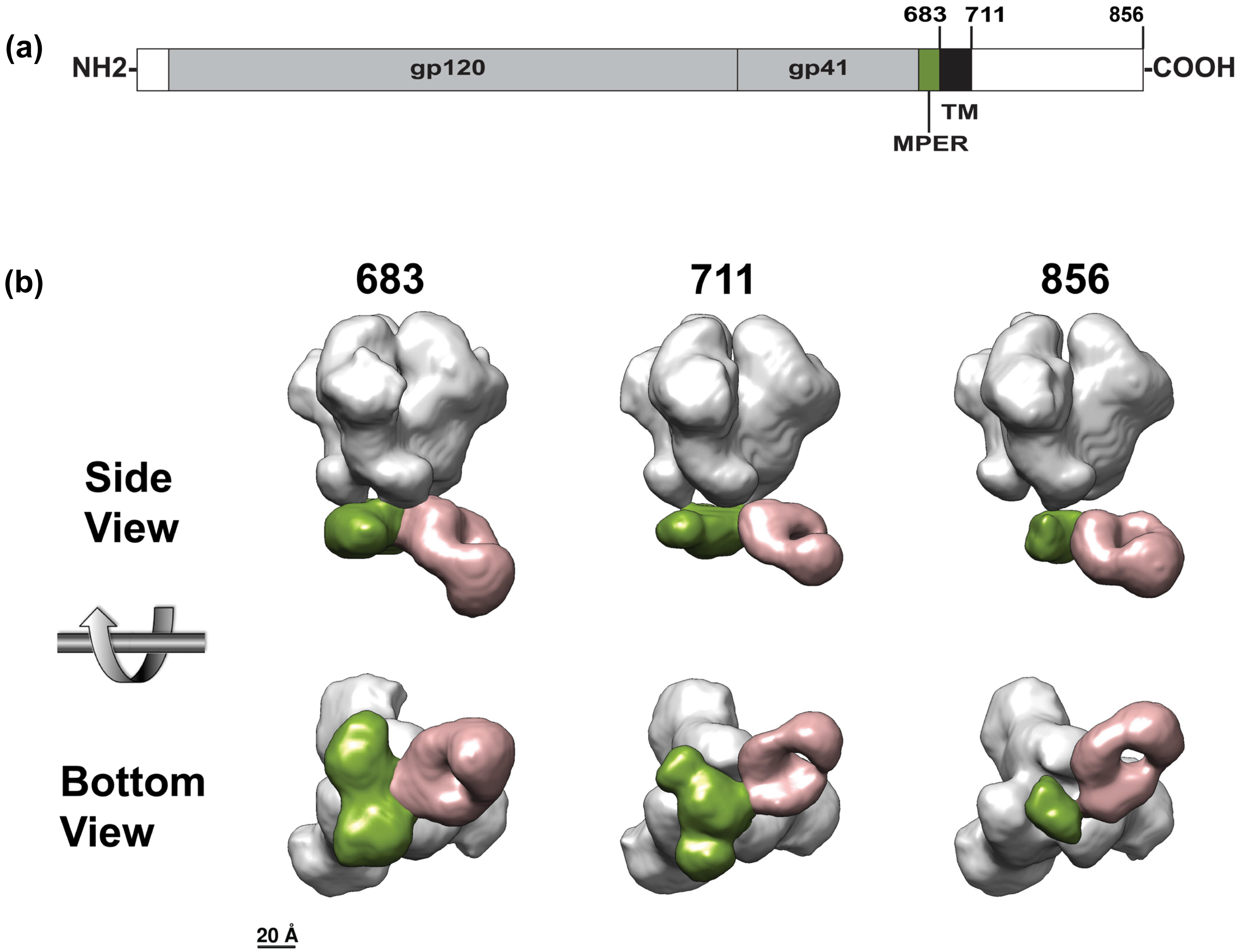

**Figure S15. NSEM Comparison of DH1317.4 Binding to Truncated versus Full-length JRFL Constructs. (a)** Schematic diagram of Env domain structure with locations of truncations indicated at residue 683 (MPER only), residue 711 (MPER + transmembrane domains), or residue 856 (full-length). **(b)** NSEM maps in side and bottom views showing DH1317.4 (brown) bound to JRFL constructs of three different lengths. The last included residue in each construct is indicated by the number above.

**Figure S16**

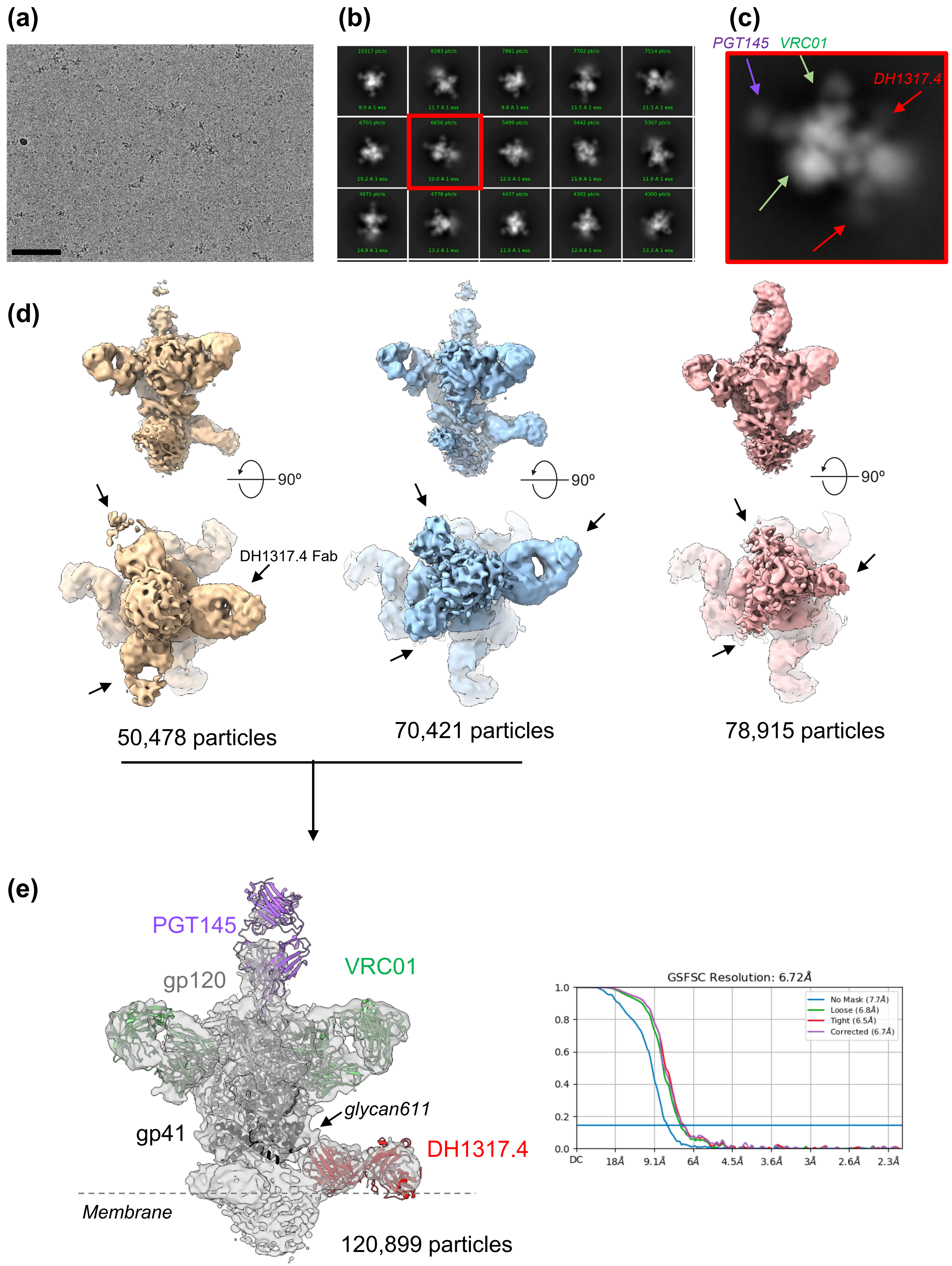
(a)
(b)
(c)
PGT145
VRC01
DH1317.4
(d)
90º
90º
90º
DH1317.4 Fab
50,478 particles
70,421 particles
78,915 particles
(e)
PGT145
VRC01
gp120
gp41
glycan611
DH1317.4
Membrane
120,899 particles
GSFSC Resolution: 6.72Å
No Mask (7.7Å)
Loose (6.8Å)
Tight (6.5Å)
Corrected (6.7Å)
1.0
0.8
0.6
0.4
0.2
0.0
DC
18Å
9.1Å
6Å
4.5Å
3.6Å
3Å
2.6Å
2.3Å

**Figure S16. Cryo-EM structure of DH1317.4 bound to HIV-1 Env JRFL full length SOSIP trimer**. **(a)** Representative micrograph. Scale bar represents 100 nm. **(b)** Representative reference-free 2D classes. **(c)** Zoomed-in view of the class average outlined with a red square in panel b, showing a side view of the Env bound to PGT145 (purple arrow), VRC01 (green arrows) and DH1317.4 (red arrows) Fabs. **(d)** 3D reconstruction of 3 distinct populations of the DH1317.4-bound Env complex. The arrows indicate the positions of the bound DH1317.4 Fabs. **(e)** Left, 3D reconstruction shown as a transparent grey surface with underlying fitted model shown in cartoon representation, with gp120 colored light grey, gp41 black, PGT145 purple, VRC01 green and DH1317.4 red. An arrow points to the density corresponding to the base of Env gp41 glycan 611. Right, Fourier Shell Correlation (FSC) plot. $FSC_{0.143}$ is indicated by a dotted line.

### Figure S17

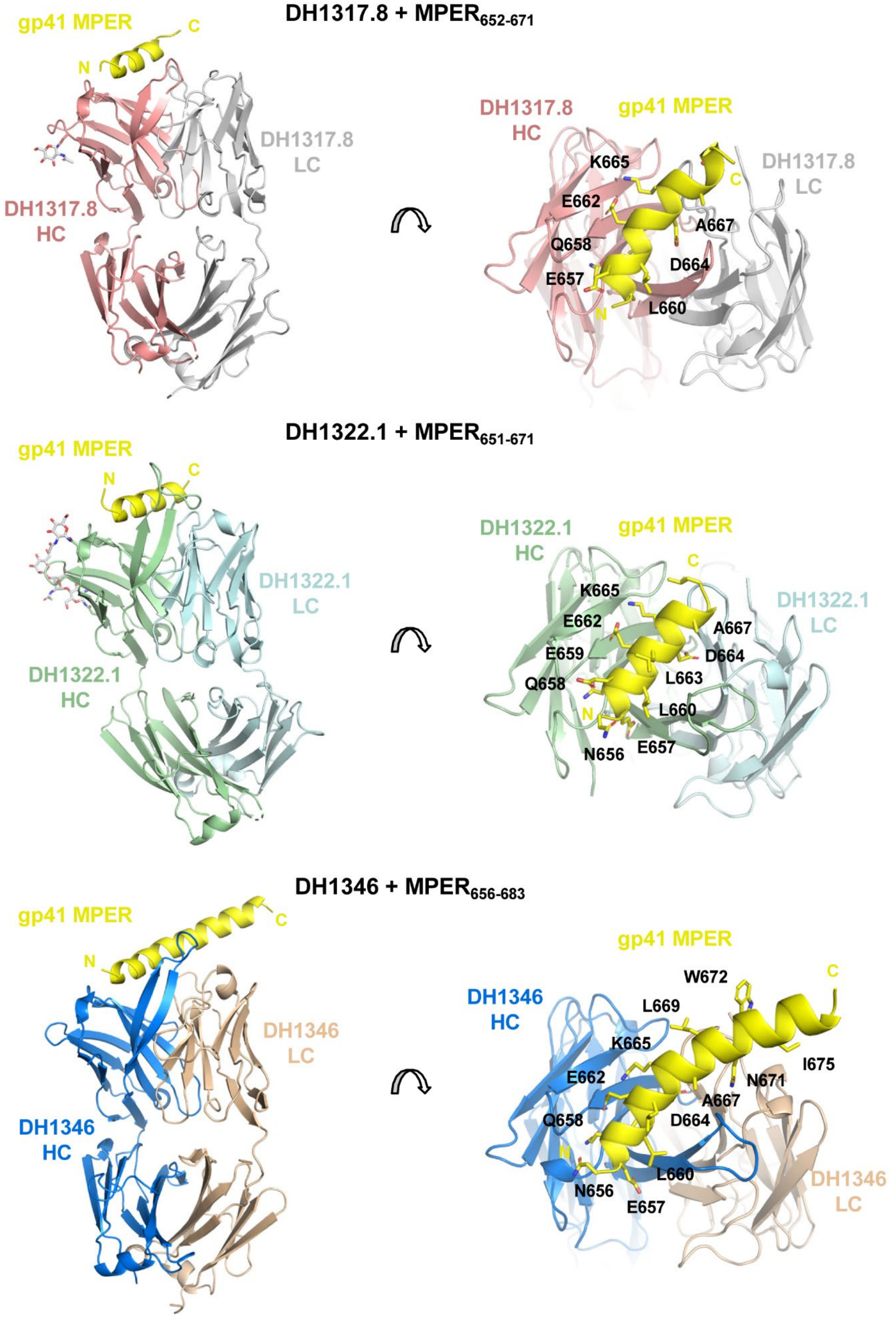

**Figure S17. Fab Crystal Structures of Polyclonal $V_H$7-4-1 Antibodies in Complex with MPER Peptides.** Crystal structures of the Fabs of $V_H$7-4-1-using neutralizing antibodies DH1317.8, DH1322.1, and DH1346 in complex with MPER proximal peptides 652-671 (DH1317.8), 651-671 (DH1322.1), and 656-683 (DH1346). Rotated closeup views are shown in the panels on the right, with MPER residues that interact with each neutralizing antibody labeled and shown in stick representation. N-linked glycans shown as gray sticks (left).

### Figure S18

(a)

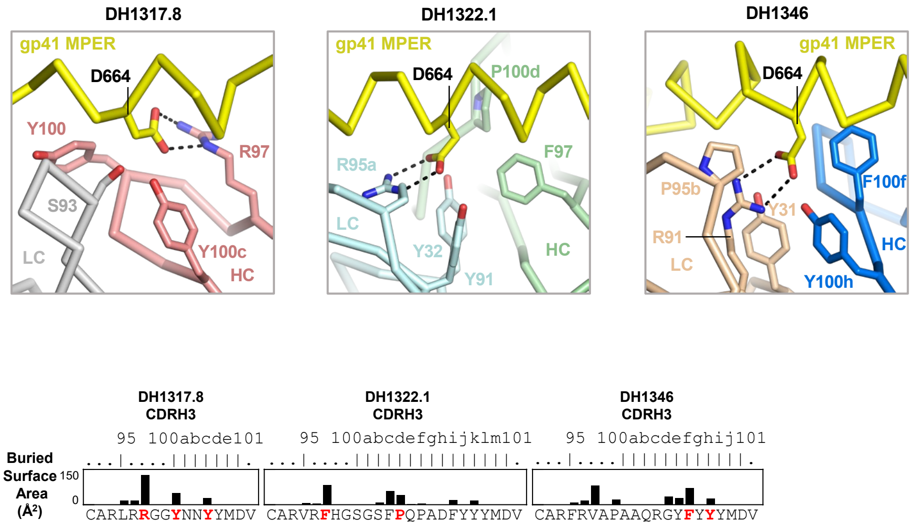

(b)

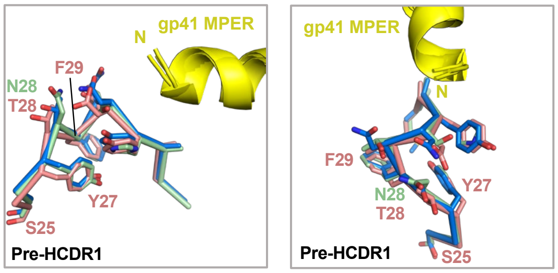

**Figure S18. Structural Features of Polyclonal $V_H$7-4-1 Antibody Interactions with MPER. (a)** Neutralizing antibody interactions with MPER residue D664 are mediated by antibody HCDR3, LCDR3, and LCDR1 residues, as shown. Buried surface areas on HCDR3 loop residues of DH1317.8, DH1322.1, and DH1346 are plotted as bars, with residues that contact D664 labeled in red. **(b)** Structural alignment of a putative lipid interacting that falls immediately prior to HCDR1 (Pre-HCDR1), shown in two orientations.

**Table S1. Tally of MPER+ Antibodies in HVTN 133 Vaccine Trial Participants**

| PTID | G# | B cells (FACS) | | Abs | | | | | | | | |
|---|---|---|---|---|---|---|---|---|---|---|---|---|
| | | Total | | TOTAL | | | MPER (+) | | MPERΔ (WT+, KO-) | | MPER Non-Δ | |
| | | Baseline | Post 3rd IMM | Pre/ Post IMM | Baseline | Post 3rd IMM | Baseline | Post 3rd IMM | Baseline | Post 3rd IMM | Baseline | Post 3rd IMM |
| 133-23 | 1 | 379,168 | 517,977 | 111 | 30 | 81 | 1 (3%) | 59 (73%) | 0 (0%) | 49 (83%) | *und* | 10 (17%) |
| 133-39 | 1 | 565,666 | 572,110 | 93 | 48 | 45 | 0 (0%) | 27 (60%) | 0 (0%) | 26 (96%) | 0 (0%) | 1 (4%) |
| 133-35 | 1 | 226,585 | 462,296 | 206 | 105 | 101 | 0 (0%) | 0 (0%) | 0 (0%) | 0 (0%) | 0 (0%) | 0 (0%) |
| 133-33 | 2 | 587,188 | 412,878 | 377 | 192 | 185 | 0 (0%) | 0 (0%) | 0 (0%) | 0 (0%) | 0 (0%) | 0 (0%) |
| 133-30^ | 2 | 504,105 | 522,629 | 143 | 45 | 98 | 1 (2%) | 1 (1%) | 0 (0%) | 0 (0%) | *und* | 1 (100%) |
| Summary | | 2,262,712 | 2,487,890 | 930 | 420 | 510 | 2 (0.5%) | 87 (17%) | 0 (0%) | 75 (86%) | 0 | 12 (14%) |

Shown in this table are the numbers and frequencies of MPER+ Abs that were isolated from five HVTN 133 vaccine trial participants who received three immunizations; participants in vaccine group (G#) 1 received 500mcg (low dose) and those in group 2 received 2000mg (high dose). Total CD19+ B cells indicated from each participant at pre- (baseline) and post 3rd immunizations were interrogated for MPER reactivity via FACS. The BCRs from MPER+ Abs were expressed recombinantly as purified IgG or monoclonal Abs and tested for binding via ELISA; positivity cutoff in antibodyome ELISA with purified IgG ≥ 0.5 $OD_{450}$. The tally and frequencies of MPER+ Abs are indicated for each subject; red – percent of total Abs at baseline or post 3rd immunization; blue – percent of MPER+ Abs at baseline or post 3rd immunization. MPER+ Abs demonstrated binding to MPER656, MPER.03 and/or SP62 peptide. MPER-differential (Δ) binding Abs had ≥2.5 fold difference in EC50 for binding to wild-type MPER.03 peptide, but not the knock-out mutant (D664AW672A), whereas MPER non-differential (Non-Δ) binding Abs had similar binding levels to both peptides. ^133-30 received three immunizations, but provided peripheral blood at month 12/ visit 9 instead of Month 6.5 or visit 7. Und: undetermined – Abs had low level binding to vaccine-matched MPER656 peptide, but not MPER.03.

**Table S2. Frequency of MPERΔ BnAb-phenotype Antibodies in Total Vaccine-recipients Post-3rd Immunization**

| Vaccine recipients | Vaccine group | Timepoint | Total B cells | Total MPER+ antibodies (% of B cells) | MPERΔ bnAb-phenotype Abs | |
|---|---|---|---|---|---|---|
| | | | | | Total (% of B cells) | Frequency |
| 133-23 | 1 | Visit 7 | 517,977 | 59 (0.01%) | 49 (0.009%) | 90 per 1M |
| 133-39 | 1 | Visit 7 | 572,110 | 27 (0.005%) | 26 (0.004%) | 40 per 1M |
| 133-35 | 1 | Visit 7 | 462,296 | 0 (0%) | 0 | 0 per 1M |
| 133-33 | 2 | Visit 7 | 412,878 | 0 (0%) | 0 | 0 per 1M |
| 133-30 | 2 | Visit 9 | 522,629 | 1 (0.0002%) | 0 | 0 per 1M |
| Total | | | 2,487,890 | 87 (0.003%) | 75 (0.003%) | 30 per 1M |

Shown in this table are the frequencies of MPERΔ bnAb-phenotype Abs among total CD19+ B cells isolated from five vaccine recipients in HVTN 133 who received the low (group 1) or high (group 2) vaccine doses. MPER+ Abs were isolated from 2 weeks post 3rd immunization in all individuals. Frequency of MPERΔ bnAb-phenotype Abs was predicted as a ratio per million total B cells based on the percent frequency of this population among total B cells studied. 90/million B cells = 1:11,111 B cells; 40/million B cells = 1:25,000 B cells; 30/million B cells = 1:33,333 B cells.

**Table S14. Cryo-EM Data Collection and Refinement Statistics.**

| Structure Name | Full-length JR-FL SOSIP Env bound to PGT145, VRC01 and DH1317.4 |
|---|---|
| **Data Collection and processing** | |
| Microscope | FEI Titan Krios |
| Detector | Gatan K3 |
| Magnification | 81000 |
| Voltage (kV) | 300 |
| Electron exposure (e-/Å^2) | 64 |
| Defocus Range (μm) | ~0.8-2.40 |
| Pixel size (Å) | 1.08 |
| Reconstruction software | cryoSPARC |
| Symmetry imposed | C1 |
| Final particle images (no.) | 120,899 |
| Map resolution (Å) | 6.7 |
| FSC threshold | 0.143 |

**Table S15. Crystallographic Data Collection and Refinement Statistics.**

| Parameter | DH1317.8 Fab + MPER (652-671) | DH1322.1 Fab + MPER (651-671) | DH1346 Fab + MPER (656-683) |
|---|---|---|---|
| **PDB ID** | 8G8A | 8G8C | 8G8D |
| **Data Collection statistics** | | | |
| Space group | $C2_1$ | $C2_1$ | $P2_12_12_1$ |
| Cell constants | | | |
| *a, b*, *c* (Å) | 165.0, 63.8, 112.0 | 75.3, 183, 84.4 | 41.1, 116, 231 |
| α, β, γ(°) | 90, 101, 90 | 90, 92.3, 90 | 90, 90, 90 |
| Wavelength (Å) | 1.0000 | 1.0000 | 1.0000 |
| Resolution (Å) | 50.0 – 2.44 (2.53 – 2.44) | 28.7 - 2.08 (2.16 - 2.08) | 36.6 – 2.03 (2.10 – 2.03) |
| *Rmerge* | 0.067 (0.61) | 0.044 (0.32) | 0.075 (0.53) |
| $I / \sigma I$ | 16.8 (1.07) | 13.0 (1.82) | 12.1 (1.3) |
| Completeness (%) | 94.8 (77.4) | 96.9 (86.6) | 91.7 (82.8) |
| Redundancy | 3.3 (2.8) | 3.8 (3.8) | 3.0 (2.5) |
| **Refinement statistics** | | | |
| Resolution (Å) | 41.9 – 2.44 (2.53 – 2.44) | 28.7 - 2.08 (2.16 - 2.08) | 36.6 – 2.03 (2.10 – 2.03) |
| Unique reflections | 40649 (3324) | 65739 (5860) | 67030 (5929) |
| *R*work / *R*free (%) | 21.7/25.1 | 22.6/24.4 | 20.0/23.6 |
| No. atoms | 6854 | 7192 | 7549 |
| Protein | 6822 | 6932 | 7120 |
| Ligand/ion | 4 | 0 | 11 |
| Water | 28 | 260 | 418 |
| *B-factors* ($Å^2$) | 82.9 | 61.1 | 45.8 |
| Protein | 83.0 | 61.4 | 45.9 |
| Ligand/ion | 64.5 | 0 | 52.8 |
| Water | 60.9 | 53.1 | 44.0 |
| R.m.s. deviations | | | |
| Bond lengths (Å) | 0.006 | 0.003 | 0.011 |
| Bond angles (°) | 0.89 | 0.56 | 0.88 |
| Ramachandran plot (%) | | | |
| Most favored regions (%) | 97.1 | 97.6 | 97.1 |
| Additional allowed regions (%) | 2.9 | 2.3 | 2.9 |
| Disallowed regions (%) | 0 | 0.11 | 0 |

Numbers in parentheses represent highest resolution shell.

**Table S16. Buried Surface Areas with Gp41 for DH1317.8, DH1322.1 and DH1346**

| | Buried surface area (Å²) | | | | | | | |
|---|---|---|---|---|---|---|---|---|
| | Antibodies | | | | | | gp41 MPER | |
| Antibody | FR1 | CDR1 | CDR2 | CDR3 | Subtotal | % | Subtotal | % |
| **DH1317.8 HC** | 1.84 | 59.41 | 179.87 | 245.25 | 486.37 | 79.0 | 606.04 | 85.5 |
| **DH1317.8 LC** | | | | 129.19 | 129.18 | 21.0 | 103.13 | 14.5 |
| **Total** | | | | | 615.55 | | 709.17 | |
| **DH1322.1 HC** | 8.83 | 68.71 | 208.63 | 279.84 | 565.03 | 72.6 | 660.21 | 76.3 |
| **DH1322.1 LC** | | 16.03 | | 197.00 | 213.03 | 27.4 | 205.23 | 23.7 |
| **Total** | | | | | 778.06 | | 865.44 | |
| **DH1346 HC** | 7.85 | 81.31 | 166.58 | 302.12 | 556.27 | 69.7 | 651.92 | 75.5 |
| **DH1346 LC** | | 47.04 | | | 241.73 | 30.3 | 211.05 | 24.5 |
| **Total** | | | | | 798.00 | | 862.97 | |

**Table S17. Illumina NGS Run Summary**

| Run | 133-23 | | | | 133-39 | | | |
|---|---|---|---|---|---|---|---|---|
| | Visit 2 | Visit 3 | Visit 5 | Visit 7 | Visit 2 | Visit 3 | Visit 5 | Visit 7 |
| Fwd Reads | 3414256 | 3518241 | 2835325 | 3698556 | 6528389 | 5435377 | 6973562 | 4916737 |
| Rev Reads | 3414256 | 3518241 | 2835325 | 3698556 | 6528389 | 5435377 | 6973562 | 4916737 |
| Merged Reads | 3140740 | 3220876 | 2656108 | 3412874 | 6007560 | 5022588 | 6442417 | 4483294 |
| Fastq quality filter | 2053459 | 2132875 | 1908198 | 2348205 | 4034830 | 3402013 | 4361682 | 2968172 |
| Unique Reads | 1759631 | 1840541 | 1627008 | 1938476 | 3463089 | 2910286 | 3743120 | 2559549 |
| Heavy Cloanalyst Unique Functional Reads | 553203 | 600586 | 416013 | 533712 | 974483 | 767187 | 1015263 | 766310 |
| Kappa Cloanalyst Unique Functional Reads | 162748 | 171977 | 157244 | 162215 | 303027 | 293439 | 342798 | 210006 |
| Lambda Cloanalyst Unique Functional Reads | 200271 | 245786 | 192327 | 223422 | 524677 | 438726 | 568729 | 337839 |
| Heavy deduplicated fxnl VDJ sequences | 389167 | 422183 | 269448 | 376805 | 736631 | 564833 | 760291 | 530804 |
| Kappa deduplicated fxnl VDJ sequences | 104707 | 112076 | 95463 | 104580 | 204881 | 192496 | 231272 | 137056 |
| Lambda deduplicated fxnl VDJ sequences | 101431 | 122800 | 83302 | 113727 | 324178 | 266421 | 351672 | 190660 |

We studied total antigen-unbiased VH sequences from B cells in PBMCs collected at visit 2 (baseline and time of immunization #1), and visits 3, 5 and 7 that were 2 weeks post 1$^{st}$, 2$^{nd}$ and 3$^{rd}$ immunizations, respectively. See methods for quality filtering, contig generation and annotation of sequences generated via Illumina NGS.

**Table S19. DH1317 Clonally-related VH Sequences with Improbable Mutations Associated with BnAb Development**

| Visit No. | Number of NGS VH reads | DH1317-like VH sequences/ Improbable Mutations | | | | | |
|---|---|---|---|---|---|---|---|
| | | Y60 | S60 | L72 | V72 | Y60+L72 | S60+V72 |
| **Visit 5** | **56** | **10** | **45** | **54** | **0** | **10** | **0** |
| **Visit 7** | **10** | **9** | **0** | **7** | **2** | **7** | **0** |

We interrogated DH1317 clonally-related VH sequences detected via Illumina NGS at visits 5 and 7, post 2nd and 3rd immunizations, respectively, for improbable mutations associated with DH1317.4 bnAb maturation. The number of unique and functional DH1317 VH reads with improbable mutations are listed in this table.
